# Severe acute respiratory syndrome (SARS) mathematical models and disease parameters: a systematic review and meta-analysis

**DOI:** 10.1101/2024.08.13.24311934

**Authors:** Christian Morgenstern, Thomas Rawson, Isobel Routledge, Mara Kont, Natsuko Imai-Eaton, Janetta Skarp, Patrick Doohan, Kelly McCain, Rob Johnson, H. Juliette T. Unwin, Tristan Naidoo, Dominic P Dee, Kanchan Parchani, Bethan N Cracknell Daniels, Anna Vicco, Kieran O. Drake, Paula Christen, Richard J Sheppard, Sequoia I Leuba, Joseph T Hicks, Ruth McCabe, Rebecca K Nash, Cosmo N Santoni, Pathogen Epidemiology Review Group, Gina Cuomo-Dannenburg, Sabine van Elsland, Sangeeta Bhatia, Anne Cori

## Abstract

We conducted a systematic review (PROSPERO CRD42023393345) of severe acute respiratory syndrome (SARS) transmission models and parameters characterising its transmission, evolution, natural history, severity, risk factors and seroprevalence. Information was extracted using a custom database and quality assessment tool.

We extracted 519 parameters, 243 risk factors, and 112 models from 288 papers. Our analyses show SARS is characterised by high lethality (case fatality ratio 10.9%), transmissibility (R_0_ range 1.1-4.59), and is prone to superspreading (20% top infectors causing up to 91% of infections). Infection risk was highest among healthcare workers and close contacts of infected individuals. Severe disease and death were associated with age and existing comorbidities. SARS’s natural history is poorly characterised, except for the incubation period and mean onset-to-hospitalisation.

Our associated R package, epireview, contains this database, which can continue to be updated to maintain a living review of SARS epidemiology and models, thus providing a key resource for informing response to future coronavirus outbreaks.

## Introduction

The COVID-19 pandemic re-emphasised the threat posed by coronaviruses to global health. Three coronaviruses to date, SARS-CoV-1, MERS-CoV, and SARS-CoV-2, have caused large disruptive epidemics affecting multiple countries (1–3). SARS-CoV-2 is now endemic in humans, as are four other coronaviruses (HCoV-229E, -NL63, -OC43, and -HKU1), jointly contributing to a large respiratory infection burden globally (4).

SARS-CoV-1 (or SARS-CoV) is the first documented coronavirus to have caused an acute epidemic in humans. Cases of atypical pneumonia were reported in November 2002 (5), with the World Health Organization (WHO) issuing a global alert in mid-March 2003 for Severe Acute Respiratory Syndrome (SARS). Later that month, a new airborne coronavirus, SARS-CoV-1, was identified as its likely cause (5). A large epidemic ensued, with major health and economic impacts (6), including over 8,400 reported cases (with ~20% among healthcare workers (HCWs)) across 32 countries up to mid-2003 (7–9). The epidemic was characterised by superspreading events, in which many infections occurred in a short timeframe in the same setting (e.g. hospitals or housing estates), and high fatality, with over 900 deaths reported globally (10,11). Although no pharmaceutical countermeasures were available at the time, the epidemic was contained within a few months using traditional control measures, including risk communication, contact tracing, isolation and quarantine (12,13). These proved effective, likely due to SARS’s natural history, namely limited transmission from asymptomatic and presymptomatic individuals (6).

In July 2003, the WHO declared the outbreak contained. A few sporadic cases, some due to laboratory infections, emerged after 2003-2004. Nonetheless, SARS-CoV-1 still constitutes a public health threat, currently on the WHO’s list of priority pathogens for research and development (15). Re-emergence of the virus from animals (bats being its main reservoir (16)) or from laboratories are concerning prospects. Indeed, there is still no effective treatment (17), and despite promising advances (18,19), there is no approved vaccine.

Many studies have investigated the epidemiology and transmission of SARS, leveraging high-quality data on cases analysed with modern infectious disease epidemiology and modelling approaches (20). Although multiple SARS systematic reviews have been published (21–24) (Supplementary Material (SM) appendix E), they mainly focused on narrow aspects of SARS epidemiology (e.g. serial intervals, incubation periods or intervention effectiveness), and their static nature means there is no up-to-date resource providing a live picture of the latest knowledge on SARS epidemiology and modelling. Our study aims to fill this gap: we systematically reviewed published peer-reviewed SARS models and key epidemiological parameters. The extracted data are available in a flexible database that can be updated as new information is produced.

Synthesising estimates of key epidemiological parameters is critical to support the response to future SARS outbreaks. Robust epidemiological parameter estimates are also key inputs to mathematical models, which will likely be integral to future outbreak responses. Early analyses of the COVID-19 pandemic relied on assumptions that the natural history of SARS-CoV-2 was similar to those of SARS-CoV-1 or MERS-CoV (25,26). Our database will provide vital information to support early modelling efforts for future epidemics of SARS and novel coronaviruses more broadly.

## Methods

We followed the Preferred Reporting Items for Systematic Reviews and Meta-Analyses (PRISMA) guidelines. We registered our study protocol with PROSPERO (International Prospective Register of Systematic Reviews, #CRD42023393345).

### Search strategy and selection criteria

We searched PubMed and Web of Science for studies published from database inception up to March 8, 2019. We repeated the search to include publications up to June 24, 2024. Results were imported into Covidence (2024) (27) and de-duplicated. Titles and abstracts, and then full texts were independently screened by two reviewers (selected from CM, TR, IR, MR, NIE, JS, PD, HJTU, TN, DPD, AC, SB), and conflicts were resolved by consensus. Non-peer-reviewed literature and non-English language studies were excluded. See Supplementary Material (SM) A.1 for further details on study selection, SM-Table A.1 for full inclusion and exclusion criteria and SM-section D for the PRISMA checklists.

### Data extraction

21% of full texts (n total=288) meeting the inclusion criteria were randomly selected and double-extracted to validate the extraction process. A consensus on discordant results was established, after which 20 reviewers (CM, TR, PD, KM, RJ, HJTU, TN, DPD, KP, BNCD, AV, KOD, PC, RJS, SIL, JTH, RM, RKN, AC, SB) independently conducted single extraction on the remaining full texts. Data on publication details, quality assessment (using a customised questionnaire, see Data analysis section and SM-A.5), transmission model details, basic and effective reproduction numbers, any epidemiological delays (e.g. incubation period or symptom-onset-to-hospitalisation or -outcome delays), case fatality ratios (CFRs), attack rates, growth rates, overdispersion, seroprevalence, and risk factors, were extracted using a Microsoft Access database (version 2305). For risk factors, we extracted only whether-or-not a factor was statistically significant, as reported in the articles, and if an analysis was adjusted for other covariates. This is because differences in reference groups and stratification made it unsuitable to compare other measures (e.g. odds ratios) across studies. We excluded systematic reviews from our study but used them to cross-check that all eligible studies were included (SM-section E). Full details of the data extraction process (SM-section A.2), database structure (SM-tables B.6-B.7) and extracted data (SM-tables B.8-B.13) are provided in SM.

Given there is a single documented SARS epidemic, we also analysed information on case and death numbers reported by country from the final report of the Hong Kong SARS Expert Committee (HKSEC) (10) to assess the global burden of the epidemic from a single source without redundancy.

### Data analysis

The risk of bias was assessed using a bespoke quality assessment (QA) questionnaire, with QA scores for each article calculated as the proportion of ‘Yes’ responses to applicable questions. A local polynomial regression was used to analyse temporal trends in QA scores (SM-Figure B.1). All manuscript figures show extracted parameters from “high QA” studies only, i.e. those with a QA score >50% (as in previous work (28)). Results from all studies are shown in SM Section B.5.1. For each parameter, we declare the total number of parameter extractions and studies and the number of high QA parameter extractions and studies.

Analyses were conducted in *R* (version 4.2.2) (29); curated data on outbreaks, models, and epidemiological parameters were added to the *epireview* R package (30) (SM-section C).

See SM section A.3.4 for how uncertainty is defined across all parameters.

Meta-analyses of published estimates were conducted using the *meta* R package (31) for the mean incubation period and the mean symptom-onset-to-hospital-admission delay. Common and random effects models were used to generate pooled delay estimates with 95% confidence interval (CI) and *I^2^* heterogeneity estimates (SM-A.3). We did not perform meta-analyses for other parameters due to insufficient estimates of central tendency paired with sample uncertainty.

In addition to our review, we used data from the final HKSEC report (10), including the number of SARS cases, hospitalisations, deaths, recoveries, imported cases by country, and the onset dates for the first and last probable cases. We used this dataset to identify countries with local transmission and calculate CFRs for each country. We used mixed-effect logistic regression to obtain a pooled CFR estimate (Figure 2, SM-A.3.2).

## Results

The search returned 28,356 potentially relevant articles. De-duplication retained 14,929 articles for title/abstract screening. 878 studies were retained for full-text screening, with 288 studies meeting the criteria for final inclusion (SM-Table A.1). As shown in Figure 1, the main reasons for study exclusion were “no reported parameters or models of interest” (n=220) and “no original estimates” (n=138). We report Cohen’s kappa for the screening and full-text review (SM-Figure B.13).

**Figure 1:**
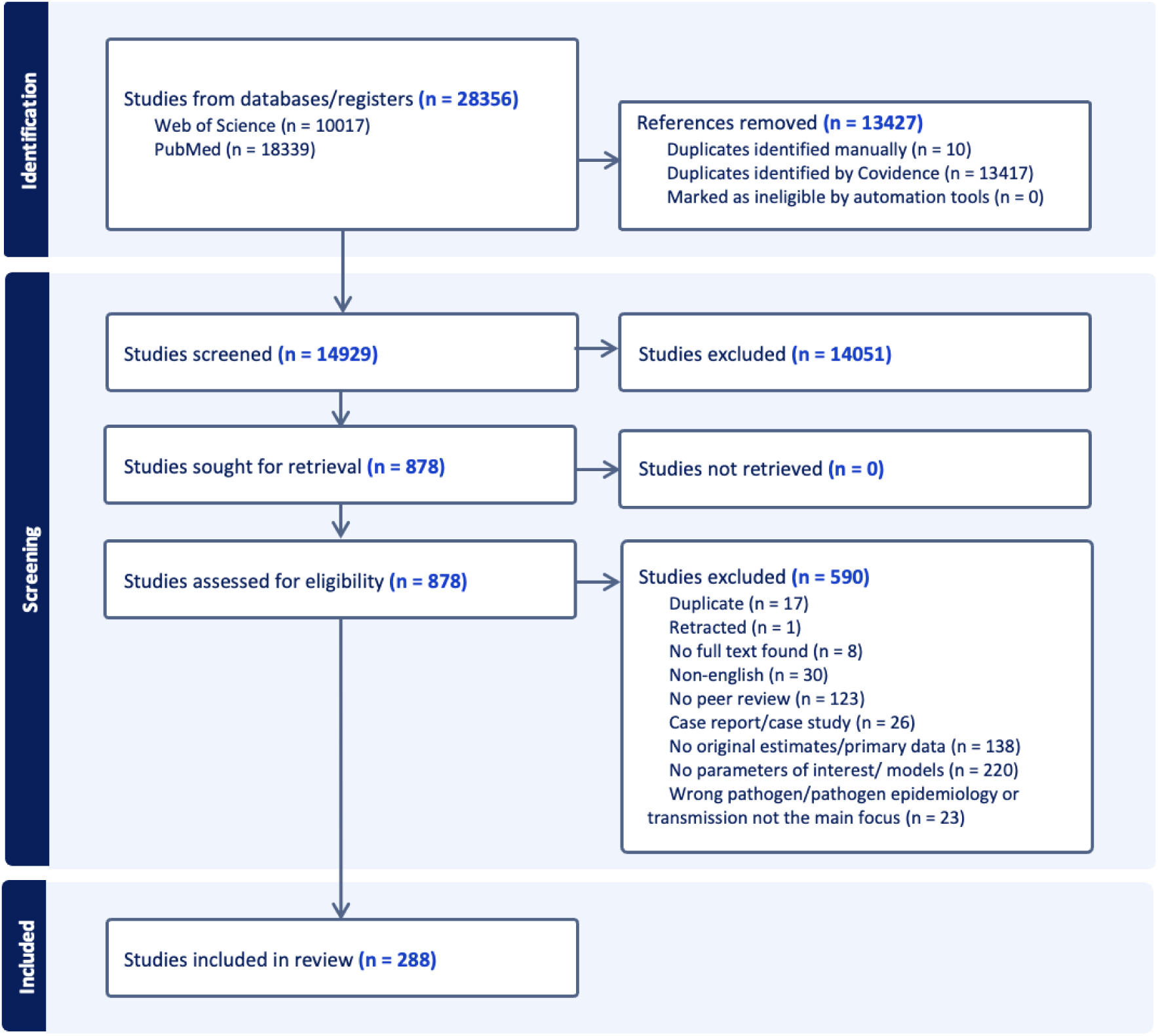
Study selection according to PRISMA guidelines and criteria described in SM-Table B.1. (Reasons for abstract exclusion not provided by Covidence).

We extracted 519 parameters and 243 risk factors from 186 articles (SM-Tables B.3-4, B.8-13 and SM-Figure B.2) and 112 models from 108 articles (SM-Table B.5 and SM-Figure B.3).

Figure 2 illustrates the global burden of the 2003-2004 SARS epidemic, as estimated from the HKSEC final report (10). China reported the most cases (5327), followed by Hong Kong (1755), Taiwan (665), Canada (251), Singapore (238), and Vietnam (63). Local transmission was not documented elsewhere, though cases were reported in other countries.

**Figure 2:**
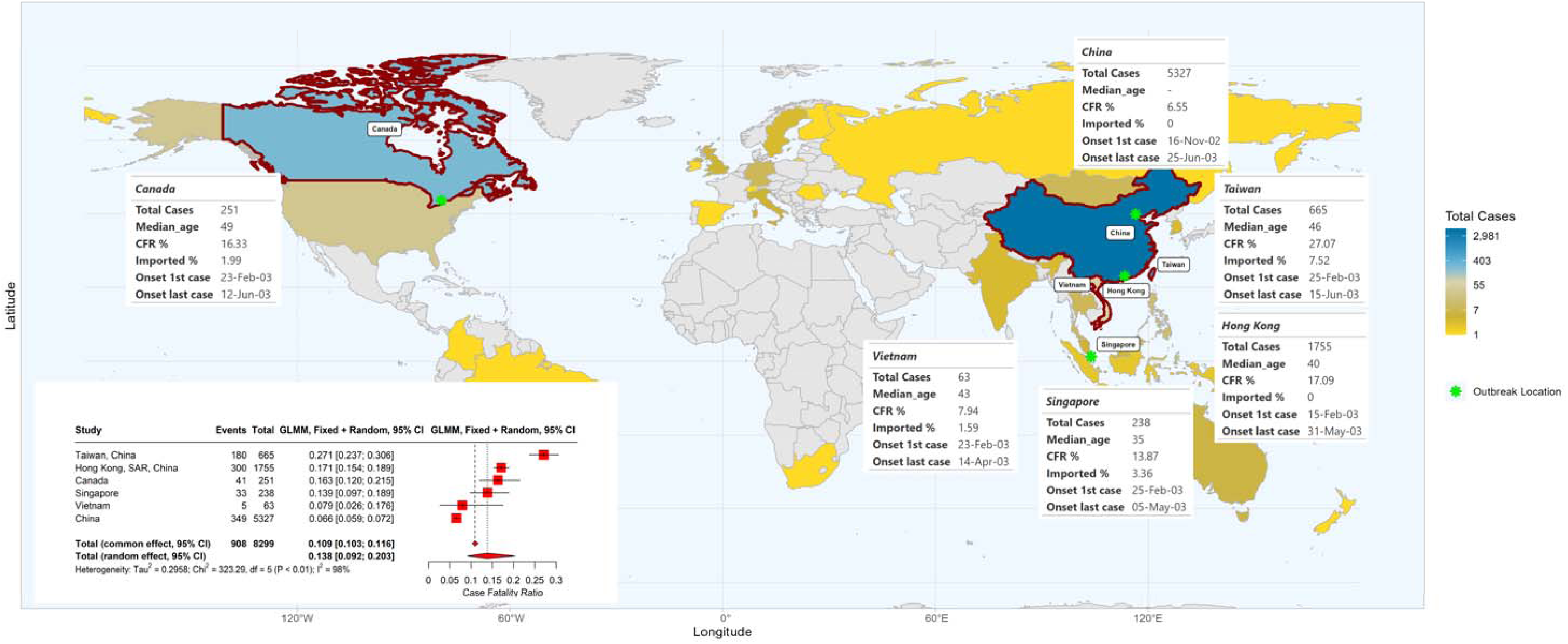
Overview of the 2003-2004 SARS epidemic, based on final report of the Hong Kong SARS Expert Committee (10). Coloured countries and special administrative regions are those which reported confirmed SARS cases. Inset tables are shown for all locations with reported local SARS-CoV-1 transmission, indicating total cases, median age (except for China, for which median age was not reported), CFR, percentage of imported cases and dates of symptom onset of first and last case. In the bottom left corner, we report the mixed-effect logistic regression estimates of adjusted CFRs for locations with local transmission: red squares indicate location-specific estimates. Red diamonds represent overall common effect estimates - in which all aggregated data are assumed to come from a single data-generating process with one common CFR, and overall random effect estimates - that allow the CFR to vary by location and accordingly give different weights to each location in the overall estimate (SM-A.3.2). The “Events” column indicates the reported number of deaths. GLMM=generalised linear mixed-effects model (SM-A.3.2).

We extracted 34 basic reproduction number (R_0_) estimates from 25 studies in total and 25 effective reproduction number (R_t_) estimates from 13 studies (SM-Tables B.4, B.8). R_0_ measures transmission without interventions and population immunity, whilst R_t_ captures transmissibility, including as influenced by immunity, control measures, and behavioural changes (32). Central R_0_ estimates from 13 high-QA studies ranged between 1.1 and 88.3 (n=16). We omit two outlying estimates from our main analysis of 88.3 (33) and 12.985 (34) due to their unique study contexts: one pertains to a nosocomial setting, and the other is from a study investigating the impact of priors on Bayesian inference. The remaining 14 R_0_ central estimates from 11 studies ranged from 1.1 to 4.59, with uncertainty intervals spanning 0.25-5.31 (Figure 3A). R_0_ estimates were similar across countries and the outbreak phase they were estimated from (Figure 3A and SM-B.4, B.8). As expected, R_t_ estimates tended to be lower. Of the 17 R_t_ estimates from 8 high-QA studies, 11 had a central value below or at the threshold R_t_=1, indicating a controlled epidemic, with the remaining six central R_t_ estimates ranging from 2.4 to 4.8 (Figure 3B). All R_t_ estimates from the start of the outbreak (n=5) were above 1, while four mid-outbreak, two end-outbreak, and two control-measure period estimates were below 1.

**Figure 3:**
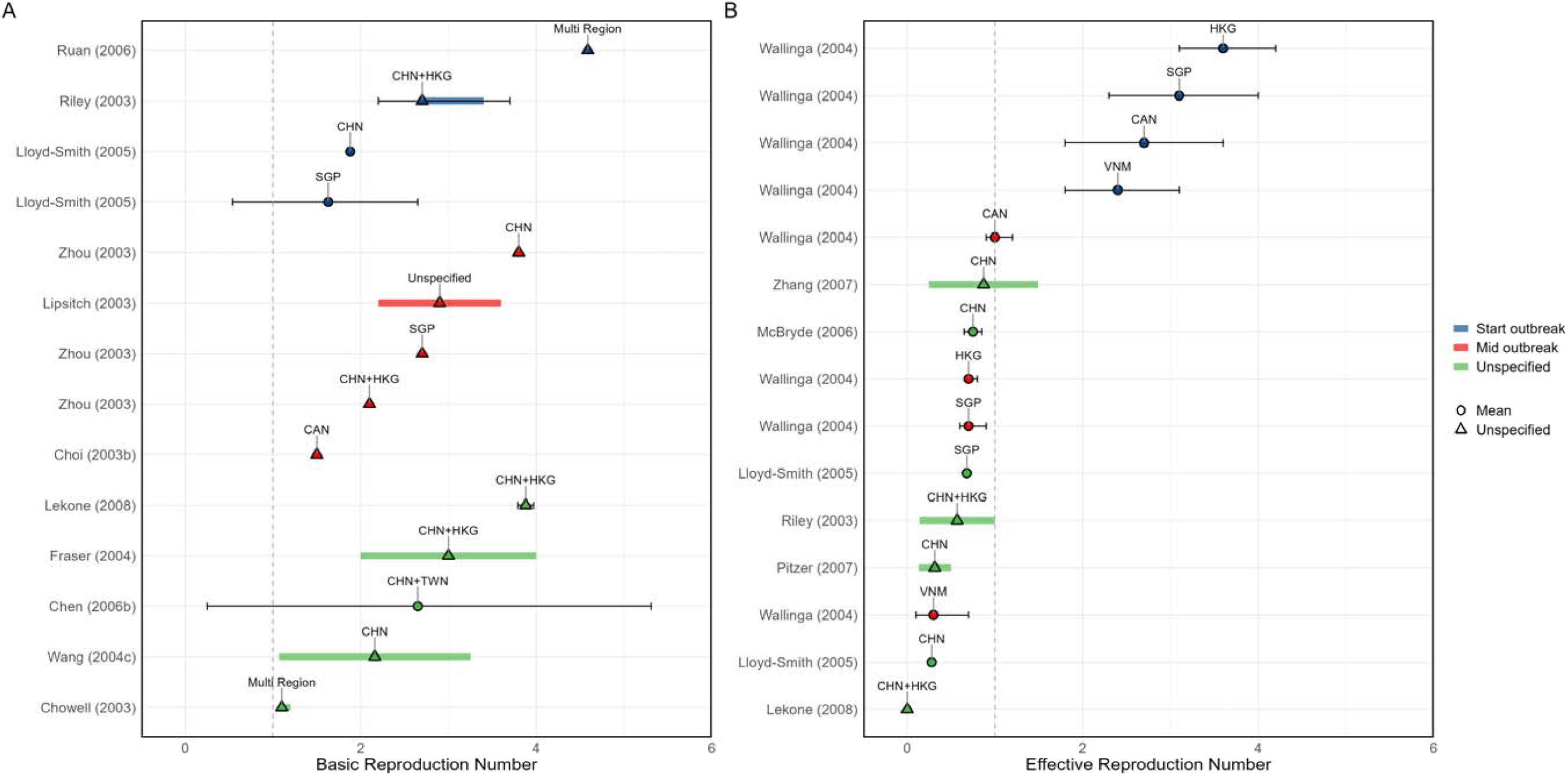
Overview of estimated SARS-CoV-1 (A) basic reproduction numbers (R_0_) and (B) effective reproduction numbers (R_t_). Circles represent mean estimates, and triangl s represent unspecified central estimates. Thin black solid lines represent uncertainty estimates, and solid shaded bars represent ranges of central estimates reported, e.g. when disaggregated by certain characteristics (e.g. age, sex, region, time) or using different estimation methods. Colours represent when during the outbreak the study was conducted, as extracted by reviewers. The vertical dashed line indicates the threshold value of 1. Estimates are labelled with the country of study. CHN = China, HKG = Hong Kong, SGP = Singapore, CAN = Canada, TWN = Taiwan, VNM = Vietnam. Outlying estimates from Kwok (2007) (33) and Moser (2015) (34) are not displayed. Only parameters from studies with a QA score > 0.5 are plotted.

Five growth-rate estimates were extracted from three studies, 33 attack rates (AR) from 22 studies, and nine secondary attack rates from eight studies (SM-Tables B.4, B.8).

Growth rate central estimates ranged from 4.22%, corresponding to the midpoint of the range of captured estimates in Zhang et al (35), to 16% per day (four estimates from two high-QA studies, all estimated in the presence of control measures, Figure 4C), corresponding to a doubling time between 4 and 16 days (SM-A.3).

**Figure 4:**
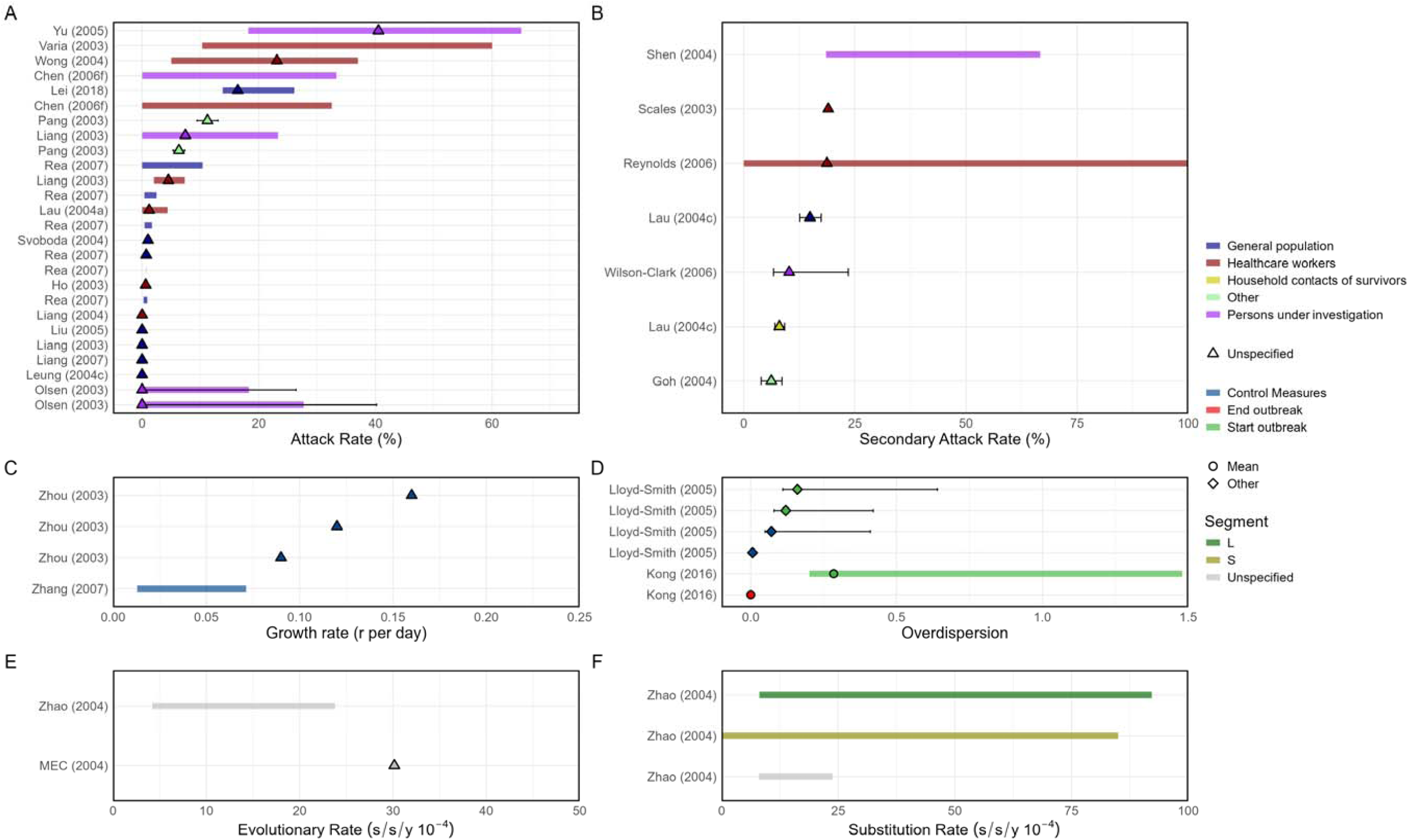
Overview of estimated SARS (A) attack rates, (B) secondary attack rates, (C) growth rates, (D) overdispersion, (E) evolutionary rates, (F) substitution rates. Circles represent mean estimates, and triangles represent unspecified central estimates. Thin solid lines represent uncertainty estimates, and solid shaded bars represent ranges of central estimates reported, e.g. when disaggregated by certain characteristics (e.g. age, sex, region, time) or using different estimation methods (e.g. compartmental, branching process models, etc). Colour represents (A & B) the study population considered, (C & D) when during the outbreak the study was conducted, with “control measures” referring to a time in the outbreak when interventions were reported to be in place, (E & F) long (L)/short (S) gene segment. In E and F, s/s/y refers to nucleotide substitutions per site per year. Only parameters from studies with a QA score > 0.5 are plotted.

AR estimates from the general population exclusively were generally low, with all but one central estimate of high-QA studies ranging from 0 to 5.18%, corresponding to the mid-point of the range of captured estimates of Rea et al (36) (n=11 from six studies) and one higher estimate at 16.4% (Figure 4A). ARs in HCWs were substantially higher, with central estimates ranging from 0.47 to 35.15% (n=7 from seven high-QA studies), as were ARs in persons under investigation (close contacts or targeted studies), with central estimates ranging from 0 to 40.5% (n=5 from four high-QA studies).

Secondary ARs showed a similar trend, with high estimates for HCWs (two central estimates from two high-QA studies ranging from 18.7 - 19%) and persons under investigation (two central estimates from two high-QA studies −10.2 and 42.6%%, corresponding to the mid-point of the range of captured estimates of Shen et al (37)), and lower estimates in other settings (three central estimates from two high-QA studies ranging from 6.2 to 14.9%, Figure 4B).

Overdispersion characterises heterogeneity in the number of secondary cases generated by one individual, with lower overdispersion estimates indicating more heterogeneity or superspreading. We extracted eight estimates of overdispersion from four studies (SM-B.8); seven estimates for the three high-QA studies had central estimates ranging from 7.52 * 10^-7^ to 0.285, with uncertainty extending as high as 1.48 (Figure 4D).

We extracted 12 parameters detailing genetic mutation from six studies in total. Filtering for only high QA studies, we extracted two mutation rate (evolutionary rate) estimates from two studies, one of which also reported three substitution rate estimates (Figure 4E-F, SM-Table B.8). Central estimates ranged from 14 to 30.15 x 10^-4^ substitutions per site per year (s/s/y) for the evolutionary rate and from 15.90 to 50.15 x 10^-4^ s/s/y for the substitution rate, where the central estimate corresponds to the mid-point of the range of captured estimates for each study. Uncertainty around those was not reported, but ranges of point estimates were provided, e.g. reflecting different fragments of the genome considered: one study reported central evolutionary rate estimates ranging from 4.2 to 23.4 x 10^-4^ s/s/y, and three studies reported wide ranges of central substitution rates estimates, from 0 to 92.20 x 10^-4^ s/s/y.

Seroprevalence estimates varied widely across studies, again depending on the population being investigated (SM-Table B12), with low seroprevalence in the general population and children and mixed groups outside hospital settings (8 seroprevalence estimates from 0 to 12.04% across five high-QA studies), moderate in HCWs (13 seroprevalence estimates from 0 to 88.9% across 11 high-QA studies), and high in persons under investigation (nine seroprevalence estimates from 0.19 to 100% across nine high-QA studies).

37 studies (31 high-QA) examined risk factors for SARS infection (SM-Table B.13, SM-Figure B.11-12). Most risk factors considered were found to be both non-significant and significant across different studies. Close or household contact with an infected individual and occupation were more frequently significantly (vs. non-significantly) associated with infection (SM-Figure B11 A). In contrast, age, sex and comorbidity were more likely to be non-significantly (vs. significantly) associated with infection.

We extracted 116 CFR estimates from 77 articles (86 estimates from 56 high-QA studies) (SM-Table B.4 and B.11, SM-Figure B.2). Half of these (n=57 estimates from 42 studies) were computed using a naïve approach (i.e. dividing the number of deaths by the number of cases), which leads to biased estimates if some cases have unknown final status. This can often be the case in real-time analyses (38). Many studies did not explicitly specify the CFR estimation method (n=37 estimates from 24 studies). Of those which corrected for right censoring (n=22 estimates from 14 studies), CFR estimates ranged from 0.1 to 30.8%. Many studies were for the same countries (e.g., nine CFR estimates from four studies for China); hence, estimates were likely not independent. We, therefore, also estimated the CFR retrospectively from data in the HKSEC final report (10); the CFR varied by country, from 6.6% (95% confidence interval (CI) 5.9-7.2%) in mainland China to 27.1% (95% CI 23.7-30.6%) in Taiwan, with a pooled random effect estimate of 13.8% (95% CI 9.2-20.3%) and a pooled common effect estimate of 10.9% (95% CI 10.3-11.6%, Figure 2).

Many studies considered potential risk factors for severe disease (21 analyses from eight studies, SM-Figure B.11 B, ten analyses from three high-QA studies) or death (82 analyses from 36 studies, SM-Figure B.11 C, 62 from 27 high-QA studies). Age, comorbidities and sex were the most studied risk factors for both severe disease and death (SM Figure B.10). Most analyses found age and comorbidities to be significant risk factors for severe disease and death. Most studies that considered sex as a risk factor for severe disease found it to be significant; there was mixed evidence of sex being a risk factor for death (14 studies significant versus 12 studies not significant). Contacts with infected individuals, in or outside the household, were not found to be a significant risk factor for severe disease (n = 1 analysis from one study), but close contact was identified as a significant risk factor for death in three out of four analyses from three studies. Occupation and hospitalisation were often identified as significant risk factors for death (10 analyses from eight studies and three analyses from two studies, respectively).

We extracted three estimates of mean serial interval (SI) from two studies and seven estimates of the infectious period from seven studies (Figure 5A-B and SM-Table C.10). The three SI estimates (all from high-QA studies) were broadly consistent, albeit uncertainty was large, encompassing values from 6.77 to 18.55 days. Four central estimates of the infectious period across four high-QA studies ranged from 4.84 to 10 days (uncertainty range 5.8-14.3 days; note not all studies report uncertainty), with a fifth high-QA study yielding a much higher central estimate of 21.6 days (uncertainty 14.9 to 26.8 days).

**Figure 5:**
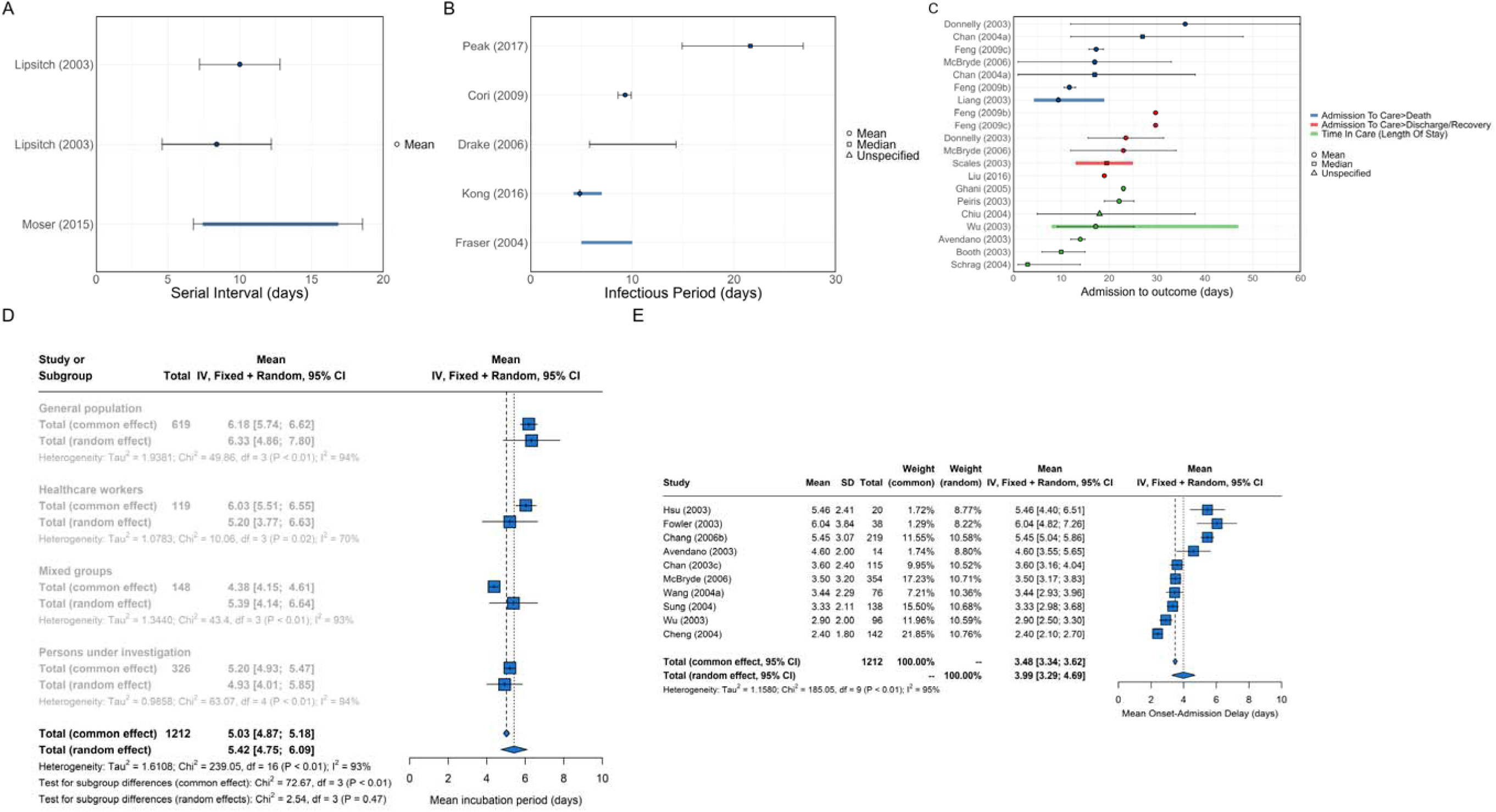
Overview of SARS epidemiological delays: estimates of (A) mean serial interval, (B) mean infectious period, (C) mean duration from hospital admission to final health outcome (i.e. death or recovery); and meta-analysis of epidemiological delays: estimates of (D) mean incubation period, (E) mean duration from symptom onset to hospital admission. In panels A, B, & C, circles represent mean estimates, squares represent median estimates, and triangles represent unspecified central estimates. Thin solid lines represent uncertainty estimates, and solid shaded bars represent the range of central estimates reported, e.g. when disaggregated by certain characteristics (e.g. age, sex, region, time) or using different estimation methods. In plot C, colour represents different final health outcomes. In D, blue squares indicate common effect and random effect estimates across different study populations, and blue diamonds represent: overall common effect estimates - in which all aggregated data are assumed to come from a single data-generating process, and overall random effect estimates. In E, blue squares represent study-specific estimates, and blue diamonds represent overall common effect and random effect estimates. Only parameters from studies with a QA score > 0.5 are plotted.

We extracted 49 incubation period estimates from 39 studies, with 42 estimates from 32 high-QA studies, including 17 estimates with sufficient information for inclusion in a meta-analysis (Figure 5D and SM-Table B.10). The pooled mean incubation period estimate was 5.03 days (95% CI 4.87-5.18) using a common effects (CE) model, and 5.42 days (95% CI 4.75-6.09) using a random effects (RE) model. The high I^2^ (93%) suggests substantial heterogeneity across studies. Meta-analyses restricted to specific study populations (general population, HCWs, mixed groups, and persons under investigation) highlighted small differences in the mean incubation period across populations, with high I^2^ throughout, indicating high heterogeneities between studies within each subgroup. The mean incubation period was shortest in persons under investigation (RE estimate 4.93 days, 95% CI 4.01-5.85) and longest in the general population (6.33, 95% CI 4.86-7.80).

We extracted 38 estimates of the delay from onset of symptoms to hospitalisation from 33 studies, including 29 estimates from 25 high-QA studies eligible for inclusion in meta-analysis (Figure 5E and SM-Table B.10). The estimated mean onset-to-hospitalisation delay was 3.48 days (95% CI 3.34-3.62 days) using the CE model and 3.99 days (95% CI 3.29-4.69 days) using the RE model, with a high I^2^ (95%) suggesting large heterogeneity across studies.

We extracted 21 estimates from14 studies characterising the delay from hospitalisation to outcome (i.e. death or recovery, Figure 5C and SM-Table B.10). Central estimates ranged from 9.4 to 35.9 days for the mean time from hospitalisation to death (seven estimates from six high-QA studies) and from 19 to 29.7 days for the mean time from hospitalisation to recovery (six estimates from six high-QA studies). We also extracted 15 estimates from 14 studies of mean time in hospital, with central estimates ranging from 3 to 23 days across seven high-QA studies.

We conducted our meta-analyses using only high-QA studies. None of the lower-QA studies provided sufficient information to be included in our meta-analyses; hence, these did not affect our results (SM-Figure B.6).

112 SARS transmission models were extracted from 108 studies (SM-Figure B.3, SM-Tables B.5, B.9). Compartmental models were the most frequent model type (n=63 from 62 studies), followed by individual-based models (n=9 from nine studies), and branching processes (n=7 from seven studies). Most compartmental models (n=43 of 63) were deterministic. Approximately half of the extracted models had been calibrated using observed data. Models captured a wide range of interventions, including quarantine and contact tracing, as well as behaviour changes (SM-Figure B.3(F)). No model had publicly available code.

## Discussion

In this systematic review, we compiled and analysed published epidemiological parameter estimates and mathematical models of SARS-CoV-1. SARS epidemiology has been well characterised overall, with 519 epidemiological parameters, 243 risk factors, and 112 mathematical models extracted in this review, covering multiple geographic regions affected by the 2003-2004 epidemic (Figure 2, SM-Figure B2). Synthesising this information in a central dynamic database, as we have done in the R package *epireview*, is critical to prepare for potential future outbreaks of SARS-CoV-1, a high-threat virus on the 2024 WHO list of priority pathogens (39). This central resource will also be useful for epidemic preparedness, to characterise the epidemiology of existing coronaviruses and anticipate the potential epidemiological profile of future ones.

SARS was first detected in China and then affected multiple other countries. Unlike with SARS-CoV-2, however, international spread was limited, with local transmission only identified in a handful of countries.

However, SARS transmissibility in the affected regions was high, with growth rate estimates translating into a doubling of cases every 4 to 16 days (Figure 4 growth rates converted to doubling times (SI A.3)), and R_0_ estimates broadly ranging from 1.1-4.59 (Figure 3A). These R_0_ estimates were obtained using varied statistical approaches and data types. They were reported in very heterogeneous formats, sometimes without any characterisation of uncertainty, making comparison and synthesis across studies challenging. These R_0_ estimates are comparable to those for the SARS-CoV-2 wildtype (~2.5) (40) and generally slightly higher than for pandemic influenza (typically 1-2.5) (41–43). Detection of the SARS epidemic and the subsequent WHO global alert rapidly prompted interventions, including quarantine, isolation, strict hygiene measures in hospitals and social distancing (13,44). These eventually led to a decline in cases, consistent with a reduction of R_t_: estimates at the start of the epidemic broadly align with the basic reproduction number, ranging from 2-4 (Figure 3B), whilst estimates later in the epidemic suggest interventions were effective, bringing R_t_ below 1.

However, transmission was highly heterogeneous: overdispersion estimates suggest significant variations in the number of secondary infections generated by each case. Following the approach of Lloyd-Smith et al. (45), the range of central overdispersion estimates (between 7.52 * 10^-7^ (post interventions) and 0.285 (pre interventions), Figure 4) suggests that approximately 91% of SARS-CoV-1 transmission can be attributed to the 20% most infectious individuals in the pre-intervention period but this increases to 99% in the post-intervention period (SI A.3.3). Hence, superspreading in SARS-CoV-1 is more prominent than for many other viruses (45), including SARS-CoV-2 (46). This characteristic makes SARS particularly threatening, as higher levels of superspreading make epidemics harder to control.

Attack rate, secondary attack rate, and seroprevalence estimates extracted in this review show that SARS-CoV-1 transmissibility highly depended on the sub-population considered, as observed during the COVID-19 pandemic (47). General population studies reported SARS-CoV-1 attack rates and seroprevalence generally under 1% (Figure 4A and SM-Table B.12), whilst estimates among HCWs were substantially higher, with central estimates ranging from 1.2% to 35.15%.

We extracted risk factors associated with infection (SM-Figure B.11). Unsurprisingly, close contact with confirmed cases was frequently identified as a significant risk factor for infection. Occupation, including HCWs, was also often reported as a significant risk factor for infection, in line with higher attack rates, secondary attack rates, and seroprevalence estimated among HCWs.

Published estimates of CFR suggest high severity, with >10% of cases fatal (SM-Table B.11 and Figure 2). This is one order of magnitude higher than SARS-CoV-2 (CFR up to ~2% for the most severe variants (48) (40)) and pandemic influenza (CFR up to ~1-3% for the 1918 influenza pandemic (49)). Such high severity likely aided case identification and intervention targeting. However, combined with a relatively high transmissibility and high superspreading, it emphasises the threat posed by SARS-CoV-1 and potential future coronaviruses that could share these characteristics.

Risk factors for hospitalisation and death were aligned with those previously identified for respiratory viruses, including age and comorbidities. Sex demonstrated mixed results, with studies equally identifying it as a significant and non-significant risk factor for SARS mortality. For COVID-19, the CFR is higher for men than for women (50). Being an HCW was frequently identified as being significantly associated with the risk of death. However, these results should be interpreted with caution as we did not extract the direction of the association.

The natural history of SARS was less well-characterised. We identified only three estimates for the central estimate of the SI, all with considerable uncertainty (Figure 5A) and one estimate for the generation time. The few estimates for the infectious period suggested a duration of around a week. However, some uncertainty and an outlier again suggested a much longer infectious period of several weeks (Figure 5B). The mean incubation period was better characterised, enabling a meta-analysis yielding a pooled RE model estimate of 5.42 days (95% CI 4.75-6.09, Figure 5D), similar to estimates for SARS-CoV-2 (51). Subgroup analyses suggested variations in mean incubation periods across populations, although this may reflect differences in study designs and associated biases. Although not a focus of our systematic review, very little evidence was found to suggest a role of asymptomatic transmission for SARS. This suggests that most SARS-CoV-1 infectious individuals were symptomatic, and their infectiousness started after symptoms, meaning that the latent period would be at least as long as the incubation period (52,53). Little or no asymptomatic transmission enables containment, as infectious individuals can be effectively isolated following symptom onset. This contrasts with SARS-CoV-2, where asymptomatic transmission has been widely documented, limiting the effectiveness of control measures that rely on symptom-based case identification and isolation (54).

Delay from symptom onset to hospitalisation is also an important marker of epidemic management, with shorter delays characterising epidemics where cases are isolated more promptly and which are more likely to be controlled. Our meta-analysis estimated a relatively short mean time from symptoms to hospitalisation of 3.99 days (95% CI 3.29-4.69) for the RE model. However, there were substantial variations between studies, with no clear indications of what factors may drive such heterogeneity (Figure 5E, SM-Figure B.8), except that the delay reduced over time due to improved population awareness (55). Further studies into drivers of delayed hospitalisation may help to control outbreaks of SARS-CoV-1 and other coronaviruses. Estimates of mean time in hospital were also highly variable between studies, even when considering only individuals who die or only individuals who recover (Figure 5C). Similarly, further investigation of drivers of such heterogeneity would help adequately prepare increased healthcare capacity for future coronavirus outbreaks.

We identified only two high-QA studies reporting estimates of the substitution and evolutionary rates for SARS-CoV-1, with widely ranging estimates for different genome sections. The paucity of estimates is unsurprising given that the SARS epidemic happened early in the 21^st^ century when genetic sequencing was not yet commonly utilised. The higher end of those ranges, with evolutionary rates up to 8*10^-3^ substitutions per site per year, emphasises the potential for SARS and similar coronaviruses to evolve rapidly, possibly even faster than SARS-CoV-2 (56).

SARS modelling studies have been continuously published since 2003. Most modelling studies contemporary with the 2003-2004 epidemic sought to infer epidemiological parameters and forecast epidemic trends based on data. In contrast, more recent modelling studies have predominantly been theoretical, using SARS as a case study to explore methodological questions. While the QA scores of non-modelling studies remained broadly constant, we noted a decline in QA scores of modelling studies over time (SM-Figure B1 (C)), possibly reflecting this shifting focus.

None of the modelling studies provided associated model code, in line with recent estimates that only 0.5% of medical studies to date provide public-access code (57). The absence of code also likely reflects a less widespread practice of open-source code in 2003 compared to 2024, with journals and funders requiring open-source code only recently (58).

Our work has limitations. Firstly, we excluded non-peer-reviewed literature and studies not in English from our review. Restriction to peer-reviewed papers ensures our review adheres to high-quality standards. The language restriction means that we may have excluded relevant studies, particularly those in Chinese (SM-Table F.17). Secondly, to keep this review manageable given the volume of peer-reviewed studies on SARS-CoV-2, we added a “SARS-CoV-2” exclusion term to our search criteria. Therefore, studies comparing SARS-CoV-1 and SARS-CoV-2 will have been omitted. Overall, given that the literature on SARS is based on a single well-documented epidemic, it is unlikely that these restrictions left out substantial pieces of knowledge not otherwise covered. Moreover, further relevant literature not covered here could, in the future, be added to our ‘epireview’ R package, where all information extracted in this review is already publicly available (30).

When SARS-CoV-2 was first identified, initial assessments of potential epidemic impact and intervention options were informed by epidemiological and modelling analyses which, in the absence of characterisation of the new coronavirus, assumed that its natural history and epidemiology (e.g. degree of superspreading) would be similar to those of SARS-CoV-1 and MERS-CoV (25,26,59–61). Similar assumptions may need to be made in future outbreaks. Our systematic review and dynamic database will provide a critical resource to support the timely development and robust parameterisation of mathematical models in future epidemics of SARS and novel coronaviruses.

## Declarations

### Availability of data and materials

https://github.com/mrc-ide/epireview/tree/main/data

### Code availability

https://github.com/mrc-ide/epireview; https://github.com/mrc-ide/priority-pathogens

### PROSPERO

CRD42023393345 (https://www.crd.york.ac.uk/prospero/display_record.php?RecordID&RecordID=393345)

### Authors’ contributions

SB, SvE, AC, and NI-E conceptualised this systematic review. CM, TR, IR, MR, NIE, JS, HJTU, AC, and SB searched the literature and screened the titles and abstracts. CM, TR, IR, MR, JS, PD, HJTU, TN, DPD, and AC reviewed all full-text articles. CM, TR, PD, KM, RJ, HJTU, TN, DPD, KP, BNCD, AV, KOD, PC, RJS, SIL, JTH, RM, RKN, AC, and SB extracted the data. CM, TR, AC and SB formally analysed, visualised, and validated the data. CM, TR, TN, CNS, and SB were responsible for software infrastructure. AC acquired funding. CM, TR, and AC were responsible for project administration. GC-D, HJTU, and RKN were responsible for training individuals on accessing Covidence and designing the Access system. CM and AC supervised the systematic review. CM, TR, AC, and SB wrote the original manuscript draft. All authors were responsible for the methodology and review and editing of the manuscript. All authors debated, discussed, edited, and approved the final version of the manuscript. All authors had final responsibility for the decision to submit the manuscript for publication.

## Data Availability

The code and data availability is available at https://github.com/mrc-ide/epireview and https://github.com/mrc-ide/priority-pathogens

https://github.com/mrc-ide/priority-pathogens

https://mrc-ide.github.io/priority-pathogens/articles/pathogen_sars.html

