## Supplementary Information for "Severe acute respiratory syndrome (SARS) mathematical models and disease parameters: a systematic review and meta-analysis"

### Contents

**A Additional Methods** **4**
A.1 Study Selection 4
A.2 Data Extraction 4
A.3 Data Analysis 5
A.3.1 Meta-analysis 5
A.3.2 CFR data synthesis 6
A.3.3 Overdispersion data analysis 6
A.3.4 Reporting Uncertainty 6
A.4 Inclusion & Exclusion Criteria 8
A.5 Quality Assessment 8

**B Additional Results** **9**
B.1 Quality Assessment Results 9
B.2 Parameter and Model Overview 10
B.3 Extraction Fields 13
B.3.1 Model extraction fields 13
B.3.2 Parameter extraction fields 13
B.4 Extracted Data 15
B.4.1 Transmission parameters 15
B.4.2 Models 23
B.4.3 Natural history 25
B.4.4 CFR 35
B.4.5 Seroprevalence 41
B.4.6 Risk factors 44
B.5 Additional Figures 54
B.5.1 Main text figures without quality assessment 54
B.5.2 Alternate variable grouping 57
B.5.3 Individual study meta-analysis 59
B.5.4 Risk factor plots 60
B.6 Screening statistics 62

**C The `epireview` package** **64**

**D PRISMA 2020 Checklists** **65**

**E Other Systematic Reviews** **68**

**F Excluded Studies** **69**

**G Pathogen Epidemiology Review Group (PERG) membership** **70**

### Overview

This supplement provides additional details of the methodology used in our literature review (section A) as well as additional results (section B), including full details of the information extracted as part of the review and synthesised in the main text. Section C introduces our R package `epireview`, where all the data gathered in this review is stored. Section D contains our PRISMA checklists. Section E details other SARS systematic reviews identified in our initial literature search and which we used to ensure no relevant study was omitted in our review. Section F lists all the studies we excluded. Finally, section G lists all members of the Pathogen Epidemiology Review Group who contributed to this work.

### A Additional Methods

This systematic review and meta-analysis of severe acute respiratory syndrome coronavirus 1 (SARS-CoV-1) is part of a series of systematic reviews of nine priority pathogens on the WHO's R&D blueprint list, which was registered on PROSPERO. Other systematic reviews in this series completed to date concerned Marburg virus disease [1], Ebola virus disease [2], and Lassa fever [3].

The code to reproduce our results is available at `https://github.com/mrc-ide/priority-pathogens`.

#### A.1 Study Selection

Original research papers in English were included if reporting on SARS-CoV-1 transmission, evolution, natural history, severity, seroprevalence, size of previous outbreaks or published mathematical transmission models. Non-peer-reviewed literature was excluded. Papers identified in the search were imported into *Covidence*, a software program used to manage systematic reviews. From a team of seven reviewers, two independent reviewers first screened titles and abstracts, followed by full-text reviews to assess eligibility for data extraction. Disagreements in eligibility determination were resolved by consensus between the two independent reviewers.

**Search term:** `(SARS OR SARS-CoV-1 OR "Severe acute respiratory syndrome") AND ((transmissi* OR epidemiolog*) OR (model* NOT imag*) OR (severity OR "case fatality ratio*" OR CFR OR "case fatality rate*" OR "mortality rate*" OR "attack rate*") OR ("infectious period*" OR "serial interval*" OR "incubation period*" OR "generation time*" OR "generation interval*" OR "latent period*" OR latency) OR (heterogeneit* OR superspread* OR "super spread*" OR super-spread* OR overdispersion OR overdispersed OR over-dispersion OR over-dispersed OR "over dispersion" OR "over dispersed") OR (infectivity OR infectiousness OR "growth rate*" OR "reproduction number*" OR "reproductive number*" OR R0 OR "reproduction ratio*" OR "reproductive rate*") OR ("pre-existing immunity" OR serological OR serology OR serosurvey*) OR (evolution* OR mutation* OR substitution*) OR (outbreak* OR cluster* OR epidemic*) OR ("risk factor*")) NOT (COVID-19 OR SARS-CoV-2)`

This search term is common to all pathogen systematic reviews in the series (apart from the first field containing the pathogen name). In addition, for this review, we added the final term above to exclude SARS-CoV-2 related papers, as these were not in scope for this review. The search was initially conducted on 08/03/19 but repeated on 18/11/2023 and 24/06/2024.

#### A.2 Data Extraction

##### Models

Details of disease transmission models were extracted, including model type, whether it modelled deterministic or stochastic processes, which transmission pathways were included, and in the case of compartmental models, the compartmental structure of human health states (e.g., SIR, SEIR). Furthermore, we extracted information on underlying model assumptions, whether the model is theoretical or fitted to data, which interventions were considered, and the availability of model code. See Table B.6 for full details.

**Parameters**

For each parameter, we extracted all available information, including types of value estimated (e.g., mean, median, standard deviation), uncertainty intervals (capturing the precision of estimates), and ranges (if, for example, multiple estimates were obtained from different populations or using different methods). Study context for parameter values included survey location and dates, sample size, basic demographic information, and timing of the survey in relation to reported outbreaks. See Table B.7 for full details.

For genomic data, if specified we noted the gene studied and if newly sequenced data were available. For reproduction numbers, we recorded the methods used for estimation (e.g. renewal equations, compartmental models, empirical methods). For case fatality ratios, we extracted whether the estimation approach accounted for cases with unknown final status or not [4]. For both case fatality ratios and seroprevalence, we additionally recorded numerators and denominators where available.

For risk factors, we extracted the outcome (e.g. infection or death), the risk factor for that outcome (e.g. age, sex or occupation), the type of occupation if specified, and whether the risk factor(s) estimates were statistically significant and/or adjusted. We chose not to extract numerical odds ratio estimates because studies may have used different stratifications or reference groups, making it challenging to compare values across studies. The information we extract offers an overview of risk factors explored across studies that may affect the risk of infection and death, which may be useful to consider when designing SARS transmission models.

For papers which reported multiple parameter values for the same parameter (e.g. for multiple locations, time periods, assumptions, sub-groups), we applied the 'rule of 5', that is, if *more than* 5 parameter values were reported, we only captured the range of values reported for this parameter. The threshold of 5 was chosen so that we can capture the location with community transmission of SARS-CoV-1.

To summarise studies, which only have ranges captured in the database, we report the midpoint of the range and state this explicitly.

#### A.3 Data Analysis

We compute doubling times as $T_d = \frac{\ln 2}{r}$, where $r$ is the reported growth rate estimate.

##### A.3.1 Meta-analysis

The meta-analysis of mean incubation periods and the mean onset-admission delay (in Figure 5 of the main text) followed a standard approach and was conducted using the `meta` R package [5].

A mixed-effects model is a linear model $y_i = \beta_0 + \sum_{j=1} \beta_j x_{ij} + u_i + \epsilon_i$, where $y_i$ are the observed data, $x_{ij}$ are explanatory variables, $\beta_j$ are fixed effect coefficients, $u_i$ are the random effects (centred around zero and independent across $i$), and $\epsilon_i$ are error terms. Meta-analysis is a special case of the above mixed-effects model with only an intercept term $\beta_0$ (a fixed-effects/common-effects model) or only an intercept term $\beta_0$ and a random-effects term $u_i$ associated with that intercept (a random-effects model).

In Figure 5D of the main text, we present our meta-analysis of the mean incubation period. The meta-analysis reports the common and random effects for each sub-group of the study population to investigate any patterns between groups (the subgroup analysis was done independently for each group).

The mean onset-admission delay meta-analysis in Figure 5E requires paired mean and standard deviation estimates of the onset-to-admission delay (the same applies to the mean incubation period meta-analysis). However, the underlying studies report several different parameter value types (e.g., means, medians, standard deviations, interquartile range, minimum/maximum), we use the functionality of *metamean*, which is implemented in the `meta` package, to infer the means and standard deviations for each study. In particular, we use the method provided by Cai et al. (2021) [6], which allows for unknown non-normal distributions.

*Data inclusion protocol for meta-analysis*

- The data is taken as extracted (post-data cleaning), and the mean and the standard deviations are inferred using the stated methods above (where possible).
- Any filtering (for sub-group, setting, country) will be stated clearly in the analysis.
- Parameter estimates, which are ranges as the 'rule of 5' was applied, were not included in the meta-analysis.
- The included studies in the mean onset-to-admission delay subgroup meta-analysis are available in Figure B.10 and for the mean incubation period meta-analysis in the main text Figure 5.

##### A.3.2 CFR data synthesis

We estimated location-specific CFRs using data from the the final report of the Hong Kong SARS Expert Committee [7]. Specifically, for each location $i$, the $CFR_i$ was calculated as the number of deaths divided by the total number of cases, computed as: cases who recovered + cases who died + cases still in hospital.

To obtain an overall estimate of the CFR using the same data, we used a generalised logistic mixed-effects model, in which the location-specific CFR estimates are transformed using the logit-transformation $y_i = \log\left(\frac{CFR_i}{1-CFR_i}\right)$ to ensure that the distribution was approximately normal, and then a generalised linear mixed-effects model is applied to the transformed CFRs to estimate the pooled effect. A comprehensive overview of the methodology is provided by Harrer et al. (2021) [8].

##### A.3.3 Overdispersion data analysis

We calculated the proportion of SARS-CoV-1 transmission events attributable to the 20% most infectious cases following the approach outlined in Lloyd-Smith et al. [9]. We take the basis reproduction number $R_0$ and the overdispersion parameter estimate $k$ from the literature [9], which parameterise the gamma distribution of the individual reproduction number $\nu$ with pdf $f_\nu(x)$ and cdf $F_\nu(x)$. The cdf for the transmission of the disease is given by:

$$F_{trans}(x) = \frac{1}{R_0}\int_0^x u f_\nu(u)du$$

This is the expected proportion of all transmissions due to infectious individuals with $\nu < x$.

The expected proportion of transmission events due to the 20% most infectious individuals was calculated as 1-$F_{trans}(x_{20})$, where $x_{20}$ is obtained by solving $1 - F_\nu(x_{20}) = 0.2$.

We used the following values for the computation:

| Location | Period | $R_0$ or $R_t$ | k | proportion of infections due to 20% most infectious |
|---|---|---|---|---|
| Singapore | pre-intervention | 1.63 | 0.16 | 0.912 |
| Singapore | post-intervention | 0.68 | 0.071 | 0.971 |
| Beijing | pre-intervention | 0.94 | 0.17 | 0.920 |
| Beijing | post-intervention | 0.28 | 0.0062 | 0.998 |

Assumed reproduction number values and overdispersion parameters $k$. For the pre-intervention period $R_0$ is used, for the post-intervention period $R_t$ for the applicable time period is used. All estimates are taken $R_0$, $R_t$ and $k$ estimates are from [9]

##### A.3.4 Reporting Uncertainty

A wide assortment of uncertainty formats and range reporting was extracted across all studies. In this section we clarify our wording when referring to uncertainty.

While most studies reported a central estimate for parameters of interest, some instead reported a range of values or we applied the 'rule of 5' as outlined in section A.2. These range of values are depicted throughout our manuscript figures via shaded colour bars in all figures. These ranges are included when we refer to "central estimates", and we specifically extract the midpoint of these ranges for plotting central estimate points.

This is different to where papers report uncertainty. We extracted both single type uncertainty estimates (e.g. standard deviation, or variance) as well as paired-type uncertainty estimates (e.g. confidence/credibility intervals). Uncertainty is depicted across all our figures using solid line error bars. Throughout the manuscript, when we refer to the "ranges of uncertainty" for a parameter, we refer to the lowest point across all uncertainty intervals to the highest point across all uncertainty intervals. For example, if three estimates for $R_0$ were given: 1.3 (95% CI 0.7-1.5), 1.4 (95% CI 1.1-1.7) and 1.5 (95% CI 1.4-1.6), we say that the ranges of uncertainty for the parameter, were 0.7 to 1.7.

Note, that we included fields to explicitly extract when reported standard deviations were of a sample, or of an estimator. However, only one extracted parameter was specifically highlighted as for an estimator, the majority were undefined.

Note, where standard errors were reported, we did not convert these to uncertainty estimates, and did not plot standard errors in our figures.

#### A.4 Inclusion & Exclusion Criteria

| Inclusion | Exclusion |
|---|---|
| Measures/estimates of human: Reproduction numbers (R, $R_0$, $R_t$, r, $R_e$), growth rate (r), doubling times, generation time, serial interval, incubation/latent period, case fatality ratio (CFR), attack rate, mutation rate (e.g. from phylogenetic study), overdispersion, risk factors (risk and the measure). | Non-English language publication |
| Mention of historical or any outbreak in humans: size, year, location, duration, spatial scale | Studies of co-infections. (local, regional, national, international). |
| Measures/estimates of animal: R, $R_0$, $R_t$, r, $R_e$, growth rate, mutation rate. | Animal studies that do not report R, $R_t$ etc. |
| Mathematical or statistical model of transmission. | Qualitative studies, e.g., KAP studies. |
| Measures of seroprevalence and negative seroprevalence in humans. | Pathogen not the primary focus of study. |
| Relative ratio of human-human vs animal introductions. | Duplicates. |
| Reviews that report inclusion criteria for reference checking. | Does not match any of the inclusion criteria. |
| For "small" pathogens[1], include case reports to potentially reconstruct serial interval distribution etc. | In-vitro studies. |
| | Non-peer reviewed publications, conference proceedings, abstracts, posters, letters to the editor |

Table A.1: Inclusion and exclusion criteria for papers.

#### A.5 Quality Assessment

| Theme | Question |
|---|---|
| Is the methodological/statistical approach suitable? (how the data are used) | 1. Clear and reproducible<br>2. Robust and appropriate for the aim [subjective criteria] |
| Are the assumptions appropriate? (input parameters/assumptions - what goes into the methodology) | 3. Clear and reproducible<br>4. Justified (published study or analysis of data)[objective criteria] |
| Are the data appropriate for the selected methodological approach? | 5. Clearly described and reproducible<br>6. Are issues in the data clearly discussed and acknowledged?<br>7. Are issues in the data accounted for in the chosen methodological approach? |

Table A.2: Quality assessment questionnaire: possible responses for each question listed were: Yes, No, and NA (non-applicable).

[1]The inclusion/exclusion criteria are reported as stated in the PROSPERO registration for the overall priority pathogen project. The threshold is determined on a case-by-case basis for each pathogen and SARS is not considered a "small" pathogen.

### B Additional Results

#### B.1 Quality Assessment Results

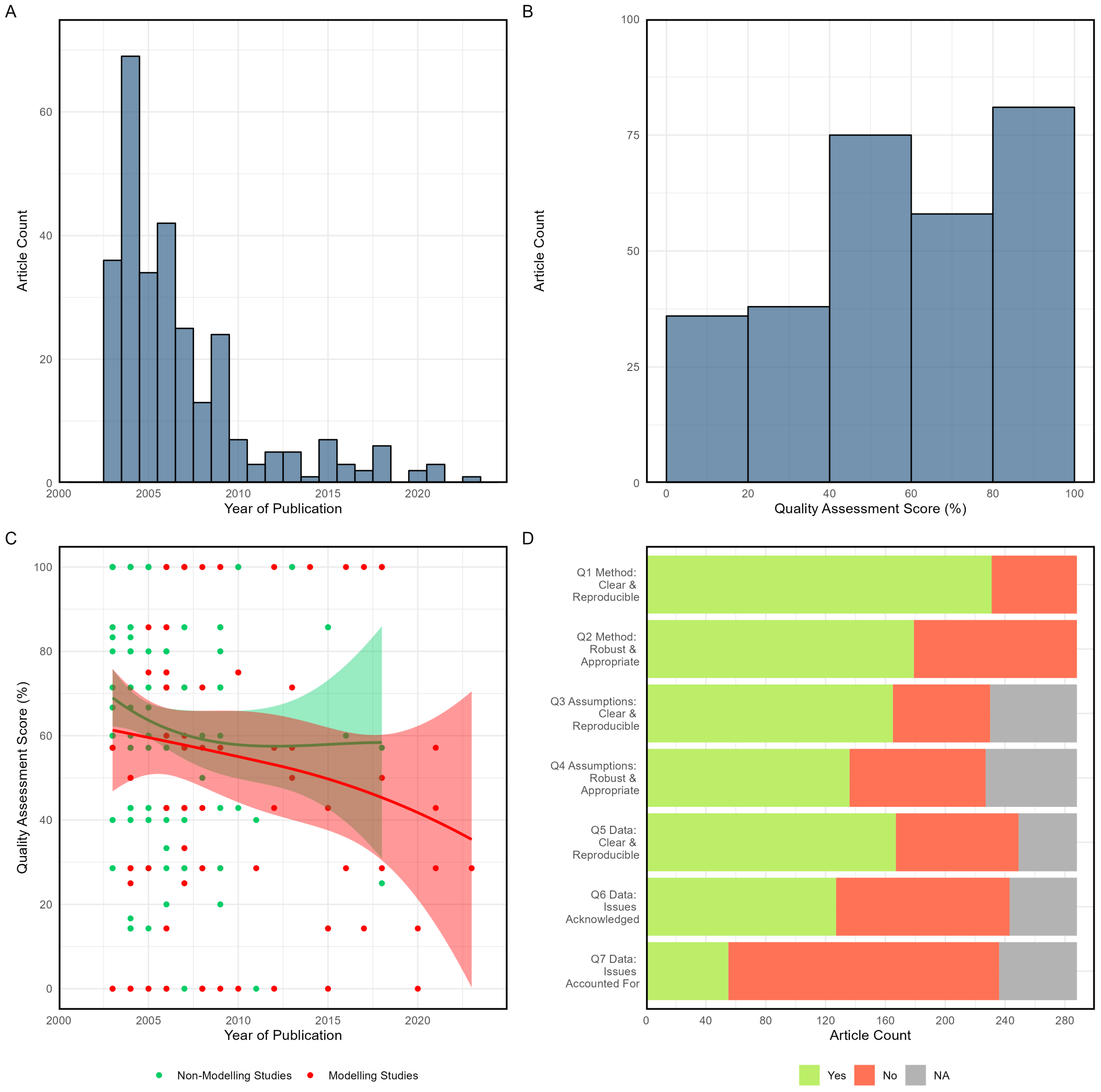

Figure B.1: (A) Article count by year of publication; (B) article count by quality assessment score (defined as percentage of 'Yes' answers relative to sum of 'Yes' and 'No' answers for each paper, removing 'NAs'); (C) quality assessment score by year of publication (time trends for articles with and without transmission models fitted via local polynomial regression); (D) article count for each quality assessment question scoring 'Yes', 'No' or 'Non-Applicable'.

#### B.2 Parameter and Model Overview

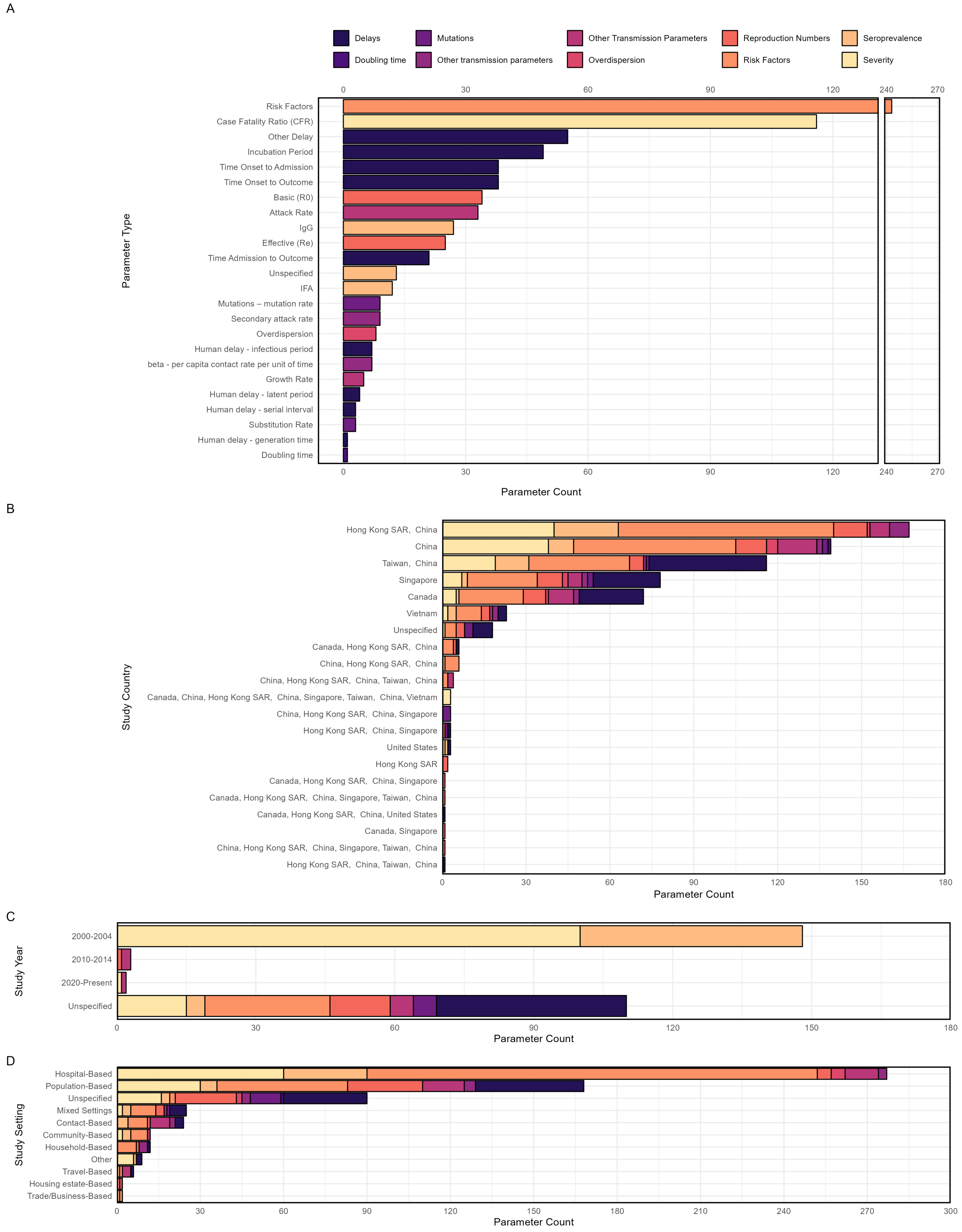

Figure B.2: Parameter type count by (A) parameter class, (B) survey country/countries, (C) survey decade (D) study setting. Colour corresponds to parameter type.

| Parameter class | **Total (no quality assessment)** | | **Quality assessment** > 0.5 | |
|---|---|---|---|---|
| | # parameters | # papers | # parameters | # papers |
| Attack rate | 33 | 22 | 26 | 16 |
| Doubling time | 1 | 1 | 0 | 0 |
| Growth rate | 5 | 3 | 4 | 2 |
| Delays | 217 | 100 | 155 | 74 |
| Mutation rates | 12 | 6 | 5 | 2 |
| Overdispersion | 8 | 4 | 7 | 3 |
| Reproduction number | 59 | 32 | 33 | 17 |
| Risk factors | 243 | 102 | 180 | 75 |
| Seroprevalence | 52 | 39 | 40 | 31 |
| Severity | 116 | 77 | 86 | 56 |
| Other transmission parameters | 16 | 14 | 14 | 12 |

Table B.3: Total parameters extracted and total number of papers these parameters were extracted from. Middle column includes all parameters. Right column only includes those papers that scored higher than 0.5 for quality assessment (see Table A.2).

| Parameter type | **Total (irrespective of QA)** | | **Quality assessment** > 0.5 | |
|---|---|---|---|---|
| | # parameters | # papers | # parameters | # papers |
| Attack rate | 33 | 22 | 26 | 16 |
| Secondary attack rate | 9 | 8 | 7 | 6 |
| Doubling time | 1 | 1 | 0 | 0 |
| Growth rate (r) | 5 | 3 | 4 | 2 |
| Delay - admission to care>death | 9 | 8 | 7 | 6 |
| Delay - admission to care>discharge/recovery | 12 | 11 | 6 | 6 |
| Delay - generation time | 1 | 1 | 1 | 1 |
| Delay - incubation period | 49 | 39 | 42 | 32 |
| Delay - infectious period | 7 | 7 | 5 | 5 |
| Delay - latent period | 4 | 4 | 1 | 1 |
| Delay - other delay (go to section) | 55 | 27 | 34 | 20 |
| Delay - serial interval | 3 | 2 | 3 | 2 |
| Delay - symptom onset>admission to care | 38 | 33 | 29 | 25 |
| Delay - symptom onset>death | 16 | 13 | 14 | 11 |
| Delay - symptom onset>discharge/recovery | 8 | 6 | 6 | 4 |
| Delay - time in care (length of stay) | 15 | 14 | 7 | 7 |
| Mutations – mutation rate | 9 | 6 | 2 | 2 |
| Mutations – substitution rate | 3 | 1 | 3 | 1 |
| Overdispersion | 8 | 4 | 7 | 3 |
| Reproduction number (Basic, R0) | 34 | 25 | 16 | 13 |
| Reproduction number (Effective, Re) | 25 | 13 | 17 | 8 |
| Risk factors | 243 | 102 | 180 | 75 |
| Seroprevalence - IFA | 12 | 9 | 10 | 7 |
| Seroprevalence - IgG | 27 | 21 | 21 | 18 |
| Seroprevalence - Unspecified | 13 | 13 | 9 | 9 |
| Severity - case fatality rate (CFR) | 116 | 77 | 86 | 56 |
| Beta - per capita contact rate per unit of time | 7 | 6 | 7 | 6 |

Table B.4: Total parameters extracted and the total number of papers these parameters were extracted from. Middle column includes all parameters. Right column only includes those papers that scored higher than 0.5 for quality assessment (see Table A.2).

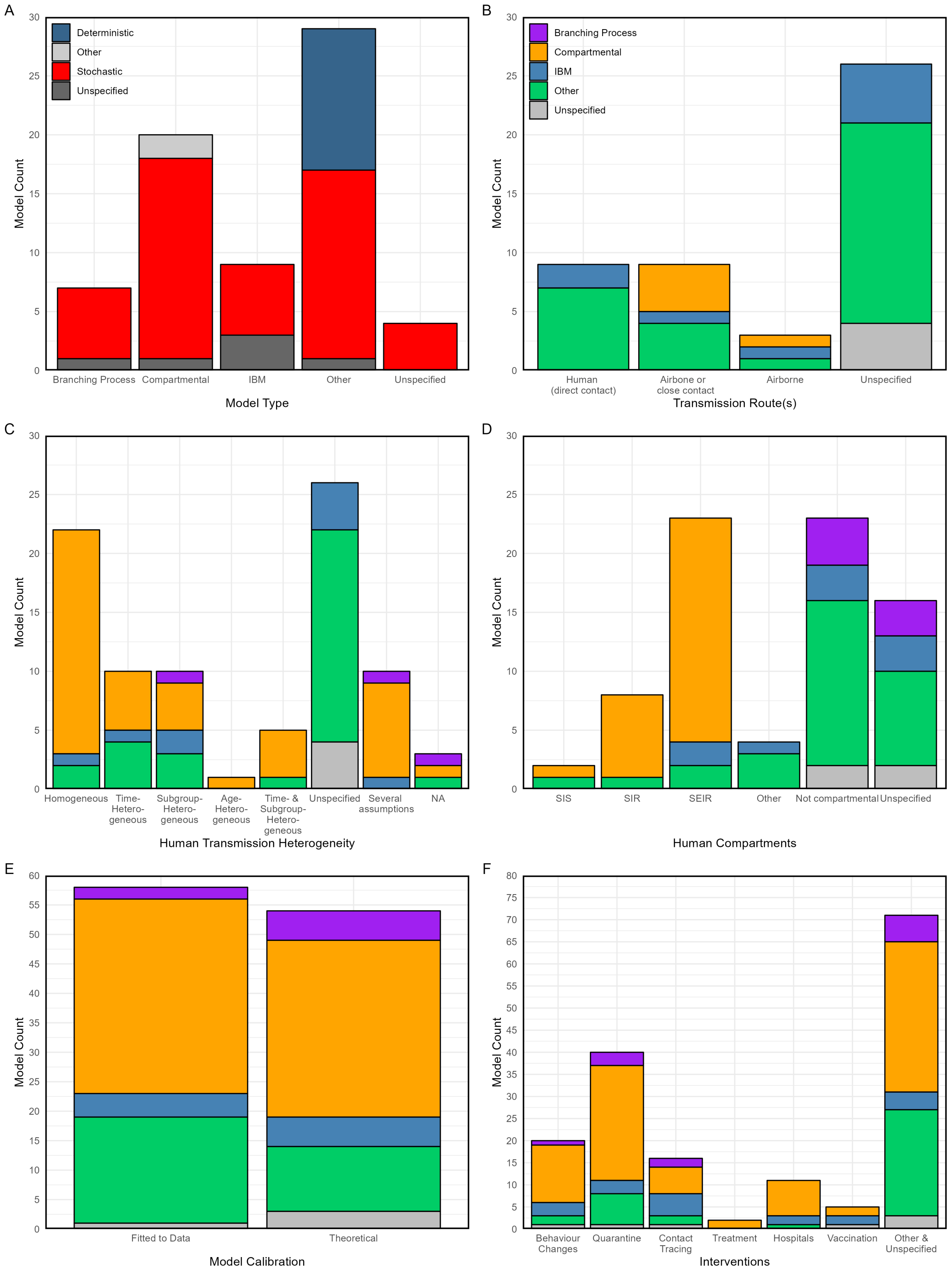

Figure B.3: Total published SARS transmission models separated by (A) model type, (B) routes of transmission considered, (C) transmission assumptions, (D) compartmental model form, (E) how the model was calibrated, (F) interventions directly modelled/considered. NA results are not plotted.

| Model Type | # Deterministic | # Stochastic | # Unspecified | # Both |
|---|---|---|---|---|
| Agent / Individual based | 0 | 6 | 3 | 0 |
| Agent / Individual based;Branching process | 0 | 0 | 1 | 0 |
| Branching process | 0 | 6 | 1 | 0 |
| Compartmental | 43 | 17 | 1 | 2 |
| Compartmental;Other | 1 | 0 | 0 | 0 |
| Other | 11 | 16 | 0 | 0 |
| Unspecified | 0 | 4 | 0 | 0 |

Table B.5: Total number of models extracted by model type and if the model is deterministic or stochastic. The model type and classification are extracted as stated in the paper.

#### B.3 Extraction Fields

##### B.3.1 Model extraction fields

| Data field | Expected data type | Variable name | Notes |
|---|---|---|---|
| Article ID | integer | article_id | ID to connect to article form |
| Model data ID | integer | model_data_id | ID assigned by database |
| Model type | character | model_type | General type of model - from dropdown list |
| Stochastic or deterministic | character | stoch_deter | Stochastic or deterministic model as reported |
| Transmission route | character | transmission_route | Transmission route(s) modelled - from dropdown list |
| Assumptions | character | assumptions | General assumptions for the model - from dropdown list |
| Compartmental type | character | compartmental_type | Specific type of compartmental model - from dropdown list |
| Theoretical model | logical | theoretical_model | Tick box whether the model was fitted to data (TRUE) or just theoretical (NA) |
| Intervention type | character | interventions_type | Type of intervention(s) modelled - from dropdown list |
| Code available | logical | code_available | Tick box whether code for model was publicly available and reported in the paper |

Table B.6: Model form fields: refer to epireview in Supplement C for dropdown options.

##### B.3.2 Parameter extraction fields

| Data field | Expected data type | Variable name | Notes |
|---|---|---|---|
| Article ID | integer | article_id | ID to connect to article form |
| Parameter data ID | integer | parameter_data_id | ID assigned by database |
| Parameter type | character | parameter_type | Category of parameter - see dropdown list |
| Parameter value | numeric | parameter_value | Central parameter value |
| Parameter exponent | integer | exponent | Parameter value exponent (base 10) |
| Inverse parameter | logical | inverse_param | Tick box to indicate that only inverse of parameter is reported (e.g., recovery rate from fitted model instead of infectious period) |
| Parameter unit | character | parameter_unit | Units for parameter value, applies to central estimate and ranges/uncertainty intervals - see dropdown list |
| Parameter value type | character | parameter_value_type | Type of central parameter value - see dropdown list |
| Parameter lower bound | numeric | parameter_lower_bound | Lower bound of the parameter range if a range was reported or if data are disaggregated |
| Parameter upper bound | numeric | parameter_upper_bound | Upper bound of the parameter range if a range was reported or if data are disaggregated |
| Parameter uncertainty - single type | character | parameter_uncertainty_singe_type | Type of uncertainty fpr central parameter value if single value was reported - see dropdown list |
| Parameter uncertainty - single value | numeric | parameter_uncertainty_single_value | Value for uncertainty for central parameter value if a single value was reported |

| | | | |
|---|---|---|---|
| Distribution type | logical | distribution_type | Type of distribution for estimated parameter - see dropdown list |
| First distribution parameter type | logical | distribution_par1_type | Type of value for first distribution parameter - see dropdown list |
| First distribution parameter value | logical | distribution_par1_value | Value for first distribution parameter (e.g. shape or scale parameter for a gamma distribution) |
| First distribution parameter uncertainty | logical | distribution_par1_uncertainty | Tick box for whether uncertainty is estimated for the first distribution parameter (TRUE) or not (FALSE) |
| Second distribution parameter type | logical | distribution_par2_type | Type of value for second distribution parameter - see dropdown list |
| Second distribution parameter value | logical | distribution_par2_value | Value for second distribution parameter (e.g. shape or scale parameter for a gamma distribution) |
| Second distribution parameter uncertainty | logical | distribution_par2_uncertainty | Tick box for whether uncertainty is estimated for the second distribution parameter (TRUE) or not (FALSE) |
| Disaggregated data available | logical | method_disaggregated | Tick box if disaggregated estimates are available (TRUE) or not (FALSE) |
| Parameter estimates disaggregated by | character | method_disaggregated_by | Categories for disaggregation of parameter estimates |
| Only disaggregated data available | logical | method_disaggregated_only | Tick box if ONLY disaggregated estimates are available (TRUE) or if a central estimate is also available (FALSE) |
| Is parameter from supplement? | logical | method_from_supplement | Tick box for whether parameter was extracted from supplement (TRUE) or not (FALSE) |
| Is parameter in a figure only? | logical | parameter_fromfigure | Tick box to indicate that parameter is plotted in a figure but not reported numerically in the text/table, and thus could not be extracted |
| Study population country | character | population_country | Country of the survey population - see dropdown list |
| Study population location | character | population_location | Region/district/province/city of the survey population - see dropdown list |
| Start day of study | integer | population_study_start_day | Study start day |
| Start month of study | character | population_study_start_month | Study start month - see dropdown list |
| Start year of study | integer | population_study_start_year | Study start year - see dropdown list |
| End day of study | integer | population_study_end_day | Study end day |
| End month of study | character | population_study_end_month | Study end month - see dropdown list |
| End year of study | integer | population_study_end_year | Study end year - see dropdown list |
| Survey timing related to outbreak | character | method_moment_value | Timing of the survey in relation to the outbreak, if specified in paper - see dropdown list |
| Study population sample size | integer | population_sample_size | Sample size of the population used for parameter estimation |
| Study population minimum age (years) | numeric | population_age_min | Minimum age of the survey population in years |
| Study population maximum age (years) | numeric | population_age_max | Maximum age of the survey population in years |
| Sex of study population | character | population_sex | Sex of survey population - see dropdown list |
| Population sample setting | character | population_sample_type | General setting of the survey - see dropdown list |
| Population group | character | population_group | Specific group of the survey population - see dropdown list |
| Genome site | character | genome_site | Site of genome or gene studied |
| Genomic sequence available? | logical | genomic_sequence_available | Tick box whether genomic sequence data are available (TRUE) or not (FALSE) |
| Reproduction number pathway | character | r_pathway | Transmission pathway that reproduction number is based on for vector-borne diseases |
| Method to estimate R | character | method_r | Method used for estimation of the reproduction number - see dropdown list |
| Other delay start point | character | other_delay_start | Start point for delays not in the parameter type dropdown list (e.g., delay from ... to ...) |
| Other delay end point | character | other_delay_end | End point for delays not in the parameter type dropdown list (e.g., delay from ... to ...) |

| | | | |
|---|---|---|---|
| Numerator | integer | cfr_ifr_numerator | Numerator of either CFR/IFR (deaths) or seroprevalence (number seropositive) estimates |
| Denominator | integer | cfr_ifr_denominator | Denominator of either CFR/IFR (cases) or seroprevalence (number tested) estimates |
| Is the CFR/IFR estimate adjusted? | character | cfr_ifr_method | Is the CFR/IFR estimate adjusted, unadjusted, or unspecified - see dropdown list |
| Outcome for risk factor(s) | character | riskfactor_outcome | Outcome for risk factor(s) - see dropdown list |
| Risk factor name | character | riskfactor_name | Risk factor name - see dropdown list |
| Risk factor occupation | character | riskfactor_occupation | If risk factor is an occupation, then specified occupation as risk factor - see dropdown list |
| Risk factor adjusted | character | riskfactor_adjusted | Adjustment status of risk factor(s)- see dropdown list |
| Risk factor significant | character | riskfactor_significant | Statistical significance of risk factor(s) - see dropdown list |
| Parameter class | character | parameter_class | General parameter class (delays, seroprevalence, reproduction numbers, mutations, severity, risk factors, relative contribution) |
| Uncertainty | character | uncertainty | Formatted uncertainty range from ‘parameter_uncertainty_lower_value‘ and ‘parameter_uncertainty_lower_value‘ in format x - x for ranges and x, x for confidence or credible intervals |
| Survey year | character | survey$_{year}$ | Dates of survey in format YYYY, YYYY-YYYY, MMM YYYY, or MMM-MMM YYYY from survey start and end variables |

Table B.7: Parameter form fields: refer to epireview in Supplement C for dropdown options.

#### B.4 Extracted Data

This section provides all extracted parameter and model data.

##### B.4.1 Transmission parameters

Table B.8 lists all extracted transmission parameters, in order of:

1. Basic Reproduction Numbers ($R_0$)
2. Effective Reproduction Numbers ($R_e$)
3. Growth rates
4. Overdispersion parameters
5. Beta (per capita contact rate per unit of time)
6. Attack rates
7. Secondary attack rates
8. Mutation rates
9. Substitution rates

| Parameter | Uncertainty | Disaggregated By | Gene | Method | Sample Size | Country | Study Dates | Study Setting | Study Group | Source |
|---|---|---|---|---|---|---|---|---|---|---|
| **Reproduction Number R0** | | | | | | | | | | |
| 0.2 | | | | Empirical (Contact Tracing) | 212 | Vietnam | 26 Feb 2003 - 28 Apr 2003 | Contact | Mixed Groups | Tuan (2007) |
| 0.58 | IQR: 0.24-1.18 | | | Unspecified | | Canada | | | | Chowell (2004) |
| 1.1 | | | | Growth Rate | | Canada, Hong Kong SAR, China, Singa-pore | 2003 - 25 Apr 2003 | Population | | Chowell (2003) |
| 1.1 | IQR: 0.44-2.29 | | | Unspecified | | Hong Kong SAR, China | | | | Chowell (2004) |
| 1.17 | IQR: 0.47-2.47 | | | Unspecified | | Singapore | | | | Chowell (2004) |
| 1.2 | | | | Growth Rate | | Hong Kong SAR, China | | | | Massad (2005) |
| 1.32 | | | | Growth Rate | | Canada | | | | Massad (2005) |
| 1.5 | | Region | | Compartmental Model | | Canada | 25 Feb 2003 - 6 Apr 2003 | Unspecified | Unspecified | Choi (2003b) |
| 1.63 | 90% CI: 0.54-2.65 | | | Branching Process | 57 | Singapore | 25 Feb 2003 - 22 Mar 2003 | | | Lloyd-Smith (2005) |
| 1.88 | 95% CI: 1.85-1.91 | | | Compartmental Model | | Taiwan, China | 25 Feb 2003 - 25 Jun 2003 | Unspecified | Unspecified | Wang (2012) |
| 1.88 | | | | Branching Process | 33 | China | 2003 | Hospital | | Lloyd-Smith (2005) |
| 2.09 | | | | Next Generation Matrix | | Singapore | 25 Mar 2003 - 27 Apr 2003 | Population | General Population | He (2023) |
| 2.1 | | | | Growth Rate | | Hong Kong SAR, China | 17 Mar 2003 - 15 May 2003 | Population | General Population | Zhou (2003) |
| 2.16 | | | | Compartmental Model | | China | 21 Apr 2003 - 24 May 2003 | Population | General Population | Wang (2003) |
| 1.0698 - 3.2524 | | | | Compartmental Model | | China | 27 Apr 2003 - 10 Jun 2003 | Population | General Population | Wang (2004c) |
| 2.23 | | | | Compartmental Model | 671 | Taiwan, China | | Population | General Population | Bombardt (2006) |
| 1.5 - 3 | | | | | | Canada, Hong Kong SAR, China, Singa-pore, Taiwan, China | | Unspecified | | Glass (2007) |
| 2.37 | | | | Compartmental Model | | China | 19 Apr 2003 - 21 Jun 2003 | Population | Persons Under Investigation | Wang (2006a) |
| 2.21 - 2.53 | | Time | | Compartmental Model | | Canada | 23 Feb 2003 - 11 Jun 2003 | Unspecified | Unspecified | Wang (2012) |
| 2.65 | 90% CI: 0.57-12.45 | | | | | Taiwan, China | 29 Apr 2003 - 8 May 2003 | Hospital | Persons Under Investigation | Liao (2008) |
| 2.65 | Normal-Log var: 2.55 | | | Other | | Taiwan, China | 24 Apr 2003 - 8 May 2003 | Unspecified | Unspecified | Chen (2006b) |

continued on next page

continued from previous page

| Parameter | Uncertainty | Disaggregated By | Gene | Method | Sample Size | Country | Study Dates | Study Setting | Study Group | Source |
|---|---|---|---|---|---|---|---|---|---|---|
| 2.7 | | | | Growth Rate | | Singapore | 17 Mar 2003 - 15 May 2003 | Population | General Population | Zhou (2003) |
| 2.7 | 95% CI: 2.2-3.7 | | | Compartmental Model | 1512 | Hong Kong SAR, China | 3 Mar 2003 - 6 May 2003 | Mixed | Mixed Groups | Riley (2003) |
| 2.2 - 3.6 | | Other, Time | | Growth Rate | 425 | | 15 Feb 2003 - 28 Mar 2003 | Mixed | Persons Under Investigation | Lipsitch (2003) |
| 2 - 4 | | | | Other | | Hong Kong SAR, China | | | | Fraser (2004) |
| 3.5256 | | | | Compartmental Model | | Canada | 25 Mar 2003 - | Population | Persons Under Investigation | Ding (2004) |
| 3.567 | | | | Compartmental Model | | Hong Kong SAR, China | 17 Mar 2003 - 1 Apr 2003 | Population | Persons Under Investigation | Ding (2004) |
| 1.5402 - 5.6304 | | Region | | Growth Rate | | China, Hong Kong SAR, China, Singapore, Taiwan, China | | Unspecified | Unspecified | Zhang (2004) |
| 3.8 | | | | Growth Rate | | China | 17 Mar 2003 - 15 May 2003 | Population | General Population | Zhou (2003) |
| 3.88 | Standard Deviation: 0.09 | | | Compartmental Model | 1755 | Hong Kong SAR, China | 15 Feb 2003 - 24 Jul 2003 | Unspecified | Unspecified | Lekone (2008) |
| 4.4544 | | | | Compartmental Model | | Singapore | 8 Mar 2003 - | Population | Persons Under Investigation | Ding (2004) |
| 4.587436164 | | | | Next Generation Matrix | | Canada, Hong Kong SAR, China | | Population | General Population | Ruan (2006) |
| 2.02 - 23.95 | 95% CI: 1.83-49.27 | Region | | Branching Process | 716 | Hong Kong SAR, China, Singapore | 15 Feb 2003 - 25 Mar 2003 | Population | General Population | Moser (2015) |
| 0.595 - 176 | | Method | | Next Generation Matrix | 390 | Hong Kong SAR, China | 11 Mar 2003 - 31 Jul 2003 | Hospital | Mixed Groups | Kwok (2007) |
| **Reproduction Number Re** | | | | | | | | | | |
| 0.001 | | | | Compartmental Model | 1755 | Hong Kong SAR, China | 15 Feb 2003 - 24 Jul 2003 | Unspecified | Unspecified | Lekone (2008) |
| 0.1 | | | | Compartmental Model | | China | 19 Apr 2003 - 21 Jun 2003 | Population | Persons Under Investigation | Wang (2006a) |
| 0.28 | | | | Branching Process | 43 | China | 2003 | Hospital | | Lloyd-Smith (2005) |
| 0.3 | 95% CI: 0.1-0.7 | | | Branching Process | | Vietnam | 12 Mar 2003 - Jul 2003 | Population | General Population | Wallinga (2004) |

continued on next page

continued from previous page

| Parameter | Uncertainty | Disaggregated By | Gene | Method | Sample Size | Country | Study Dates | Study Setting | Study Group | Source |
|---|---|---|---|---|---|---|---|---|---|---|
| 0.13 - 0.5 | | | | Empirical (Contact Tracing) | 2658 | China | 2003 | Household | Unspecified | Pitzer (2007) |
| 0.14 - 1 | | Time | | | 1512 | Hong Kong SAR, China | 3 Mar 2003 - 6 May 2003 | Mixed | Mixed Groups | Riley (2003) |
| 0.68 | | | | Branching Process | 114 | Singapore | 22 Mar 2003 | | | Lloyd-Smith (2005) |
| 0.7 | 95% CI: 0.7-0.8 | | | Branching Process | | Hong Kong SAR | 12 Mar 2003 - Jul 2003 | Population | General Population | Wallinga (2004) |
| 0.7 | 95% CI: 0.6-0.9 | | | Branching Process | | Singapore | 12 Mar 2003 - Jul 2003 | Population | General Population | Wallinga (2004) |
| 0.75 | 95% CrI: 0.65-0.85 | | | Compartmental Model | 354 | China | Feb 2003 - May 2003 | Unspecified | Persons Under Investigation | McBryde (2006) |
| 0.861 | Standard Deviation: 0.413 | | | Compartmental Model | | | 24 Mar 2003 - 2 May 2023 | | | Feng (2009a) |
| 1.4937 - 0.2485 | | Time | | Growth Rate | | China | 21 Apr 2003 - 12 May 2003 | Population | Persons Under Investigation | Zhang (2007) |
| 0.95 | 95% CI: 0.67-1.23 | | | Branching Process | | Canada, Singa-pore | 23 Feb 2003 - 11 May 2003 | Hospital | Persons Under Investigation | Chowell (2015) |
| 1 | 95% CI: 0.9-1.2 | | | Branching Process | | Canada | 12 Mar 2003 - Jul 2003 | Population | General Population | Wallinga (2004) |
| 1.2849 | | | | Compartmental Model | | Singapore | - 1 Apr 2003 | Population | Persons Under Investigation | Ding (2004) |
| 1.3901 | | | | Compartmental Model | | Canada | - 15 Apr 2003 | Population | Persons Under Investigation | Ding (2004) |
| 1.7068 | | | | Compartmental Model | | Hong Kong SAR, China | 1 Apr 2003 - 14 Apr 2003 | Population | Persons Under Investigation | Ding (2004) |
| 2.4 | 95% CI: 1.8-3.1 | | | Branching Process | | Vietnam | Feb 2003 - 11 Mar 2003 | Population | General Population | Wallinga (2004) |
| 2.7 | 95% CI: 1.8-3.6 | | | Branching Process | | Canada | Feb 2003 - 11 Mar 2003 | Population | General Population | Wallinga (2004) |
| 3.1 | 95% CI: 2.3-4 | | | Branching Process | | Singapore | Feb 2003 - 11 Mar 2003 | Population | General Population | Wallinga (2004) |
| 3.16 | Standard Deviation: 2.25 | | | Compartmental Model | | Hong Kong SAR, China | 24 Feb 2003 - 24 Mar 2003 | | | Feng (2009a) |
| 3.5 | 90% CrI: 1.5-7.7 | Time | | Growth Rate | 425 | | | | | Lipsitch (2003) |
| 3.6 | 95% CI: 3.1-4.2 | | | Branching Process | | Hong Kong SAR | Feb 2003 - 11 Mar 2003 | Population | General Population | Wallinga (2004) |
| 4.23 | | | | | | Taiwan, China | 6 May 2003 - 4 Jun 2003 | | | Hsieh (2004) |
| 4.8 | 95% CrI: 2.2-8.8 | | | Compartmental Model | 354 | China | Feb 2003 - May 2003 | Unspecified | Persons Under Investigation | McBryde (2006) |
| **Growth Rate** | | | | | | | | | | |
| 0.013 - 0.0713 | | Time | | Growth Rate | | China | 21 Apr 2003 - 12 May 2003 | Population | Persons Under Investigation | Zhang (2007) |
| 0.09 | | | | | | Hong Kong SAR, China | 17 Mar 2003 - 15 May 2003 | Population | General Population | Zhou (2003) |
| 0.12 | | | | | | Singapore | 17 Mar 2003 - 15 May 2003 | Population | General Population | Zhou (2003) |

continued on next page

continued from previous page

| Parameter | Uncertainty | Disaggregated By | Gene | Method | Sample Size | Country | Study Dates | Study Setting | Study Group | Source |
|---|---|---|---|---|---|---|---|---|---|---|
| 0.16 | | | | | | China | 17 Mar 2003 - 15 May 2003 | Population | General Population | Zhou (2003) |
| 0.142 - 0.859 per day | | | | | 250 | Canada | 23 Feb 2003 - 12 Jun 2003 | Community | | Hsieh (2006a) |
| **Overdispersion** | | | | | | | | | | |
| 2.6311 $10^{-11}$ | Standard Deviation: 14.077 $10^{-11}$ | | | Compartmental Model | 2048 | China | 20 Apr 2003 - 4 Jun 2003 | Hospital | Persons Under Investigation | Kong (2016) |
| 1.1882 $10^{-5}$ | Standard Deviation: 0.575 $10^{-5}$ | | | Compartmental Model | 2048 | China | 7 Mar 2003 - 20 Apr 2003 | Hospital | Persons Under Investigation | Kong (2016) |
| 0.0062 | | | | Branching Process | 43 | China | 2003 | Hospital | | Lloyd-Smith (2005) |
| 0.071 | 90% CI: 0.049-0.41 | | | Branching Process | 114 | Singapore | 22 Mar 2003 | | | Lloyd-Smith (2005) |
| 0.12 | 90% CI: 0.08-0.42 | | | Branching Process | 33 | China | 2003 | Hospital | | Lloyd-Smith (2005) |
| 0.16 | 90% CI: 0.11-0.64 | | | Branching Process | 57 | Singapore | 25 Feb 2003 - 22 Mar 2003 | | | Lloyd-Smith (2005) |
| 0.2 | 95% CI: 0.13-0.27 | | | Branching Process | | Canada | 23 Feb 2003 - 11 May 2003 | Hospital | Persons Under Investigation | Chowell (2015) |
| 331 mnc | 95% CI: 295-331 mnc | | | | 331 | Hong Kong SAR, China | 18 Mar 2003 - 20 Mar 2003 | Housing | | Riley (2003) |
| **Beta** | | | | | | | | | | |
| 0.062 Unspecified | | | | | 1512 | Hong Kong SAR, China | 3 Mar 2003 - 6 May 2003 | Mixed | Mixed Groups | Riley (2003) |
| 0 - 1.49 per day $10^{-1}$ | | Time | | Compartmental Model | | Hong Kong SAR, China | 14 Feb 2003 - 3 Jun 2003 | | | Mubayi (2021) |
| 0.149 | Standard Deviation: 0.003 | | | Compartmental Model | 1755 | Hong Kong SAR, China | 15 Feb 2003 - 24 Jul 2003 | Unspecified | Unspecified | Lekone (2008) |
| 0.347 per day | 95% CI: 0.3108-0.3837 per day | | | | 461 | Taiwan, China | 25 Feb 2003 - 25 Jun 2003 | Population | General Population | Hsieh (2007) |
| 0.5459 per day | Standard Deviation: 0.0335 per day | | | Compartmental Model | 2048 | China | 7 Mar 2003 - 4 Jun 2003 | Hospital | Persons Under Investigation | Kong (2016) |
| 0.68 | | | | | | Singapore | 2003 | Population | | Chowell (2003) |
| 0.75 | | | | | | Hong Kong SAR, China | 2003 | Population | | Chowell (2003) |
| **Attack Rate** | | | | | | | | | | |
| 0 | | Other | | | 363 | China | 2003 - 23 May 2023 | Contact | Persons Under Investigation | Zeng (2009) |
| 0 % | 95% CI: 0-26.4 % | Other | | | | China, Hong Kong SAR, China, Taiwan, China | | Travel | Persons Under Investigation | Olsen (2003) |

continued on next page

continued from previous page

| Parameter | Uncertainty | Disaggregated By | Gene | Method | Sample Size | Country | Study Dates | Study Setting | Study Group | Source |
|---|---|---|---|---|---|---|---|---|---|---|
| 0 % | 95% CI: 0-40.2 % | Other | | | | China, Hong Kong SAR, China, Taiwan, China | | Travel | Persons Under Investigation | Olsen (2003) |
| 8.9 $10^{-5}$ | | | | | | Hong Kong SAR, China | Mar 2003 - Jun 2003 | Population | General Population | Leung (2004c) |
| 18.6 $10^{-5}$ | | Age, Sex | | | | China | 8 Mar 2003 - 28 May 2003 | Population | General Population | Liang (2007) |
| 28.3 $10^{-5}$ | | Region | | | 632683 | China | 14 Mar 2003 - 22 May 2003 | Population | General Population | Liang (2003) |
| 21.993 per 10k | | Region | | | | China | 1 Mar 2003 - 16 Aug 2003 | Population | General Population | Liu (2005) |
| 465 $10^{-5}$ | | | | | | China | 5 Mar 2003 - 20 May 2003 | Hospital | Healthcare Workers | Liang (2004) |
| 0.25 - 0.88 % | | Time | | | 8662 | Canada | 23 Feb 2003 - 2 Jul 2003 | Population | General Population | Rea (2007) |
| 0.61 % | | Time | | | 40 | Hong Kong SAR, China | 25 Mar 2003 - 31 Mar 2003 | Hospital | Healthcare Workers | Ho (2003) |
| 0.7 % | 95% CI: 0.54-0.9 % | | | | 8662 | Canada | 23 Feb 2003 - 2 Jul 2003 | Population | General Population | Rea (2007) |
| 0.68 - 0.72 % | | Sex | | | 8662 | Canada | 23 Feb 2003 - 2 Jul 2003 | Population | General Population | Rea (2007) |
| 1 % | 95% CI: 0.8-1.1 % | | | | 23103 | Canada | 23 Feb 2003 - 1 Jul 2003 | Population | General Population | Svoboda (2004) |
| 0.42 - 1.68 % | | Age | | | 8662 | Canada | 23 Feb 2003 - 2 Jul 2003 | Population | General Population | Rea (2007) |
| 1.2 % | | Occupation, Region, Time | | | 339 | Hong Kong SAR, China | 4 Mar 2003 - 31 May 2003 | Hospital | Healthcare Workers | Lau (2004a) |
| 0.39 - 2.45 % | | Other | | | 8662 | Canada | 23 Feb 2003 - 2 Jul 2003 | Population | General Population | Rea (2007) |
| 4.5 % | | Other | | | 402 | China | 14 Mar 2003 - 22 May 2003 | Contact | Healthcare Workers | Liang (2003) |
| 0 - 10.36 % | | Level of Exposure | | | 8662 | Canada | 23 Feb 2003 - 2 Jul 2003 | Population | General Population | Rea (2007) |
| 6.3 % | 95% CI: 5.3-7.3 % | Other | | | | China | | Contact | Other | Pang (2003) |
| 6.7 % | 95% CI: 0.6-14 % | | | | 212 | Vietnam | 26 Feb 2003 - 28 Apr 2003 | Contact | Mixed Groups | Tuan (2007) |
| 7.1 % | | | | | 742 | Hong Kong SAR, China | | Hospital | Healthcare Workers | Ip (2004) |
| 7.4 % | | Region | | | 474 | China | 14 Mar 2003 - 22 May 2003 | Contact | Persons Under Investigation | Liang (2003) |
| 9.7 % | | Other | | | 1140 | China | 16 Apr 2003 - 12 May 2003 | Hospital | Mixed Groups | Wang (2006b) |
| 10 % | | Other | | | 70 | Singapore | 5 Apr 2023 - | Hospital | Mixed Groups | Tan (2004) |
| 11.2 % | 95% CI: 9.4-13 % | Age | | | | China | | Contact | Other | Pang (2003) |
| 0 - 32.5 % | | Other | | | | Singapore | 1 Mar 2003 - 31 May 2003 | Hospital | Healthcare Workers | Chen (2006f) |
| 16.4 % | | Other | | | 110 | China | 15 Mar 2003 - | Travel | General Population | Lei (2018) |
| 0 - 33.3 % | | Other | | | | Singapore | 1 Mar 2003 - 31 May 2003 | Hospital | Persons Under Investigation | Chen (2006f) |
| 6.3 - 28.8 | | Other | | | 669 | China | 2003 - 23 May 2023 | Contact | Persons Under Investigation | Zeng (2009) |
| 23.1 % | | Other | | | 65 | Hong Kong SAR, China | 4 Mar 2003 - 10 Mar 2003 | Hospital | Healthcare Workers | Wong (2004) |
| 10.3 - 60 % | | Occupation | | | | Canada | 14 Mar 2003 - 15 Apr 2003 | Hospital | Healthcare Workers | Varia (2003) |

continued on next page

continued from previous page

| Parameter | Uncertainty | Disaggregated By | Gene | Method | Sample Size | Country | Study Dates | Study Setting | Study Group | Source |
|---|---|---|---|---|---|---|---|---|---|---|
| 40.5 % | | Other, Time | | | 74 | Hong Kong SAR, China | 4 Mar 2003 - Mar 2003 | Hospital | Persons Under Investigation | Yu (2005) |
| 57 % | | | | | 80 | Singapore | Apr 2003 - Jun 2003 | Hospital | Healthcare Workers | Wilder-Smith (2005) |
| **Secondary Attack Rate** | | | | | | | | | | |
| 4.2 % | 95% CI: 1.5-7 % | | | | 212 | Vietnam | 26 Feb 2003 - 28 Apr 2003 | Contact | Mixed Groups | Tuan (2007) |
| 6.2 % | 95% CI: 3.9-8.6 % | | | | 417 | Singapore | 24 Feb 2003 - 29 Apr 2003 | | Other | Goh (2004) |
| 8 % | | Other, Time | | | 2139 | Hong Kong SAR, China | 4 Apr 2003 - 10 Jun 2003 | Population | Household Contacts Of Survivors | Lau (2004c) |
| 8.8 % | | Other | | | 697 | Hong Kong SAR, China | 28 Feb 2003 - 8 Jun 2003 | Household | Household Contacts Of Survivors | Chan (2004b) |
| 10.2 % | 95% CI: 6.7-23.5 % | | | | 176 | Canada | 25 May 2003 - 31 Oct 2003 | Household | Persons Under Investigation | Wilson-Clark (2006) |
| 14.9 % | 95% CI: 12.6-17.4 % | Other, Time | | | 881 | Hong Kong SAR, China | 4 Apr 2003 - 10 Jun 2003 | Household | General Population | Lau (2004c) |
| 18.7 % | | Occupation | | | 193 | Vietnam | 26 Feb 2003 - 24 Mar 2003 | Hospital | Healthcare Workers | Reynolds (2006) |
| 19 % | | | | | 31 | Canada | 23 Mar 2003 - | Hospital | Healthcare Workers | Scales (2003) |
| 18.5 - 66.7 % | | Other | | | 77 | China | 5 Feb 2003 - 4 May 2003 | Contact | Persons Under Investigation | Shen (2004) |
| **Mutation Rate** | | | | | | | | | | |
| 8.26 Unspecified $10^{-6}$ | SD_s: 2.16 Unspecified $10^{-6}$ | | | | 61 | China | 31 Jan 2003 - | Unspecified | Unspecified | MEC (2004) |
| 0.42 - 2.38 s/s/y $10^{-3}$ | | Method, Region | | | 16 | China, Hong Kong SAR, China, Singapore | 14 Apr 2003 - 29 Aug 2003 | Unspecified | Persons Under Investigation | Zhao (2004) |
| 0.121 % | | | | | 33 | | | Unspecified | Unspecified | Wang (2004b) |
| 0.636 % | | | | | 33 | | | Unspecified | Unspecified | Wang (2004b) |
| 0.079 - 3.333 % | | Other | | | 33 | | | Unspecified | Unspecified | Wang (2004b) |
| 8 Unspecified $10^{-6}$ | | | Synonymous; multiple sites comprising 91.25 | | 33 | | | Unspecified | Unspecified | Wang (2004b) |
| 4.3 Unspecified $10^{-6}$ | | | whole genome | | 12 | Singapore | - 2003 | Unspecified | Unspecified | Vega (2004) |
| 5.7 Unspecified $10^{-6}$ | | | whole genome | | 6 | Singapore | - 2003 | Unspecified | Unspecified | Vega (2004) |
| 0.11 - 0.07 m/g/g | | | whole genome | | 10 | Taiwan, China | Mar 2003 - Jun 2003 | Mixed | Persons Under Investigation | Yeh (2004) |
| **Substitution Rate** | | | | | | | | | | |

continued on next page

continued from previous page

| Parameter | Uncertainty | Disaggregated By | Gene | Method | Sample Size | Country | Study Dates | Study Setting | Study Group | Source |
|---|---|---|---|---|---|---|---|---|---|---|
| 0.8 - 2.38 s/s/y $10^{-3}$ | | | | | 11 | Hong Kong SAR, China, Singapore | 14 Apr 2003 - 29 Aug 2003 | Unspecified | Persons Under Investigation | Zhao (2004) |
| 0.81 - 9.22 s/s/y $10^{-3}$ | | Other | non-synonymous sites | | 16 | China, Hong Kong SAR, China, Singapore | 14 Apr 2003 - 29 Aug 2003 | Unspecified | Persons Under Investigation | Zhao (2004) |
| 0 - 8.5 s/s/y $10^{-3}$ | | Other | synonymous sites | | 16 | China, Hong Kong SAR, China, Singapore | 14 Apr 2003 - 29 Aug 2003 | Unspecified | Persons Under Investigation | Zhao (2004) |

Table B.8: Transmission parameters grouped by parameter type. Study characteristics are reported for each parameter estimate. Note that s/s/y stands for nucleotide substitutions per site per year and m/g/g stands for mutations per genome per generation. If an exponent is stated for the parameter value the same exponent applies for the uncertainty parameters (without stating it again).

##### B.4.2 Models

Table B.9 lists all extracted transmission models, in order of:

1. Agent / individual based models
2. Branching process models
3. Compartmental models
4. Other / unspecified models

| Transmission Route(s) | Human Transmission Heterogeneity | Human Compartments | Fitted | Interventions | Source |
|---|---|---|---|---|---|
| **Agent / Individual based - NA** | | | | | |
| Human-Human | Age, Groups | | Fitted | Behaviour Changes, Vaccination | Pourbohloul (2005) |
| | Unspecified | | Theoretical | Hospitals, Other, Quarantine | Huang (2010) |
| | | | Fitted | Behaviour Changes | Durham (2012) |
| **Agent / Individual based - Stochastic** | | | | | |
| Human-Human | Groups | SEIR | Theoretical | Contact Tracing | Duan (2013) |
| | | | Fitted | Contact Tracing, Quarantine | Fraser (2004) |
| | Groups | Other | Theoretical | Behaviour Changes, Contact Tracing, Hospitals, Other, Quarantine | Hsieh (2006b) |
| | Time | | Fitted | | Xiao (2017) |
| | | SEIR | Theoretical | Contact Tracing, Other | Duan (2021) |
| | | | Theoretical | Contact Tracing, Vaccination | Xu (2009) |
| **Agent / Individual based;Branching process - NA** | | | | | |
| | Time | | Theoretical | Other, Quarantine | Peak (2017) |
| **Branching Process** | | | | | |
| | | | Fitted | | Li (2004) |
| | Groups | | Fitted | Contact Tracing, Other, Quarantine | Lloyd-Smith (2005) |
| | Groups | | Theoretical | | Nandi (2018) |
| | Unspecified | | Theoretical | | Wallinga (2004) |
| | Unspecified | | Theoretical | | Goubar (2009) |
| | | | Theoretical | Quarantine | Klinkenberg (2006) |
| **Branching process - NA** | | | | | |
| Human-Human | | | Theoretical | Behaviour Changes, Contact Tracing, Other, Quarantine | Becker (2005) |
| **Compartmental - Deterministic** | | | | | |
| Human-Human | | SIR | Fitted | | Choi (2003b) |
| Human-Human | | SEIR | Theoretical | | Ruan (2006) |
| Human-Human | | SEIR | Theoretical | Contact Tracing, Quarantine | Nishiura (2004) |
| Human-Human | | Other | Fitted | Quarantine | Nishiura (2009) |
| Human-Human | | Other | Fitted | | Nishiura (2009) |
| Human-Human | | Other | Fitted | | Ng (2003) |
| Human-Human | | Other | Theoretical | Contact Tracing, Quarantine | Mubayi (2010) |
| Human-Human | | Other | Theoretical | Behaviour Changes, Other | Zhang (2007) |
| Human-Human | | | Fitted | Other | Denphedtnong (2013) |
| Human-Human | Age | SIR | Fitted | Contact Tracing, Other, Quarantine | Huo (2015) |
| Human-Human | Time | Other | Fitted | Other, Quarantine | Ding (2004) |
| Human-Human | Time, Unspecified | Other | Fitted | Quarantine | Zhou (2004) |
| Human-Human | Unspecified | Other | Fitted | Hospitals | Zhang (2004) |
| Human-Human | Unspecified | Other | Theoretical | Quarantine | Zhang (2005) |
| Human-Human | | SIR | Theoretical | | Khatua (2020) |
| Human-Human | | SEIR | Theoretical | | Jana (2020) |
| | | SIR | Fitted | | Mubayi (2021) |
| | | SEIR | Fitted | Quarantine | Bombardt (2006) |
| | | SEIR | Theoretical | Other, Quarantine | Yan (2008) |
| | | SEIR | Theoretical | Other, Quarantine | Safi (2010) |
| | | Other | Theoretical | Quarantine | Gumel (2004) |
| | | | Theoretical | Quarantine, Vaccination | Siriprapaiwan (2018) |
| | Age, Groups | | Fitted | Behaviour Changes | Chowell (2003) |
| | Groups | Other | Theoretical | Hospitals, Quarantine | Brauer (2015) |
| | Groups | Other | Fitted | Hospitals | He (2023) |
| | Groups, | Other | Theoretical | Behaviour Changes, Quarantine | Hsu (2007) |

continued on next page

continued from previous page

| Transmission Route(s) | Human Transmission Heterogeneity | Human Compartments | Fitted | Interventions | Source |
|---|---|---|---|---|---|
| | Groups, | Other | Theoretical | Behaviour Changes, Quarantine | Hsu (2006) |
| | Groups, Time | Other | Fitted | Behaviour Changes | Kwok (2007) |
| | Time | SEIR | Fitted | Other | Wang (2006a) |
| | Time | Other | Fitted | Behaviour Changes, Quarantine | Wang (2004c) |
| | Time | Other | Theoretical | Quarantine | McLeod (2006) |
| | Unspecified | SIR | Fitted | | Wang (2012) |
| | Unspecified | SIR | Fitted | | Hirose (2012) |
| | Unspecified | SEIR | Theoretical | | Alvarez (2015) |
| | Unspecified | SEIR | Theoretical | Other, Quarantine | Cantun-Avila (2021) |
| | Unspecified | Other | Theoretical | Behaviour Changes, Contact Tracing, Vaccination | Gjorgjieva (2005) |
| | Unspecified | Other | Theoretical | Quarantine | Zhu (2008) |
| | Unspecified | Other | Theoretical | Behaviour Changes | Wang (2015) |
| | Unspecified | Other | Theoretical | Behaviour Changes, Quarantine | Safi (2011) |
| | Unspecified | Other | Theoretical | | Naheed (2014) |
| | | SEIR | Fitted | | Kong (2016) |
| | | Other | Fitted | | Hsieh (2004) |
| | | Other | Fitted | Hospitals, Quarantine | Feng (2009a) |
| **Compartmental - Deterministic;Stochastic** | | | | | |
| Human-Human | Groups, Time | SIR | Fitted | Other | Zong-Mao (2012) |
| | Time | SEIR | Theoretical | Hospitals | Fukutome (2007) |
| **Compartmental - NA** | | | | | |
| | Groups, Time | Other | Theoretical | Hospitals, Quarantine | Webb (2004) |
| **Compartmental - Stochastic** | | | | | |
| Human-Human | | SEIR | Fitted | Behaviour Changes, Other | Nishiura (2005) |
| Human-Human | | Other | Theoretical | Contact Tracing, Quarantine, Treatment | Bogaards (2007) |
| Human-Human | | SIS | Fitted | Behaviour Changes, Treatment | Aya (2018) |
| Human-Human | Groups | SEIR | Theoretical | Behaviour Changes, Contact Tracing, Other, Quarantine | Lloyd-Smith (2003) |
| Human-Human | Time | SEIR | Fitted | | Lekone (2008) |
| Human-Human | | SEIR | Fitted | | Drake (2006) |
| | | SEIR | Fitted | | Lai (2013) |
| | | Other | Fitted | Quarantine | Colizza (2007) |
| | Groups | SEIR | Theoretical | | Nandi (2018) |
| | Groups | SEIR | Theoretical | | Hollingsworth (2007) |
| | Groups, Time | Other | Fitted | Behaviour Changes, Hospitals, Other | Riley (2003) |
| | Groups, Time | Other | Fitted | | McBryde (2006) |
| | Time | SEIR | Fitted | | Cori (2009) |
| | Unspecified | Other | Fitted | | Brockmann (2013) |
| | Unspecified | Other | Theoretical | Hospitals | Colizza (2008) |
| | | Other | Fitted | Quarantine | Lipsitch (2003) |
| | | SIR, SIS | Theoretical | | Maeno (2010) |
| **Compartmental;Other - Deterministic** | | | | | |
| | | Other | Fitted | Hospitals, Quarantine | Hsieh (2007) |
| **Other - Deterministic** | | | | | |
| Human-Human | | | Fitted | | Yoneyama (2012) |
| Human-Human | Unspecified | | Fitted | | Zhang (2004) |
| | Groups, Time | Other | Theoretical | Quarantine, Other | Roberts (2004) |
| | Time | SEIR | Theoretical | | Wang (2008) |
| | Unspecified | | Fitted | | Wang (2012) |
| | Unspecified | | Fitted | Contact Tracing, Quarantine | Han (2004b) |
| | Unspecified | | Fitted | Behaviour Changes | Wang (2003) |
| | Unspecified | Other | Fitted | | Zhou (2003) |
| | | | Fitted | | Pitzer (2007) |
| | | | Theoretical | | Masuda (2004) |
| | | | Fitted | | Hsieh (2006a) |
| **Other - Stochastic** | | | | | |
| Human-Human | Groups | | Theoretical | | Fujie (2007) |
| Human-Human | Groups | | Theoretical | Quarantine | Fang (2004) |
| Human-Human | Unspecified | | Fitted | | Kuk (2009) |
| Human-Human | | | Fitted | | yip (2008) |
| Human-Human | | | Theoretical | | Zhen (2007) |
| | | SIR | Theoretical | | Liao (2005) |
| | | | Theoretical | Other | Glass (2006) |
| | Groups | SIS | Theoretical | Other | Wan (2008) |
| | Time | | Fitted | | Glass (2007) |
| | Time | | Fitted | | Chen (2008) |
| | Unspecified | | Fitted | Behaviour Changes, Other, Quarantine | Chan (2006) |

continued on next page

continued from previous page

| Transmission Route(s) | Human Transmission Heterogeneity | Human Compartments | Fitted | Interventions | Source |
|---|---|---|---|---|---|
| | Unspecified | | Fitted | Contact Tracing, Quarantine | Liao (2008) |
| | | | Fitted | Other | Chen (2006b) |
| | | SEIR | Fitted | | Small (2006) |
| | | | Theoretical | | Cauchemez (2006a) |
| | | | Fitted | | Cao (2016) |
| **Unspecified - Stochastic** | | | | | |
| | Unspecified | | Theoretical | Behaviour Changes, Contact Tracing, Quarantine, Vaccination | Krumkamp (2009) |
| | Unspecified | | Theoretical | | Campbell (2018) |
| | Unspecified | | Theoretical | | Shi (2003) |
| | | | Fitted | | Cauchemez (2006b) |

Table B.9: Characteristics of extracted transmission models.

##### B.4.3 Natural history

Table B.10 lists all extracted natural history parameters. These are parameters capturing delays from one epidemiological period/event to another. These are given in order of:

1. Incubation period
2. Latent period
3. Infectious period
4. Symptom onset to admission delay
5. Symptomatic period
6. Other delay
7. Generation time
8. Serial interval
9. Admission to recovery delay
10. Admission to death delay
11. Time in Care
12. Symptom onset to recovery
13. Symptom onset to death

| Delay (days) | Statistic | Uncertainty | Disaggregated By | Sample Size | Country | Study Dates | Study Setting | Study Group | Source |
|---|---|---|---|---|---|---|---|---|---|
| **Incubation Period** | | | | | | | | | |
| 2.4 days | Mean | Other: 1.5 days | | 17 | China | 22 Dec 2002 - Mar 2003 | Hospital | General Population | Xiao (2003) |
| 3 days | Median | Range: 2-6 days | | 11 | Hong Kong SAR, China | 4 Mar 2003 - 10 Mar 2003 | Hospital | Persons Under Investigation | Wong (2004) |
| 3.5 days | Mean | SD_s: 3 days | Age, Sex | 10 | Canada | 23 Mar 2003 - | Hospital | Healthcare Workers | Avendano (2003) |
| 3.7 days | Mean | 95% CrI: 2.6-5.8 days | Other | 302 | Hong Kong SAR, China | Feb 2003 - Jul 2003 | Hospital | Mixed Groups | Virlogeux (2015) |
| 4 days | Median | Range: 2-10 days | | 42 | Canada | 14 Mar 2003 - 15 Apr 2003 | Hospital | Persons Under Investigation | Varia (2003) |
| 4 days | | Range: 2-8 days | | 22 | Hong Kong SAR, China, Taiwan, China | | Travel | Persons Under Investigation | Olsen (2003) |
| 4 days | Median | Range: 1-18 days | | 19 | Canada, Hong Kong SAR, China, United States | | Unspecified | Unspecified | Meltzer (2004) |
| 4 days | Median | Range_s: 2-8 days | | 7 | Singapore | Feb 2003 - Mar 2003 | Hospital | Mixed Groups | Hsu (2003) |
| 4 days | Median | Range_s: 3-6 days | | 32 | Taiwan, China | 17 Apr 2003 - 26 Apr 2003 | Hospital | Mixed Groups | Chen (2003) |
| 4 days | Mean | SD_s: 3 days | Age, Sex | 4 | Canada | 23 Mar 2003 - | Hospital | Healthcare Workers | Avendano (2003) |
| 4.09 days | Mean | Range: 0-14 days | | 16 | Hong Kong SAR, China | 2003 | Hospital | Persons Under Investigation | Tam (2007) |
| 4.4 days | Mean | SD_s: 4.6 days | | 168 | Hong Kong SAR, China | | Population | General Population | Lau (2010) |
| 4.6 days | Standard Deviation | 95% CrI: 3.6-6 days | Other | 234 | Hong Kong SAR, China | Feb 2003 - Jul 2003 | Hospital | Persons Under Investigation | Virlogeux (2015) |
| 4.6 days | Mean | Variance: 15.9 days | | 1755 | Hong Kong SAR, China | | Population | Persons Under Investigation | Leung (2009b) |
| 4.6 days | Mean | 95% CI: 3.8-5.8 days | | 81 | Hong Kong SAR, China | 15 Feb 2003 - 31 May 2003 | Unspecified | Persons Under Investigation | Leung (2004a) |
| 4.7 days | Mean | 95% CrI: 4.1-5.4 days | Other | 234 | Hong Kong SAR, China | Feb 2003 - Jul 2003 | Hospital | Persons Under Investigation | Virlogeux (2015) |
| 4.7 days | Mean | Range: 1-12 days | | 59 | Canada | 23 Feb 2003 - 1 Jul 2003 | Population | General Population | Svoboda (2004) |
| 4.8 days | Mean | 95% CrI: 4.2-5.5 days | Other | 1453 | Hong Kong SAR, China | Feb 2003 - Jul 2003 | Hospital | Mixed Groups | Virlogeux (2015) |
| 4.8 days | Median | Other: 3.8 days | | 46 | Taiwan, China | 18 Apr 2003 - 31 May 2003 | Hospital | Persons Under Investigation | Lim (2003) |
| 4.82 days | Mean | 95% CI: 4.23-5.5 days | | 2658 | China | 2003 | Household | | Pitzer (2007) |
| 4.83 days | Mean | Weibull Shape: 1.91 days | | 198 | Singapore | Mar 2003 - Mar 2003 | Mixed | Persons Under Investigation | Kuk (2005) |
| 5 days | Mean | Range: 2-10 days | | 42 | Canada | 14 Mar 2003 - 15 Apr 2003 | Hospital | Persons Under Investigation | Varia (2003) |
| 5 days | Median | Range: 1-15 days | | 7 | Canada | 23 Mar 2003 - | Hospital | Healthcare Workers | Scales (2003) |
| 5 days | Mean | | | 249 | Taiwan, China | 1 Nov 2002 - 31 Dec 2003 | Hospital | Persons Under Investigation | Liu (2016) |
| 5 days | Mean | Range: 2-15 days | | | Singapore | | Hospital | Mixed Groups | Leong (2006b) |
| 5 days | Median | Other: 9 days | | 50 | Singapore | 25 Feb 2003 - 11 May 2003 | Population | Persons Under Investigation | Goh (2006) |
| 5 days | | | Region | | Canada | 25 Feb 2003 - 6 Apr 2003 | Unspecified | Unspecified | Choi (2003b) |

continued on next page

continued from previous page

| Delay | Statistic | Uncertainty | Disaggregated By | Sample Size | Country | Study Dates | Study Setting | Study Group | Source |
|---|---|---|---|---|---|---|---|---|---|
| 5.1 days | Mean | Standard Deviation: 2.2 days | | 50 | Singapore | 25 Feb 2003 - 11 May 2003 | Population | Persons Under Investigation | Goh (2006) |
| 5.1 days | Mean | Normal-Log var: 18.3 days | Region | 317 | Canada, Hong Kong SAR, China | | Population | Persons Under Investigation | Cowling (2007) |
| 5.29 days | Mean | Variance: 12.33 days | Age, Occupation, Other, Region, Sex | 209 | China | 2003 | Mixed | Persons Under Investigation | Cai (2006) |
| 5.3 days | Mean | Gamma Scale: 0.26 days | | 85 | China | Feb 2003 - May 2003 | Unspecified | Persons Under Investigation | McBryde (2006) |
| 3 - 8 days | | | | 15 | Singapore | 24 Mar 2003 - 15 Apr 2003 | Hospital | Persons Under Investigation | Chow (2004) |
| 5.7 days | Mean | | | 33 | China | 5 Feb 2003 - 4 May 2003 | Hospital | Persons Under Investigation | Shen (2004) |
| 5.7 days | Mean | SD_s: 9.7 days | | 97 | China | | Population | General Population | Lau (2010) |
| 5.9 days | Mean | Standard Deviation: 3.5 days | | 96 | China | 30 Jan 2003 - 10 Mar 2003 | Hospital | Mixed Groups | Wu (2003) |
| 1 - 11 days | Other | | Other | 10 | Hong Kong SAR, China | 22 Feb 2003 - 22 Mar 2003 | Contact | Persons Under Investigation | Tsang (2003b) |
| 6 days | Median | Range_s: 1-15 days | | 98 | Taiwan, China | 1 Nov 2002 - 31 Dec 2003 | Hospital | Healthcare Workers | Liu (2016) |
| 6 days | Median | Range_s: 2-16 days | | 138 | Hong Kong SAR, China | 11 Mar 2003 - 25 Mar 2003 | Hospital | Persons Under Investigation | Lee (2003) |
| 2 - 10 days | | | | 15 | Taiwan, China | Apr 2003 - May 2003 | Population | Children | Chang (2004b) |
| 6 days | Median | IQR: 3-10 days | | 144 | Canada | 7 Mar 2003 - 10 Apr 2003 | Hospital | General Population | Booth (2003) |
| 6 days | Mean | | | | China | Apr 2003 - | Unspecified | Unspecified | Zhou (2004) |
| 6.4 days | Mean | 95% CI: 5.2-7.7 days | | 1425 | Hong Kong SAR, China | 20 Feb 2003 - 28 Apr 2003 | Hospital | General Population | Donnelly (2003) |
| 6.9 days | Mean | SD_s: 6.1 days | | 210 | Taiwan, China | | Population | General Population | Lau (2010) |
| 2 - 12 days | Other | | Level of Exposure | | Singapore | 28 Feb 2003 - 10 Apr 2003 | | | Hsu (2004) |
| 7 days | Median | Range_s: 4-12 days | | 13 | Singapore | Feb 2003 - Mar 2003 | Hospital | Mixed Groups | Hsu (2003) |
| 7.8 days | Mean | | | 595 | China | 5 Mar 2003 - 20 May 2003 | Hospital | Persons Under Investigation | Liang (2004) |
| 10 days | | | | 45 | Hong Kong SAR, China | Mar 2003 - | Hospital | Healthcare Workers | Cheng (2005b) |
| 10.66 days | Other | 95% CI: 9.24-13.68 days | | 128 | Hong Kong SAR, China | | Hospital | Persons Under Investigation | Farewell (2005) |
| 11.44 - 11.83 days | | | | | Hong Kong SAR, China | 17 Mar 2003 - 10 May 2003 | | | Ng (2003) |
| **Latent Period** | | | | | | | | | |
| 4 days | Median | Range: 2-11 days | | 111 | China | 16 Apr 2003 - 12 May 2003 | Hospital | Mixed Groups | Wang (2006b) |
| 4.49 days | Mean | SD_s: 2.63 days | | | China | | Unspecified | Unspecified | Bombardt (2006) |
| 5 days | Mean | | | | China | 19 Apr 2003 - 21 Jun 2003 | Population | Persons Under Investigation | Wang (2006a) |
| 6.868132 days | Mean | Range: 4.694836-6.997901 days | | 2048 | China | 7 Mar 2003 - 4 Jun 2003 | Hospital | Persons Under Investigation | Kong (2016) |
| **Infectious Period** | | | | | | | | | |

continued on next page

continued from previous page

| Delay | Statistic | Uncertainty | Disaggregated By | Sample Size | Country | Study Dates | Study Setting | Study Group | Source |
|---|---|---|---|---|---|---|---|---|---|
| 4 days | Mean | | | | China | 19 Apr 2003 - 21 Jun 2003 | Population | Persons Under Investigation | Wang (2006a) |
| 0.2064 days | Mean | Standard Deviation: 0.0118 days | | 2048 | China | 7 Mar 2003 - 4 Jun 2003 | Hospital | Persons Under Investigation | Kong (2016) |
| 5 - 10 days | Unspecified | | | | Hong Kong SAR, China | | | | Fraser (2004) |
| 8.3 days | | 95% CI: 5.8-14.3 days | | | Singapore | 2003 | | | Drake (2006) |
| 9.3 days | Mean | 95% CrI: 8.6-9.9 days | | 1467 | Hong Kong SAR, China | Mar 2003 - Jul 2003 | Hospital | Persons Under Investigation | Cori (2009) |
| 12.5 days | Mean | SD_s: 5.6 days | | | Hong Kong SAR, China | | Unspecified | Unspecified | Bombardt (2006) |
| 21.6 days | Median | 95% CI: 14.9-26.8 days | | | | | | | Peak (2017) |
| **Onset - Admission** | | | | | | | | | |
| 2 days | Mean | | | 98 | Taiwan, China | 1 Nov 2002 - 31 Dec 2003 | Hospital | Healthcare Workers | Liu (2016) |
| 2.4 days | Mean | Standard Deviation: 1.8 days | | 142 | Hong Kong SAR, China | 2003 | Hospital | Persons Under Investigation | Cheng (2004) |
| 2.7 days | Mean | | | | China | | Population | General Population | Lau (2010) |
| 2.7 days | Mean | | | 40 | Hong Kong SAR, China | 25 Mar 2003 - 5 May 2003 | Hospital | Healthcare Workers | Ho (2003) |
| 2.8 days | Mean | | | | Taiwan, China | | Population | General Population | Lau (2010) |
| 2.8 days | Mean | | | 303 | Hong Kong SAR, China | 1 Jun 2004 - 31 Dec 2004 | Hospital | Persons Under Investigation | Lee (2006b) |
| 2.89 days | Mean | SD_s: 2.1 days | | | | | Unspecified | Unspecified | Bombardt (2006) |
| 2.9 days | Mean | Standard Deviation: 2 days | | 96 | China | 30 Jan 2003 - 10 Mar 2003 | Hospital | Mixed Groups | Wu (2003) |
| 3 days | Median | Range: 1-12 days | | 76 | Taiwan, China | 8 Mar 2003 - 15 Jun 2003 | Hospital | Persons Under Investigation | Wang (2004a) |
| 3 days | Median | Range: 0-11 days | | 138 | Hong Kong SAR, China | 11 Mar 2003 - 25 Mar 2003 | Hospital | Persons Under Investigation | Sung (2004) |
| 3 days | Mean | | | 249 | Taiwan, China | 1 Nov 2002 - 31 Dec 2003 | Hospital | Persons Under Investigation | Liu (2016) |
| 3 days | Median | | | | Singapore | | Hospital | Mixed Groups | Leong (2006b) |
| 3 days | Median | | Disease Generation, Other, Symptoms | 401 | China | Dec 2002 - Jun 2003 | Hospital | Persons Under Investigation | Chen (2006c) |
| 3 days | Mean | Standard Deviation: 3.1 days | Other | 268 | Hong Kong SAR, China | 28 Feb 2003 - 8 Jun 2003 | Hospital | Healthcare Workers | Chan (2004b) |
| 3.1 days | Mean | Range_s: 0.2-16 days | Occupation, Other | 263 | Singapore | 13 Mar 2003 - 31 May 2003 | Hospital | Persons Under Investigation | Chong (2005) |
| 3.1 days | Mean | 95% CI: 0.79-2.08 days | Occupation, Other | 263 | Singapore | 13 Mar 2003 - 31 May 2003 | Hospital | Persons Under Investigation | Chong (2005) |
| 3.5 days | Mean | Gamma Scale: 0.37 days | | 354 | China | Feb 2003 - May 2003 | Unspecified | Persons Under Investigation | McBryde (2006) |
| 3.6 days | Mean | | | | Hong Kong SAR, China | | Population | General Population | Lau (2010) |
| 3.6 days | Mean | SD_s: 2.4 days | | 115 | Hong Kong SAR, China | 9 Mar 2003 - 31 May 2003 | Hospital | General Population | Chan (2003c) |

continued on next page

continued from previous page

| Delay | Statistic | Uncertainty | Disaggregated By | Sample Size | Country | Study Dates | Study Setting | Study Group | Source |
|---|---|---|---|---|---|---|---|---|---|
| 3.8 days | Mean | 95% CI: 3.7-4 days | Age, Occupation, Other, Region, Sex, Time | 5258 | China | 16 Nov 2002 - 28 May 2003 | Population | Persons Under Investigation | Feng (2009b) |
| 3.8 days | Mean | 95% CI: 3.7-4 days | | | China | 16 Nov 2002 - 28 May 2003 | Population | Persons Under Investigation | Feng (2009c) |
| 3.96 days | Mean | SD_s: 4.28 days | | 5327 | China | Nov 2002 - Jun 2003 | Population | Persons Under Investigation | Cao (2016) |
| 4 days | Mean | Range: 0-13 days | | 45 | Vietnam | 26 Feb 2003 - 28 Apr 2003 | Hospital | Persons Under Investigation | Tuan (2007) |
| 2 - 6 days | Median | IQR: 1-14 days | Time | | China | | | | Pang (2003) |
| 4 days | | IQR: 2-6 days | | 273 | Canada | Feb 2003 - Jun 2003 | Hospital | Persons Under Investigation | Muller (2006) |
| 3 - 5 days | Mean | | | 1425 | Hong Kong SAR, China | 20 Feb 2003 - 28 Apr 2003 | Hospital | General Population | Donnelly (2003) |
| 4.1 days | Mean | | | 1876 | China | 8 Mar 2003 - 28 May 2003 | Hospital | Persons Under Investigation | Liang (2007) |
| 4.3 days | Mean | Standard Deviation: 1.6 days | | | China | 14 Mar 2003 - 22 May 2003 | Hospital | Persons Under Investigation | Liang (2003) |
| 4.4 days | | SD_s: 2.7 days | | 1312 | Hong Kong SAR, China | 2003 | Hospital | General Population | Chan (2007) |
| 4.6 days | Mean | Range_s: 1-8 days | Age, Sex | 14 | Canada | 23 Mar 2003 - | Hospital | Healthcare Workers | Avendano (2003) |
| 4.8 days | Mean | SD_e: 2.3 days | | 29 | Taiwan, China | 28 Mar 2003 - 30 Jun 2003 | Hospital | Persons Under Investigation | Jang (2004) |
| 2.2 - 7.4 days | Mean | | Time | 1755 | Hong Kong SAR, China | 15 Feb 2003 - 31 May 2003 | Mixed | Persons Under Investigation | Leung (2004a) |
| 5 days | Median | IQR_s: 2-7 days | | 38 | Canada | - 15 Apr 2003 | Hospital | Persons Under Investigation | Fowler (2003) |
| 5 days | Median | Range_s: 0-17 days | | 219 | China | Feb 2003 - May 2003 | Population | General Population | Chang (2006b) |
| 6 days | Mean | | | 3 | Vietnam | 26 Feb 2003 - 28 Apr 2003 | Contact | Persons Under Investigation | Tuan (2007) |
| 6 days | Median | | | 20 | Singapore | 28 Feb 2003 - 10 Apr 2003 | Hospital | | Hsu (2004) |
| 6 days | Median | Range_s: 0-9 days | Age, Sex, Time | 20 | Singapore | Feb 2003 - Mar 2003 | Hospital | Mixed Groups | Hsu (2003) |
| 6.8 days | Mean | | Time | | Singapore | 3 Mar 2003 - 9 Mar 2003 | Population | Persons Under Investigation | Goh (2006) |
| **Symptomatic Period** | | | | | | | | | |
| 10.3 days | Unspecified | | | 62 | Taiwan, China | 8 Mar 2003 - 15 Jun 2003 | Hospital | Persons Under Investigation | Wang (2004a) |
| **Other Human Delay** | | | | | | | | | |
| 0.16 days | Median | 95% CI: 0-0.67 days | | | | | | | Peak (2017) |
| 1.2 days | Mean | Range_s: 0-10 days | | 27 | China | 22 Apr 2003 - | Hospital | Mixed Groups | Han (2004a) |
| 2.1 days | Mean | Standard Deviation: 3.2 days | | | China | 14 Mar 2003 - 22 May 2003 | Hospital | Persons Under Investigation | Liang (2003) |
| 2.11 days | Mean | | | | Taiwan, China | 6 May 2003 - 4 Jun 2003 | | | Hsieh (2004) |
| 1.203 - 3.6398 days | Mean | | Other, Time | 475 | Taiwan, China | 2003 - 31 Dec 2004 | Population | General Population | Hsieh (2005) |
| 2 - 4 days | | | Other | 105 | Singapore | 1 Mar 2003 - 31 May 2003 | Hospital | Mixed Groups | Chen (2006f) |

continued on next page

continued from previous page

| Delay | Statistic | Uncertainty | Disaggregated By | Sample Size | Country | Study Dates | Study Setting | Study Group | Source |
|---|---|---|---|---|---|---|---|---|---|
| 3 days | Median | IQR: 2-5 days | | 144 | Canada | 7 Mar 2003 - 10 Apr 2003 | Hospital | General Population | Booth (2003) |
| 3.7 days | Mean | Range: 0-12 days | | 44 | Hong Kong SAR, China | 14 Mar 2003 - 10 Jun 2003 | Hospital | Children | Leung (2004c) |
| 3.9 days | Mean | Standard Deviation: 3.2 days | | 225 | Canada | 23 Feb 2003 - 1 Jul 2003 | Population | General Population | Svoboda (2004) |
| 3.9 days | Mean | SD_s: 4.8 days | | 21 | Singapore | 18 Mar 2003 - | Hospital | | Ho (2004) |
| 4 days | Mean | Other: 3.6-4.6 days | | 390 | Hong Kong SAR, China | 11 Mar 2003 - 31 Jul 2003 | Hospital | Persons Under Investigation | Kwok (2007) |
| 4.2 days | Median | Range_s: 0-12 days | Other | 105 | Singapore | 1 Mar 2003 - 31 May 2003 | Hospital | Mixed Groups | Chen (2006f) |
| 4.4 days | Mean | Standard Deviation: 2.3 days | | 218 | Hong Kong SAR, China | 26 Feb 2003 - 31 Mar 2003 | Hospital | Persons Under Investigation | Tsang (2003a) |
| 4.6 days | Mean | Range_s: 1-13 days | | 17 | Taiwan, China | 20 Mar 2003 - 5 Jul 2003 | Hospital | Persons Under Investigation | Chen (2005d) |
| 5 days | Median | Range: 3-7 days | | 10 | Hong Kong SAR, China | 22 Feb 2003 - 22 Mar 2002 | Contact | Persons Under Investigation | Tsang (2003b) |
| 5 days | Mean | Range_s: 1-10 days | | 29 | Taiwan, China | 28 Mar 2003 - 30 Jun 2003 | Hospital | Persons Under Investigation | Jang (2004) |
| 5.6 days | Mean | Range_s: 2-9 days | | 6 | China | 22 Apr 2003 - | Hospital | Mixed Groups | Han (2004a) |
| 5.8 days | Mean | SD_s: 2.8 days | | 25 | Taiwan, China | 28 Mar 2003 - 30 Jun 2003 | Hospital | Mixed Groups | Jang (2004) |
| 5.8 days | Mean | SD_s: 5.4 days | | 167 | Hong Kong SAR, China | 10 Mar 2003 - 5 Jun 2003 | Hospital | Persons Under Investigation | Chan (2005) |
| 6.37 days | Mean | 95% CI: 5.29-7.75 days | | 1425 | Hong Kong SAR, China | 20 Feb 2003 - 28 Apr 2003 | Hospital | General Population | Donnelly (2003) |
| 6.4 days | Mean | SD_s: 6 days | | 54 | | 12 Mar 2003 - 18 Apr 2003 | Hospital | Persons Under Investigation | Gomersall (2004) |
| 6.5 days | Mean | SD_s: 2.9 days | | 15 | Taiwan, China | 28 Mar 2003 - 30 Jun 2003 | Hospital | Mixed Groups | Jang (2004) |
| 6.8 days | Mean | SD_s: 2.9 days | | 29 | Taiwan, China | 28 Mar 2003 - 30 Jun 2003 | Hospital | Persons Under Investigation | Jang (2004) |
| 7.1 days | Mean | Range: 1-17 days | | 44 | Hong Kong SAR, China | 14 Mar 2003 - 10 Jun 2003 | Hospital | Children | Leung (2004c) |
| 7.5443 days | Mean | | | 327 | Taiwan, China | 2003 - 31 Dec 2004 | Other | General Population | Hsieh (2005) |
| 7.7647 days | Mean | | | 17 | Taiwan, China | 2003 - 31 Dec 2004 | Other | General Population | Hsieh (2005) |
| 8 days | Median | IQR_s: 5-10 days | | 38 | Canada | - 15 Apr 2003 | Hospital | Persons Under Investigation | Fowler (2003) |
| 8 days | Median | IQR_s: 6-12 days | | 29 | Canada | - 15 Apr 2003 | Hospital | Persons Under Investigation | Fowler (2003) |
| 8.3 days | Mean | Range: 3-15 days | | 9 | Hong Kong SAR, China | 14 Mar 2003 - 10 Jun 2003 | Hospital | Children | Leung (2004c) |
| 8.3 days | Mean | Range_s: 5-13 days | | 10 | China | 22 Apr 2003 - | Hospital | Mixed Groups | Han (2004a) |
| 8.5 days | Median | IQR_s: 5-22.3 days | | 54 | | 12 Mar 2003 - 18 Apr 2003 | Hospital | Persons Under Investigation | Gomersall (2004) |
| 8.6 days | Mean | Standard Deviation: 14 days | Other | 20 | Hong Kong SAR, China | 9 Mar 2003 - 28 Apr 2003 | Hospital | Unspecified | Chueng (2004) |
| 9.2 days | Mean | SD_s: 4 days | | 29 | Taiwan, China | 28 Mar 2003 - 30 Jun 2003 | Hospital | Persons Under Investigation | Jang (2004) |
| 9.2 days | Unspecified | Range: 2-21 days | | 234 | Singapore | | Hospital | Mixed Groups | Leong (2006a) |

continued on next page

continued from previous page

| Delay | Statistic | Uncertainty | Disaggregated By | Sample Size | Country | Study Dates | Study Setting | Study Group | Source |
|---|---|---|---|---|---|---|---|---|---|
| 9.5 days | | Standard Deviation: 4.7 days | | 67 | Hong Kong SAR, China | 13 Mar 2003 - 31 May 2003 | Hospital | | Gomersall (2006) |
| 9.6 days | Mean | SD_s: 4.6 days | | 54 | | 12 Mar 2003 - 18 Apr 2003 | Hospital | Persons Under Investigation | Gomersall (2004) |
| 9.7 days | Mean | SD_s: 2.2 days | | 9 | Taiwan, China | 28 Mar 2003 - 30 Jun 2003 | Hospital | Mixed Groups | Jang (2004) |
| 10.5 days | Median | IQR_s: 5-28 days | | | Canada | - 15 Apr 2003 | Hospital | Persons Under Investigation | Fowler (2003) |
| 10.7 days | Mean | SD_s: 3.8 days | | 30 | China | 22 Apr 2003 - | Hospital | Mixed Groups | Han (2004a) |
| 11.38 days | Mean | | | | Taiwan, China | 6 May 2003 - 4 Jun 2003 | | | Hsieh (2004) |
| 3 - 20 days | | | | 21 | China | 22 Apr 2003 - | Hospital | Mixed Groups | Han (2004a) |
| 12.56 days | Mean | | | | Taiwan, China | 6 May 2003 - 4 Jun 2003 | | | Hsieh (2004) |
| 12.7 days | | SD_s: 6.3 days | | 1312 | Hong Kong SAR, China | 2003 | Hospital | General Population | Chan (2007) |
| 13 days | | IQR: 6-24 days | | 67 | Hong Kong SAR, China | 13 Mar 2003 - 31 May 2003 | Hospital | | Gomersall (2006) |
| 13 days | Median | Range: 2-60 days | | 45 | Hong Kong SAR, China | 24 Mar 2003 - 4 May 2003 | Hospital | Persons Under Investigation | Chu (2005) |
| 13.1 days | Mean | SD_s: 10.3 days | | 21 | Taiwan, China | 28 Mar 2003 - 30 Jun 2003 | Hospital | Mixed Groups | Jang (2004) |
| 14 days | Median | IQR_s: 8-25 days | | 27 | | 12 Mar 2003 - 18 Apr 2003 | Hospital | Persons Under Investigation | Gomersall (2004) |
| 14 days | | | Region | | Canada | 25 Feb 2003 - 6 Apr 2003 | Unspecified | Unspecified | Choi (2003b) |
| 14 days | | Range: 7-30 days | Other | 80 | Hong Kong SAR, China | 20 Mar 2003 - 26 May 2003 | Hospital | Unspecified | Cheng (2005a) |
| 18.8 days | Mean | Range_s: 11-40 days | | 5 | Taiwan, China | 28 Mar 2003 - 30 Jun 2003 | Hospital | Mixed Groups | Jang (2004) |
| 24.31 days | Mean | | | | Taiwan, China | 6 May 2003 - 4 Jun 2003 | | | Hsieh (2004) |
| 30 days | Median | Range: 2-81 days | | 45 | Hong Kong SAR, China | 24 Mar 2003 - 4 May 2003 | Hospital | Persons Under Investigation | Chu (2005) |
| 33.7 days | Mean | Standard Deviation: 13.5 days | Other | 20 | Hong Kong SAR, China | 9 Mar 2003 - 28 Apr 2003 | Hospital | Unspecified | Chueng (2004) |
| 35.2 days | Mean | Standard Deviation: 15.4 days | | 45 | Hong Kong SAR, China | 24 Mar 2003 - 4 May 2003 | Hospital | Persons Under Investigation | Chu (2005) |
| **Generation Time** | | | | | | | | | |
| 10.6 days | Other | | | 1512 | Hong Kong SAR, China | 3 Mar 2003 - 6 May 2003 | Mixed | Mixed Groups | Riley (2003) |
| **Serial Interval** | | | | | | | | | |
| 8.4 days | Mean | Standard Deviation: 3.8 days | Time | 205 | Singapore | 25 Feb 2003 - 5 May 2003 | Population | Persons Under Investigation | Lipsitch (2003) |
| 10 days | Mean | Standard Deviation: 2.8 days | | | Singapore | 25 Feb 2003 - Mar 2003 | Population | Persons Under Investigation | Lipsitch (2003) |
| 7.42 - 16.86 days | Mean | 95% CI: 6.77-18.55 days | Region | 716 | Hong Kong SAR, China, Singapore | 15 Feb 2003 - 25 Mar 2003 | Population | General Population | Moser (2015) |
| **Admission - Recovery** | | | | | | | | | |
| 15.9 days | Mean | Range: 1-101 days | | | Singapore | | Hospital | Unspecified | Leong (2006a) |

continued on next page

continued from previous page

| Delay | Statistic | Uncertainty | Disaggregated By | Sample Size | Country | Study Dates | Study Setting | Study Group | Source |
|---|---|---|---|---|---|---|---|---|---|
| 19 days | Mean | | | 249 | Taiwan, China | 1 Nov 2002 - 31 Dec 2003 | Hospital | Persons Under Investigation | Liu (2016) |
| 19.5 days | Median | | | 7 | Canada | 23 Mar 2003 - | Hospital | Healthcare Workers | Scales (2003) |
| 22.6 days | Mean | SD_s: 22 days | | 25 | Taiwan, China | 28 Mar 2003 - 30 Jun 2003 | Hospital | Mixed Groups | Jang (2004) |
| 23 days | Mean | Gamma Scale: 0.18 days | | 354 | China | Feb 2003 - May 2003 | Unspecified | Persons Under Investigation | McBryde (2006) |
| 23.1 days | Mean | SD_s: 7.8 days | | | Hong Kong SAR, China | | Unspecified | Unspecified | Bombardt (2006) |
| 23.5 days | Mean | Variance: 62.1 days | | 1425 | Hong Kong SAR, China | 20 Feb 2003 - 28 Apr 2003 | Hospital | General Population | Donnelly (2003) |
| 23.94 days | | | | | Taiwan, China | 6 May 2003 - 4 Jun 2003 | | | Hsieh (2004) |
| 25.19 days | Mean | SD_s: 22.43 days | | | China | Nov 2002 - Jun 2003 | Population | Persons Under Investigation | Cao (2016) |
| 29.6 days | Mean | Range: 3-101 days | | | Singapore | | Hospital | Other | Leong (2006a) |
| 29.7 days | Mean | 95% CI: 29.3-30 days | Age, Occupation, Other, Region, Sex, Time | 4591 | China | 16 Nov 2002 - 28 May 2003 | Population | Persons Under Investigation | Feng (2009b) |
| 29.7 days | Mean | 95% CI: 29.3-30 days | | | China | 16 Nov 2002 - 28 May 2003 | Population | Persons Under Investigation | Feng (2009c) |
| **Admission - Death** | | | | | | | | | |
| 6 days | Median | | | 8 | Taiwan, China | 26 Mar 2003 - 25 May 2003 | Hospital | Persons Under Investigation | Wong (2003) |
| 9.4 days | Mean | | Age | 41 | China | 14 Mar 2003 - 22 May 2003 | Hospital | Persons Under Investigation | Liang (2003) |
| 11.7 days | Mean | 95% CI: 10.6-13 days | Age, Occupation, Other, Region, Sex, Time | 343 | China | 16 Nov 2002 - 28 May 2003 | Population | Persons Under Investigation | Feng (2009b) |
| 17 days | Mean | Gamma Scale: 0.068 days | | 354 | China | Feb 2003 - May 2003 | Unspecified | Persons Under Investigation | McBryde (2006) |
| 17 days | Median | | | 25 | Hong Kong SAR, China | 22 Feb 2003 - 31 May 2003 | Hospital | Other | Chan (2004a) |
| 17.3 days | Mean | 95% CI: 15.8-18.8 days | | | China | 16 Nov 2002 - 28 May 2003 | Population | Persons Under Investigation | Feng (2009c) |
| 27 days | Median | | | 52 | Hong Kong SAR, China | 22 Feb 2003 - 31 May 2003 | Hospital | Persons Under Investigation | Chan (2004a) |
| 35.9 days | Mean | Variance: 572.9 days | | 1425 | Hong Kong SAR, China | 20 Feb 2003 - 28 Apr 2003 | Hospital | General Population | Donnelly (2003) |
| 36.87 days | | | | | Taiwan, China | 6 May 2003 - 4 Jun 2003 | | | Hsieh (2004) |
| **Time In Care** | | | | | | | | | |
| 3 days | Median | Range: 1-14 days | | 90 | United States | 17 Mar 2003 - 30 Jul 2003 | Population | Persons Under Investigation | Schrag (2004) |
| 10 days | Median | IQR: 6-15 days | | 144 | Canada | 7 Mar 2003 - 10 Apr 2003 | Hospital | General Population | Booth (2003) |
| 12 days | Median | Range: 1-101 days | | 234 | Singapore | | Hospital | Mixed Groups | Leong (2006b) |
| 14 days | Mean | Range_s: 12-15 days | | 14 | Canada | 23 Mar 2003 - | Hospital | Healthcare Workers | Avendano (2003) |
| 17.2 days | Mean | Standard Deviation: 8 days | | 96 | China | 30 Jan 2003 - 10 Mar 2003 | Hospital | Mixed Groups | Wu (2003) |
| 18 days | | Range: 5-38 days | | 16 | Taiwan, China | 1 May 2003 - 15 Jun 2003 | Hospital | Healthcare Workers | Chiu (2004) |

continued on next page

continued from previous page

| Delay | Statistic | Uncertainty | Disaggregated By | Sample Size | Country | Study Dates | Study Setting | Study Group | Source |
|---|---|---|---|---|---|---|---|---|---|
| 18.7 days | Mean | Standard Deviation: 12 days | | 78 | Taiwan, China | Aug 2003 - Feb 2004 | Hospital | Persons Under Investigation | Liu (2009b) |
| 22.1 days | Mean | Standard Deviation: 3.1 days | Symptoms, Time | 75 | China | 24 Mar 2003 - 28 Mar 2003 | | | Peiris (2003) |
| 23 days | Mean | | Other | 1606 | Hong Kong SAR, China | 2 Apr 2003 - May 2003 | Hospital | Persons Under Investigation | Ghani (2005) |
| 23.1 days | Mean | Other: 11.9 days | | 78 | China | 22 Dec 2002 - Mar 2003 | Hospital | General Population | Xiao (2003) |
| 23.1 days | Unspecified | Other: 12.3 days | | 98 | China | Dec 2002 - Jun 2003 | Hospital | General Population | Mo (2006) |
| 24.5 days | Mean | Standard Deviation: 7.4 days | | 62 | Vietnam | 26 Feb 2003 - May 2003 | Hospital | Persons Under Investigation | Vu (2004) |
| 24.9 days | Mean | IQR_s: 14.8-33.3 days | Disease Generation | 67 | Taiwan, China | 20 Mar 2003 - 5 Jul 2003 | Hospital | Persons Under Investigation | Chen (2005d) |
| 29.02 days | Mean | | | 801 | China | Mar 2003 - Jun 2003 | Hospital | Mixed Groups | Lu (2005) |
| **Onset - Recovery** | | | | | | | | | |
| 19 days | Median | | | | Taiwan, China | | Population | General Population | Lau (2010) |
| 23 days | Median | | | | Hong Kong SAR, China | | Population | General Population | Lau (2010) |
| 24 days | Mean | | | 98 | Taiwan, China | 1 Nov 2002 - 31 Dec 2003 | Hospital | Healthcare Workers | Liu (2016) |
| 26 days | Mean | Gamma Scale: 0.22 days | | 354 | China | Feb 2003 - May 2003 | Unspecified | Persons Under Investigation | McBryde (2006) |
| 26.47 days | Mean | Variance: 25.8-27.2 days | Age, Sex | 1755 | Hong Kong SAR, China | 15 Feb 2003 - 31 May 2003 | Mixed | Persons Under Investigation | Leung (2004a) |
| 26.5 days | Mean | Variance: 194.9 days | | 1755 | Hong Kong SAR, China | | Population | Persons Under Investigation | Leung (2009b) |
| 32.79 days | Mean | | | 801 | China | Mar 2003 - Jun 2003 | Hospital | Mixed Groups | Lu (2005) |
| 36 days | Median | | | | China | | Population | General Population | Lau (2010) |
| **Onset - Death** | | | | | | | | | |
| 10 days | Median | | | | Taiwan, China | | Population | General Population | Lau (2010) |
| 11 days | Median | | Sex | 53 | China | 15 Mar 2003 - 14 May 2003 | Population | General Population | Zhang (2006) |
| 12 days | Median | Range: 4-42 days | | 15 | Taiwan, China | 8 Mar 2003 - 15 Jun 2003 | Hospital | Persons Under Investigation | Wang (2004a) |
| 12 days | Mean | | | 249 | Taiwan, China | 1 Nov 2002 - 31 Dec 2003 | Hospital | Persons Under Investigation | Liu (2016) |
| 12.8 days | Mean | Range: 4-31 days | | 23 | China | 14 Mar 2003 - 22 May 2003 | Hospital | Persons Under Investigation | Liang (2003) |
| 14.5 days | Mean | | | 98 | Taiwan, China | 1 Nov 2002 - 31 Dec 2003 | Hospital | Healthcare Workers | Liu (2016) |
| 19 days | Median | IQR_s: 11-22 days | | 13 | Canada | - 15 Apr 2003 | Hospital | Persons Under Investigation | Fowler (2003) |
| 21 days | Mean | Gamma Scale: 0.11 days | | 354 | China | Feb 2003 - May 2003 | Unspecified | Persons Under Investigation | McBryde (2006) |
| 21 days | Median | | | | Hong Kong SAR, China | | Population | General Population | Lau (2010) |
| 23.5 days | Mean | SD_s: 13.2 days | | | Hong Kong SAR, China | | Unspecified | Unspecified | Bombardt (2006) |
| 23.66 days | Mean | Variance: 22-25.3 days | Age, Sex | 1755 | Hong Kong SAR, China | 15 Feb 2003 - 31 May 2003 | Mixed | Persons Under Investigation | Leung (2004a) |

continued on next page

continued from previous page

| Delay | Statistic | Uncertainty | Disaggregated By | Sample Size | Country | Study Dates | Study Setting | Study Group | Source |
|---|---|---|---|---|---|---|---|---|---|
| 23.7 days | | Variance: 221 days | | 1755 | Hong Kong SAR, China | | Population | Persons Under Investigation | Leung (2009b) |
| 24 days | Median | | | | China | | Population | General Population | Lau (2010) |
| 24.8 days | Mean | Range: 9-57 days | | 17 | Canada | 14 Mar 2003 - 15 Apr 2003 | Hospital | Persons Under Investigation | Varia (2003) |
| 27 days | Mean | Range: 5-108 days | | 20 | Canada | | Hospital | Persons Under Investigation | Hwang (2005) |
| 32.4 days | Mean | Standard Deviation: 10.6 days | | 34 | Hong Kong SAR, China | | Hospital | Persons Under Investigation | Hung (2009) |

Table B.10: Human Delays grouped by published parameter. Study characteristics are reported for each parameter value.

###### B.4.4 CFR

Table B.11 lists all extracted CFRs by country/countries of reporting.

| CFR (%) | Uncertainty | Disaggregated By | Method | Deaths | Sample Size | Study Dates | Study Setting | Study Group | Source |
|---|---|---|---|---|---|---|---|---|---|
| 25.9 | | | Naive | 14 | 54 | 12 Mar 2003 - 18 Apr 2003 | Hospital | Persons Under Investigation | Gomersall (2004) |
| **Canada** | | | | | | | | | |
| 12.5 | | | Naive | 1 | 8 | 17 Mar 2003 - 4 Jul 2003 | | | Yoshikura (2011) |
| 13.3 | | Age, Other | Naive | 17 | 128 | 14 Mar 2003 - 10 Jul 2003 | Hospital | Persons Under Investigation | Varia (2003) |
| 13.4 - 18.7 | | Other | | | | 2 Mar 2003 - 21 Apr 2003 | Population | General Population | Lau (2009) |
| 16.9 | | | Naive | 38 | 225 | 23 Feb 2003 - 1 Jul 2003 | Population | General Population | Svoboda (2004) |
| 30 | | Region | | | | 25 Feb 2003 - 6 Apr 2003 | Unspecified | Unspecified | Choi (2003b) |
| **Canada; China; Hong Kong SAR; China; Singapore; Taiwan; China; Vietnam** | | | | | | | | | |
| 9.56 | | Region | Naive | 774 | 8096 | | Unspecified | Unspecified | Hirose (2007) |
| 10.4 | | | Adjusted | | | 10 Apr 2003 - 21 Apr 2003 | Unspecified | Unspecified | Galvani (2003) |
| 14.7 | | | Adjusted | | | 10 Apr 2003 - 12 May 2003 | Unspecified | Unspecified | Galvani (2003) |
| **China** | | | | | | | | | |
| 1 | | | Naive | 1 | 96 | 30 Jan 2003 - 10 Mar 2003 | Hospital | Mixed Groups | Wu (2003) |
| 3.3 | 95% CI: 2.1-4.4 % | Age, Occupation, Other, Sex | Naive | 30 | 917 | | Population | General Population | Lau (2010) |
| 3.6 | | | Naive | 54 | 1504 | 16 Nov 2002 - 2003 | Population | General Population | Na (2009) |
| 3.8 | | | Naive | 55 | 1454 | 16 Nov 2002 - 30 Apr 2003 | Community | General Population | Xu (2004) |
| 3.84 | | | Adjusted | | | Apr 2003 - May 2003 | Population | Persons Under Investigation | Cui (2003) |
| 5 | | | Naive | 1 | 20 | 27 Mar 2003 - 4 Jul 2003 | | | Yoshikura (2011) |
| 5.31 | Range_s: 4.86-5.87 % | Sex | Naive | 134 | 2521 | 15 Mar 2003 - 14 May 2003 | Population | General Population | Zhang (2006) |
| 5.36 | | | Adjusted | | | Apr 2003 - May 2003 | Population | Persons Under Investigation | Cui (2003) |
| 5.4 | | | Naive | 24 | 449 | 16 Nov 2002 - 2003 | Population | General Population | Na (2009) |
| 5.58 | | | Adjusted | | | Apr 2003 - May 2003 | Population | Persons Under Investigation | Cui (2003) |
| 6.23 | | | Unspecified | 25 | 401 | Dec 2002 - Jun 2003 | Hospital | Persons Under Investigation | Chen (2006c) |
| 6.4 | | Age | Unspecified | 156 | 2444 | 5 Mar 2003 - 20 May 2003 | Hospital | Persons Under Investigation | Liang (2004) |
| 6.4 | | Age, Occupation, Other, Region, Sex, Time | Naive | 343 | 5327 | 16 Nov 2002 - 2003 | Population | General Population | Na (2009) |
| 6.4 | | | Unspecified | 5327 | 343 | 16 Nov 2002 - 28 May 2003 | Population | Persons Under Investigation | Feng (2009c) |
| 6.55 | | Region | Adjusted | 349 | 5327 | Apr 2003 - May 2003 | Population | Persons Under Investigation | Cui (2003) |

continued on next page

continued from previous page

| CFR (%) | Uncertainty | Disaggregated By | Method | Deaths | Sample Size | Study Dates | Study Setting | Study Group | Source |
|---|---|---|---|---|---|---|---|---|---|
| 7 | | | Naive | 5 | 75 | 24 Mar 2003 - 28 Mar 2003 | | | Peiris (2003) |
| 6.7 | | | Naive | 8 | 120 | 22 Apr 2003 - | Hospital | Mixed Groups | Han (2004a) |
| 7.28 | | | Naive | 120 | 1649 | 2003 - 7 Jul 2003 | Population | Persons Under Investigation | Liu (2006a) |
| 7.3 | | | Naive | 184 | 2522 | 16 Nov 2002 - 2003 | Population | General Population | Na (2009) |
| 7.56 | | Region | Adjusted | 185 | 2445 | 1 Mar 2003 - 16 Aug 2003 | Hospital | Persons Under Investigation | Liu (2005) |
| 7.6 | | Occupation, Region, Time | Unspecified | 192 | 2521 | 8 Mar 2003 - 28 May 2003 | Population | Persons Under Investigation | Liang (2007) |
| 7.62 | | Time | | | | 21 Apr 2003 - 13 Jun 2003 | Other | Persons Under Investigation | Chen (2009a) |
| 7.66 | | Age, Disease Generation, Occupation, Other, Region, Sex, Time | Naive | 193 | 2521 | Mar 2003 - 16 Aug 2003 | Mixed | Persons Under Investigation | Chen (2005b) |
| 7.66 | | | Adjusted | | | Apr 2003 - May 2003 | Population | Persons Under Investigation | Cui (2003) |
| 8 | | Time | Unspecified | 191 | 2521 | 21 Apr 2003 - 2 Jul 2003 | Unspecified | Persons Under Investigation | Yip (2005a) |
| 8 | | | Adjusted | | | Apr 2003 - May 2003 | Population | Persons Under Investigation | Cui (2003) |
| 8.4 | | | Adjusted | 190 | | 5 Mar 2003 - 20 May 2003 | Hospital | Persons Under Investigation | Liang (2004) |
| 8.7 | | Age, Method | Adjusted | 41 | 473 | 14 Mar 2003 - 22 May 2003 | Population | Persons Under Investigation | Liang (2003) |
| 9 | | | Naive | 7 | 78 | 22 Dec 2002 - Mar 2003 | Hospital | General Population | Xiao (2003) |
| 9.9 | | | Naive | 28 | 282 | 16 Nov 2002 - 2003 | Population | General Population | Na (2009) |
| 11.7 | | | Unspecified | 13 | 111 | 16 Apr 2003 - 12 May 2003 | Hospital | Persons Under Investigation | Wang (2006b) |
| 12 | | | Naive | 21 | 175 | 16 Nov 2002 - 2003 | Population | General Population | Na (2009) |
| 12.7 | | | Naive | | | 16 Nov 2002 - 30 Apr 2003 | Community | General Population | Xu (2004) |
| 15 | | Age, Occupation, Other, Sex | Naive | 17 | 111 | 2003 - 6 2003 | Hospital | Mixed Groups | Wei (2009) |
| 10 - 20 | | Other | Unspecified | | | 3 Mar 2003 - | Hospital | Mixed Groups | Cooper (2009) |
| 20 | | Disease Generation | Unspecified | | | 5 Feb 2003 - 4 May 2003 | Hospital | Persons Under Investigation | Shen (2004) |
| 1 - 52 | | | | | | 21 Apr 2003 - 13 Jun 2003 | Other | Persons Under Investigation | Chen (2004) |
| 26.7 | | | Naive | 8 | 30 | 22 Apr 2003 - | Hospital | Mixed Groups | Han (2004a) |
| **Hong Kong SAR; China** | | | | | | | | | |
| 0 | | | Unspecified | | | Mar 2003 - Jun 2003 | Population | General Population | Leung (2004c) |
| 0.064 - 0.1442 | | Method, Time | Adjusted | | | 2 Apr 2003 - May 2003 | Hospital | Persons Under Investigation | Jewell (2007) |
| 0.24 - 3.69 | | Other, Region | Adjusted | | | | Unspecified | Unspecified | Hirose (2007) |
| 2.3 | 95% CI: 0-6.8 % | | | 1 | 44 | 16 Apr 2003 - | Hospital | Persons Under Investigation | Chan (2003b) |
| 3.6 | | | Naive | 5 | 138 | 11 Mar 2003 - | Hospital | Persons Under Investigation | Lee (2003) |
| 7.6 | | | Naive | | | 22 Feb 2003 - 31 May 2003 | Hospital | Persons Under Investigation | Chan (2004a) |

continued on next page

continued from previous page

| CFR (%) | Uncertainty | Disaggregated By | Method | Deaths | Sample Size | Study Dates | Study Setting | Study Group | Source |
|---|---|---|---|---|---|---|---|---|---|
| 9.1 | | | Naive | 119 | 1312 | 2003 | Hospital | General Population | Chan (2007) |
| 10.1 | | | Adjusted | 22 | 218 | 26 Feb 2003 - 31 Mar 2003 | Hospital | Persons Under Investigation | Tsang (2003a) |
| 10.9 | | | Naive | 15 | 138 | 11 Mar 2003 - 28 Jul 2003 | Hospital | Persons Under Investigation | Sung (2004) |
| 11.8 | | | Naive | 172 | 1462 | Mar 2003 - Jul 2003 | Population | General Population | Antonio (2005) |
| 12 | 95% CI: 8-16 % | Age | Naive | 32 | 267 | 26 Feb 2003 - 31 Mar 2003 | Hospital | Mixed Groups | Choi (2003a) |
| 12 | | | Naive | 24 | 201 | 26 Feb 2003 - 10 Apr 2003 | Hospital | General Population | Chau (2004) |
| 12.5 | | Other | Naive | 10 | 80 | 20 Mar 2003 - 26 May 2003 | Hospital | Unspecified | Cheng (2005a) |
| 12.9 | 95% CI: 0-25.8 % | | | | 31 | 16 Apr 2003 - | Hospital | Persons Under Investigation | Chan (2003b) |
| 13.2 | 95% CI: 9.8-16.8 % | Age | | | | 20 Feb 2003 - 28 Apr 2003 | Hospital | General Population | Donnelly (2003) |
| 14 | 95% CI: 5.2-26.3 % | | | | 343 | 16 Apr 2003 - | Hospital | General Population | Chan (2003b) |
| 14.2 | | Age | Adjusted | | 1606 | 2 Apr 2003 - May 2003 | Hospital | Persons Under Investigation | Ghani (2005) |
| 15 | | Symptoms | Naive | 3 | 20 | 9 Mar 2003 - 28 Apr 2003 | Hospital | Unspecified | Chueng (2004) |
| 15.6 | 95% CI: 9.8-22.8 % | | | | 634 | 16 Apr 2003 - | Hospital | General Population | Chan (2003b) |
| 15.7 | | | | 18 | 115 | 9 Mar 2003 - 31 May 2003 | Hospital | General Population | Chan (2003c) |
| 17 | | Time | Unspecified | 299 | 1755 | 12 Mar 2003 - 31 Jul 2003 | Hospital | Persons Under Investigation | Yip (2005a) |
| 17 | | Age, Occupation, Sex | Naive | 302 | 1755 | Feb 2003 - Jul 2003 | Hospital | Persons Under Investigation | Virlogeux (2015) |
| 17 | | | Naive | 299 | 1755 | | Population | Persons Under Investigation | Leung (2009b) |
| 17.1 | | Time | | | | 19 Mar 2003 - 4 Jun 2003 | Other | Persons Under Investigation | Chen (2009a) |
| 17.16 | Standard Error: 1.35 % | | Adjusted | | | - 25 May 2003 | Unspecified | Unspecified | Hirose (2009) |
| 17.16 | Normal SD: 1.35 % | | Adjusted | | | - 25 May 2003 | Unspecified | Unspecified | Hirose (2007) |
| 17.2 | | | Naive | 302 | 1755 | 19 Mar 2003 - 2 Apr 2003 | | | Nishiura (2009) |
| 17.2 | 95% CI: 15.4-19 % | Age, Occupation, Other, Sex | Naive | 302 | 1755 | | Population | General Population | Lau (2010) |
| 17.2 | | Age, Occupation, Other, Sex, Symptoms, Time | Unspecified | 302 | 1755 | 15 Feb 2003 - 31 May 2003 | Mixed | Persons Under Investigation | Leung (2004a) |
| 17.2 | | | Adjusted | | | 15 Feb 2003 - 24 Jul 2003 | Unspecified | Unspecified | Lekone (2008) |
| 17.2 | | | Naive | 302 | 1755 | | Hospital | Persons Under Investigation | Cowling (2006) |
| 15.4 - 19.2 | | Other | | | | | Population | General Population | Lau (2009) |
| 17.3 | Normal SD: 0.92 % | | Adjusted | | | - 11 Jul 2003 | Unspecified | Unspecified | Hirose (2007) |
| 15 - 20 | | Time | | | | Apr 2003 - 31 Jul 2003 | Hospital | Persons Under Investigation | Yip (2005b) |

continued on next page

continued from previous page

| CFR (%) | Uncertainty | Disaggregated By | Method | Deaths | Sample Size | Study Dates | Study Setting | Study Group | Source |
|---|---|---|---|---|---|---|---|---|---|
| 18 | | | | | | 19 Mar 2003 - 13 Jun 2003 | Other | Persons Under Investigation | Chen (2004) |
| 18.1 | 95% CI: 10.5-28.1 % | | Adjusted | | | 19 Mar 2003 - 27 Mar 2003 | | | Nishiura (2009) |
| 22.1 | | | Naive | 34 | 154 | | Hospital | Persons Under Investigation | Hung (2009) |
| 3.7 - 64.7 | | Age, Occupation, Sex | | | | Mar 2003 - 22 Sep 2003 | Hospital | Persons Under Investigation | Karlberg (2004) |
| 43.3 | 95% CI: 35.2-52.4 % | Age | | | | 20 Feb 2003 - 28 Apr 2003 | Hospital | General Population | Donnelly (2003) |
| 60 | | | Naive | | | 22 Feb 2003 - 31 May 2003 | Hospital | Other | Chan (2004a) |
| **Singapore** | | | | | | | | | |
| 11.8 | | | Unspecified | 30 | 234 | | Hospital | Mixed Groups | Leong (2006a) |
| 11.8 | | | Naive | 30 | | | Hospital | Mixed Groups | Leong (2006b) |
| 13.9 | | Age | Naive | 33 | 238 | 25 Feb 2003 - 11 May 2003 | Population | Persons Under Investigation | Goh (2006) |
| 14 | | | Naive | 1 | 7 | 5 Apr 2023 - | Hospital | Mixed Groups | Tan (2004) |
| 15.53 | | Time | | | | 14 Mar 2003 - 4 Jun 2003 | Other | Persons Under Investigation | Chen (2009a) |
| 15 - 19 | | Time | | | | Apr 2003 - | Hospital | Persons Under Investigation | Yip (2005b) |
| 18 | | | | | | 14 Mar 2003 - 13 Jun 2003 | Other | Persons Under Investigation | Chen (2004) |
| **Taiwan; China** | | | | | | | | | |
| 6.25 | | | Naive | 1 | 16 | 1 May 2003 - 15 Jun 2003 | Hospital | Healthcare Workers | Chiu (2004) |
| 6.5 | | | Unspecified | 4 | 62 | 8 Mar 2003 - 15 Jun 2003 | Hospital | Other | Wang (2004a) |
| 10.7 | | | Naive | 37 | 346 | 14 Mar 2003 - 30 Jul 2003 | Hospital | Persons Under Investigation | Chen (2005c) |
| 12.24 | | | Unspecified | 12 | 98 | 1 Nov 2002 - 31 Dec 2003 | Hospital | Healthcare Workers | Liu (2016) |
| 12.5 | | | Naive | 1 | 8 | 1 May 2003 - 20 Jun 2003 | | | Yoshikura (2011) |
| 13.8 | | | Naive | 4 | 29 | 28 Mar 2003 - 30 Jun 2003 | Hospital | Persons Under Investigation | Jang (2004) |
| 14.1 | | | Adjusted | | | 25 Feb 2003 - 25 Jun 2003 | Population | General Population | Hsieh (2007) |
| 15 | | | Naive | 8 | | 26 Mar 2003 - 25 May 2003 | Hospital | Persons Under Investigation | Wong (2003) |
| 17 | | | Naive | 8 | 46 | 18 Apr 2003 - 31 May 2003 | Hospital | Persons Under Investigation | Lim (2003) |
| 21.1 | | | Naive | 73 | 346 | 14 Mar 2003 - 30 Jul 2003 | Hospital | Persons Under Investigation | Chen (2005c) |
| 21.1 | | | Naive | 73 | 346 | | Population | General Population | Chang (2006a) |
| 24.5 | | | Unspecified | 61 | 249 | 1 Nov 2002 - 31 Dec 2003 | Hospital | Persons Under Investigation | Liu (2016) |
| 27 | | Age, Level of Exposure, Other, Sex | Naive | 181 | 668 | 14 Mar 2003 - 30 Jul 2003 | Hospital | Persons Under Investigation | Chen (2005c) |
| 27.6 | 95% CI: 24.2-31 % | Age, Occupation, Other, Sex | Naive | 180 | 664 | | Population | General Population | Lau (2010) |
| 30.8 | | | Adjusted | 24 | 78 | Aug 2003 - Feb 2004 | Hospital | Persons Under Investigation | Liu (2009b) |

continued on next page

continued from previous page

| CFR (%) | Uncertainty | Disaggregated By | Method | Deaths | Sample Size | Study Dates | Study Setting | Study Group | Source |
|---|---|---|---|---|---|---|---|---|---|
| 31.3 | | Disease Generation | Naive | 21 | 67 | 20 Mar 2003 - 5 Jul 2003 | Hospital | Persons Under Investigation | Chen (2005d) |
| 38 | | | Unspecified | 20 | 52 | 26 Apr 2003 - 25 May 2003 | Hospital | Persons Under Investigation | Ko (2004a) |
| 47 | | | Naive | 8 | | 26 Mar 2003 - 25 May 2003 | Hospital | Persons Under Investigation | Wong (2003) |
| 78.6 | | | Unspecified | 11 | 14 | 8 Mar 2003 - 15 Jun 2003 | Hospital | Other | Wang (2004a) |
| **United States** | | | | | | | | | |
| 0 | | | Naive | 0 | 398 | 17 Mar 2003 - 30 Jul 2003 | Population | Persons Under Investigation | Schrag (2004) |
| **Vietnam** | | | | | | | | | |
| 9.7 | | | Naive | 6 | 62 | 26 Feb 2003 - May 2003 | Hospital | Persons Under Investigation | Vu (2004) |
| 22 - 50 | | Occupation | Unspecified | | | 26 Feb 2003 - 24 Mar 2003 | Hospital | Healthcare Workers | Reynolds (2006) |

Table B.11: Case Fatality Rates (CFRs) grouped by country. Study characteristics are reported for each CFR estimate.

###### B.4.5 Seroprevalence

Table B.12 lists all extracted seroprevalence estimates by country/countries of reporting.

| Seroprevalence (%) | Uncertainty | Disaggregated By | Assay | Number Seropositive | Sample Size | Study Dates | Study Setting | Study Group | Source |
|---|---|---|---|---|---|---|---|---|---|
| **Canada** | | | | | | | | | |
| **96.1** | | | Unspecified | 124 | 129 | 23 Feb 2003 - 1 Jul 2003 | Population | Persons Under Investigation | Svoboda (2004) |
| **China** | | | | | | | | | |
| 34.8 - 100 | | Time | IFA | | | Feb 2003 - Mar 2003 | Hospital | Other | Shi (2005) |
| **1.2** | | | IgG | | | 16 Nov 2002 - 30 Apr 2003 | Community | General Population | Xu (2004) |
| **13** | | | IgG | | | 16 Nov 2002 - 30 Apr 2003 | Trade | Animal Workers | Xu (2004) |
| **26** | | | IgG | 8 | 31 | 28 Apr 2003 - 28 Aug 2003 | Population | General Population | Wu (2004) |
| **0.3 - 88.9** | | Other | IgG | | | May 2003 - May 2003 | Hospital | Healthcare Workers | Chen (2005a) |
| 39.8 - 100 | | Time | IgG | | | Feb 2003 - Mar 2003 | Hospital | Other | Shi (2005) |
| **55 - 98** | | Other | IgG | | | 5 Mar 2003 - 20 May 2003 | Population | Persons Under Investigation | Liang (2004) |
| **90** | | | IgG | | 390 | 14 Mar 2003 - 22 May 2003 | Population | Persons Under Investigation | Liang (2003) |
| **93** | | | IgG | 70 | 75 | 24 Mar 2003 - 28 Mar 2003 | | | Peiris (2003) |
| **China, Hong Kong SAR, China** | | | | | | | | | |
| 0.14 | | | IgG | | 1453 | Jun 2003 - Jun 2003 | Mixed | General Population | Shi (2005) |
| **Hong Kong SAR, China** | | | | | | | | | |
| **0** | 95% CI: 0-0.6 % | | IFA | 0 | 29 | 22 May 2003 - 31 May 2003 | | Healthcare Workers | Yu (2004a) |
| **0** | | | IFA | 0 | 361 | Sep 2003 - Oct 2003 | Population | Children | Lee (2006a) |
| **0.57** | | | IFA | 2 | 353 | Sep 2003 - Oct 2003 | Population | Children | Lee (2006a) |
| **1.29** | | Symptoms | IFA | 1 | 77 | 14 Mar 2003 - 10 Jun 2003 | Hospital | Children | Leung (2004c) |
| **75.8** | | | IFA | 25 | 33 | 26 Feb 2003 - 31 Mar 2003 | Hospital | Other | Tsang (2003a) |
| **85.1** | | | IFA | 74 | 87 | 26 Feb 2003 - 31 Mar 2003 | Hospital | Other | Tsang (2003a) |
| **88** | | Symptoms | IFA | 44 | 50 | 14 Mar 2003 - 10 Jun 2003 | Hospital | Children | Leung (2004c) |
| 0 | | | IgG | 0 | 192 | | Hospital | Healthcare Workers | Ip (2004) |
| **0** | | | IgG | 0 | 674 | Mar 2003 - May 2003 | Hospital | Healthcare Workers | Chan (2003a) |
| **0.19** | 95% CI: 0.02-0.67 % | | IgG | 2 | 1068 | | Contact | Persons Under Investigation | Leung (2009a) |
| **0.19** | 95% CI: 0.02-0.67 % | | IgG | 2 | 1068 | | Contact | Unspecified | Leung (2004b) |
| 2.3 | | | IgG | 3 | 131 | | Hospital | Healthcare Workers | Ip (2004) |
| **3 - 6.1** | | Age, Occupation, Symptoms | IgG | | | Mar 2003 - May 2003 | Mixed | Mixed Groups | Woo (2004) |
| **5.1** | | | IgG | 29 | 574 | 22 May 2003 - 31 May 2003 | | Healthcare Workers | Yu (2004a) |
| **42** | | | IgG | 8 | 19 | 22 Feb 2003 - 31 May 2003 | Hospital | Other | Chan (2004a) |

continued on next page

continued from previous page

| Seroprevalence (%) | Uncertainty | Disaggregated By | Assay | Number Seropositive | Sample Size | Study Dates | Study Setting | Study Group | Source |
|---|---|---|---|---|---|---|---|---|---|
| **92** | | | IgG | 46 | 50 | 22 Feb 2003 - 31 May 2003 | Hospital | Persons Under Investigation | Chan (2004a) |
| **98.41** | | | IgG | 124 | 126 | 11 Mar 2003 - 25 Mar 2003 | Hospital | Persons Under Investigation | Sung (2004) |
| **100** | | | IgG | | | 4 Mar 2003 - 10 Mar 2003 | Hospital | Persons Under Investigation | Wong (2004) |
| **0.8** | | Age, Occupation, Symptoms | Unspecified | | | Mar 2003 - May 2003 | Mixed | Mixed Groups | Woo (2004) |
| **75.6** | | Time | Unspecified | 34 | 45 | Mar 2003 - | Hospital | Healthcare Workers | Cheng (2005b) |
| **78** | | | Unspecified | | | 26 Feb 2003 - 31 Mar 2003 | Hospital | Mixed Groups | Choi (2003a) |
| **95** | | Symptoms | Unspecified | 19 | 20 | 9 Mar 2003 - 28 Apr 2003 | Hospital | Unspecified | Chueng (2004) |
| **96.6** | | | Unspecified | 171 | 177 | 10 Mar 2003 - 5 Jun 2003 | Hospital | Persons Under Investigation | Chan (2005) |
| **Singapore** | | | | | | | | | |
| **2.2** | | Level of Exposure, Other | Unspecified | 8 | 372 | 22 Apr 2003 - 5 Jun 2003 | Hospital | Healthcare Workers | Ho (2004) |
| 56 | | Symptoms | Unspecified | 45 | 80 | Apr 2003 - Jun 2003 | Hospital | Healthcare Workers | Wilder-Smith (2005) |
| **Taiwan, China** | | | | | | | | | |
| **0.19** | 95% CI: 0.02-0.7 % | Age | IFA | 2 | 1030 | Aug 2003 - Dec 2003 | Community | General Population | Tsai (2008) |
| **0.8** | 95% CI: 0.2-3 % | | IFA | 2 | 238 | 15 Jul 2003 - 10 Aug 2003 | Other | Healthcare Workers | Ko (2004b) |
| **4.7** | | | IFA | 9 | 193 | 30 Mar 2003 - 30 Jun 2003 | Hospital | Healthcare Workers | Chang (2004a) |
| **0** | | | IgG | 0 | 115 | 26 Apr 2003 - 26 May 2003 | Hospital | Healthcare Workers | Liu (2006b) |
| 0.41 | | Occupation, Sex | IgG | 9 | 2197 | 1 Jul 2003 - | Hospital | Healthcare Workers | Wang (2007) |
| 1.02 | | Occupation, Sex | IgG | 9 | 882 | 1 Jul 2003 - | Contact | Healthcare Workers | Wang (2007) |
| **12.04** | 95% CI: 10.11-14.18 % | Age | IgG | 124 | 1030 | Aug 2003 - Dec 2003 | Community | General Population | Tsai (2008) |
| **80** | | | IgG | 12 | 15 | 26 Apr 2003 - 26 May 2003 | Hospital | Healthcare Workers | Liu (2006b) |
| **86.7** | | | IgG | 13 | 15 | 1 May 2003 - 15 Jun 2003 | Hospital | Healthcare Workers | Chiu (2004) |
| **92.7** | | | IgG | 38 | 41 | 8 Mar 2003 - 15 Jun 2003 | Hospital | Persons Under Investigation | Wang (2004a) |
| **3** | | | Unspecified | 20 | 658 | 9 Jun 2003 - 15 Jun 2003 | Hospital | Healthcare Workers | Chen (2006a) |
| 84.8 | | | Unspecified | 28 | 33 | 20 Mar 2003 - 5 Jul 2003 | Hospital | Persons Under Investigation | Chen (2005d) |
| **United States** | | | | | | | | | |
| 0 | | Symptoms | IFA | 0 | 127 | 23 Feb 2003 | Travel | Persons Under Investigation | Vogt (2006) |
| **Vietnam** | | | | | | | | | |
| 2 - 15.5 | | Level of Exposure | Unspecified | | | Oct 2003 - May 2004 | Contact | General Population | Nishiyama (2008) |
| **29** | | Occupation | Unspecified | 36 | 124 | 26 Feb 2003 - 24 Mar 2003 | Hospital | Healthcare Workers | Reynolds (2006) |
| 98.4 | | | Unspecified | 61 | 62 | 26 Feb 2003 - May 2003 | Hospital | Persons Under Investigation | Vu (2004) |

Table B.12: Seroprevalence data grouped by assay type used. Study characteristics are reported for each seroprevalence estimate. Estimates in bold are from high-quality studies (quality assessment score > 0.5).

##### B.4.6 Risk factors

Table B.13 lists all extracted risk factors, including whether they were found to be significant or not significant. These are given in order of risks associated with:

1. Infection
2. Severe disease
3. Probable case
4. Superspreading
5. Other
6. Death
7. Serology
8. Attack rate
9. Incidence
10. Mortality
11. Recovery
12. Incubation period
13. Admission to death delay
14. Admission to discharge delay
15. Incidence rate

| Risk Factor | Significance | Method | Sample Size | Country | Study Dates | Study Setting | Study Group | Source |
|---|---|---|---|---|---|---|---|---|
| **Infection** | | | | | | | | |
| Age, Close Contact, Other | Significant | Not Adjusted | 2139 | Hong Kong SAR; China | 4 Apr 2003 - 10 Jun 2003 | Population | Household Contacts Of Survivors | Lau (2004c) |
| Age, Occupation, Other, Close Contact | Significant | Not Adjusted | 176 | Canada | 25 May 2003 - 31 Oct 2003 | Household | Persons Under Investigation | Wilson-Clark (2006) |
| Age, Occupation, Other, Sex | Significant | Unspecified | 1856 | China | May 2003 - May 2003 | Hospital | Healthcare Workers | Chen (2005a) |
| Age, Other, Sex | Significant | Not Adjusted | 127 | Vietnam | 26 Feb 2003 - 7 Apr 2003 | Hospital | General Population | Nishiura (2005) |
| Close Contact | Significant | | 112 | China; Hong Kong SAR; China | | Contact | Persons Under Investigation | Olsen (2003) |
| Close Contact | Significant | Unspecified | 47 | Hong Kong SAR; China | 4 Mar 2003 - 10 Mar 2003 | Hospital | Healthcare Workers | Wong (2004) |
| Close Contact, Occupation | Significant | Unspecified | 31 | Canada | 23 Mar 2003 - | Hospital | Healthcare Workers | Scales (2003) |
| Close Contact, Occupation, Other | Significant | Not Adjusted | 193 | Vietnam | 26 Feb 2003 - 24 Mar 2003 | Hospital | Healthcare Workers | Reynolds (2006) |
| Close Contact, Occupation, Other | Significant | Not Adjusted | 263 | Singapore | 13 Mar 2003 - 31 May 2003 | Hospital | Persons Under Investigation | Chong (2005) |
| Close Contact, Occupation, Other, Sex | Significant | Unspecified | 658 | Taiwan; China | 9 Jun 2003 - 15 Jun 2003 | Hospital | Healthcare Workers | Chen (2006a) |
| Close Contact, Other | Significant | Adjusted | 86 | Singapore | 1 Mar 2003 - 31 Mar 2003 | Hospital | Healthcare Workers | TELEMAN (2004) |
| Close Contact, Other | Significant | Adjusted | 8662 | Canada | 23 Feb 2003 - 2 Jul 2003 | Population | General Population | Rea (2007) |
| Close Contact, Other | Significant | Adjusted | 443 | China | Apr 2004 - Jun 2004 | Hospital | Healthcare Workers | PEI (2006) |
| Close Contact, Other | Significant | Adjusted | 2139 | Hong Kong SAR; China | 4 Apr 2003 - 10 Jun 2003 | Population | Household Contacts Of Survivors | Lau (2004c) |
| Close Contact, Other | Significant | Adjusted | 477 | China | 5 Mar 2003 - 17 May 2003 | Hospital | General Population | Liu (2009a) |
| Close Contact, Other | Significant | Not Adjusted | 86 | Singapore | 1 Mar 2003 - 31 Mar 2003 | Hospital | Healthcare Workers | TELEMAN (2004) |
| Close Contact, Other | Significant | Not Adjusted | 216 | Hong Kong SAR; China | 28 Mar 2003 - 25 May 2003 | Hospital | Healthcare Workers | Lau (2004d) |
| Close Contact, Other | Significant | Not Adjusted | 477 | China | 5 Mar 2003 - 17 May 2003 | Hospital | General Population | Liu (2009a) |
| Cormobidity, Occupation, Other | Significant | Not Adjusted | 98 | Singapore | 1 Mar 2003 - 31 May 2003 | Hospital | Persons Under Investigation | Chen (2006e) |
| Hospitalisation, Household Contact, Non-household Contact, Other, Social gathering | Significant | Unspecified | 225 | Canada | 23 Feb 2003 - 1 Jul 2003 | Hospital | Unspecified | Svoboda (2004) |
| Hospitalisation, Occupation, Other | Significant | Adjusted | 85 | Vietnam | Oct 2003 - May 2004 | Contact | General Population | Nishiyama (2008) |
| Household Contact | Significant | Unspecified | 16 | Hong Kong SAR; China | 13 Mar 2003 - 17 May 2003 | Hospital | Persons Under Investigation | Cheng (2005c) |
| Household Contact, Other | Significant | Adjusted | 212 | Vietnam | 26 Feb 2003 - 28 Apr 2003 | Contact | Mixed Groups | Tuan (2007) |
| Household Contact, Other | Significant | Not Adjusted | 212 | Vietnam | 26 Feb 2003 - 28 Apr 2003 | Contact | Mixed Groups | Tuan (2007) |
| Occupation, Other | Significant | Adjusted | 748 | China | May 2003 - May 2003 | Hospital | Healthcare Workers | Chen (2009b) |
| Occupation, Other | Significant | Unspecified | 32 | Canada | 8 Mar 2003 - 3 Apr 2003 | Hospital | Healthcare Workers | Loeb (2004) |
| Other | Significant | Adjusted | 74 | Hong Kong SAR; China | 4 Mar 2003 - Mar 2003 | Hospital | Persons Under Investigation | Yu (2005) |
| Other | Significant | Adjusted | 187 | Hong Kong SAR; China | 21 Mar 2003 - 1 Apr 2003 | Community | Persons Under Investigation | Yu (2004b) |
| Other | Significant | Adjusted | 254 | Hong Kong SAR; China | 15 Mar 2003 - 24 Mar 2003 | Hospital | Healthcare Workers | Seto (2003) |

continued on next page

continued from previous page

| Risk Factor | Occupation | Method | Sample Size | Country | Study Dates | Study Setting | Study Group | Source |
|---|---|---|---|---|---|---|---|---|
| Other | Significant | Adjusted | 624 | Canada | 5 Mar 2003 - 12 Jun 2003 | Hospital | Healthcare Workers | Raboud (2010) |
| Other | Significant | Adjusted | 330 | Hong Kong SAR; China | 4 Apr 2003 - 10 Jun 2003 | Population | General Population | Lau (2004b) |
| Other | Significant | Adjusted | 216 | Hong Kong SAR; China | 28 Mar 2003 - 25 May 2003 | Hospital | Healthcare Workers | Lau (2004d) |
| Other | Significant | Adjusted | 98 | Singapore | 1 Mar 2003 - 31 May 2003 | Hospital | Persons Under Investigation | Chen (2006e) |
| Other | Significant | Not Adjusted | 624 | Canada | 5 Mar 2003 - 12 Jun 2003 | Hospital | Healthcare Workers | Raboud (2010) |
| Other | Significant | Not Adjusted | 330 | Hong Kong SAR; China | 4 Apr 2003 - 10 Jun 2003 | Population | General Population | Lau (2004b) |
| Other | Significant | Not Adjusted | 37174 | Hong Kong SAR; China | 17 Apr 2003 - 17 Aug 2003 | Hospital | Healthcare Workers | Lau (2005) |
| Other | Significant | Not Adjusted | 86 | Singapore | 1 Mar 2003 - 22 Mar 2003 | Hospital | Healthcare Workers | Leung (2004c) |
| Other | Significant | Not Adjusted | 76 | Canada | 1 Apr 2003 - 22 Apr 2003 | Hospital | Persons Under Investigation | Fowler (2004) |
| Other | Significant | Unspecified | 662 | China | 16 Nov 2002 - 30 Apr 2003 | Community | General Population | Xu (2004) |
| Other | Significant | Unspecified | 1018 | Singapore | 13 Mar 2003 - 31 May 2003 | Hospital | Persons Under Investigation | Tham (2004) |
| Other | Significant | Unspecified | | China | 1 Mar 2003 - 16 Aug 2003 | Population | General Population | Liu (2005) |
| Other, Close Contact | Significant | Adjusted | 176 | Canada | 25 May 2003 - 31 Oct 2003 | Household | Persons Under Investigation | Wilson-Clark (2006) |
| Age, Close Contact, Cormobidity, Household Contact, Non-household Contact, Other, Sex | Not Significant | Not Adjusted | 212 | Vietnam | 26 Feb 2003 - 28 Apr 2003 | Contact | Mixed Groups | Tuan (2007) |
| Age, Close Contact, Cormobidity, Occupation, Other | Not Significant | Not Adjusted | 86 | Singapore | 1 Mar 2003 - 31 Mar 2003 | Hospital | Healthcare Workers | TELEMAN (2004) |
| Age, Cormobidity, Other | Not Significant | Adjusted | 74 | Hong Kong SAR; China | 4 Mar 2003 - Mar 2003 | Hospital | Persons Under Investigation | Yu (2005) |
| Age, Cormobidity, Other | Not Significant | Adjusted | 85 | Vietnam | Oct 2003 - May 2004 | Hospital | General Population | Nishiyama (2008) |
| Age, Cormobidity, Other, Sex | Not Significant | Not Adjusted | 624 | Canada | 5 Mar 2003 - 12 Jun 2003 | Hospital | Healthcare Workers | Raboud (2010) |
| Age, Occupation | Not Significant | Not Adjusted | 176 | Canada | 25 May 2003 - 31 Oct 2003 | Household | Persons Under Investigation | Wilson-Clark (2006) |
| Age, Occupation, Other | Not Significant | Not Adjusted | 127 | Vietnam | 26 Feb 2003 - 7 Apr 2003 | Hospital | General Population | Nishiura (2005) |
| Age, Other, Sex | Not Significant | Adjusted | 2139 | Hong Kong SAR; China | 4 Apr 2003 - 10 Jun 2003 | Population | Household Contacts Of Survivors | Lau (2004c) |
| Age, Other, Sex | Not Significant | Not Adjusted | 98 | Singapore | 1 Mar 2003 - 31 May 2003 | Hospital | Persons Under Investigation | Chen (2006e) |
| Age, Sex | Not Significant | Adjusted | 8662 | Canada | 23 Feb 2003 - 2 Jul 2003 | Population | General Population | Rea (2007) |
| Close Contact, Occupation | Not Significant | Not Adjusted | 193 | Vietnam | 26 Feb 2003 - 24 Mar 2003 | Hospital | Healthcare Workers | Reynolds (2006) |
| Close Contact, Other | Not Significant | Not Adjusted | 330 | Hong Kong SAR; China | 4 Apr 2003 - 10 Jun 2003 | Population | General Population | Lau (2004b) |
| Close Contact, Other | Not Significant | Not Adjusted | 216 | Hong Kong SAR; China | 28 Mar 2003 - 25 May 2003 | Hospital | Healthcare Workers | Lau (2004d) |
| Close Contact, Other | Not Significant | Not Adjusted | 477 | China | 5 Mar 2003 - 17 May 2003 | Hospital | General Population | Liu (2009a) |
| Hospitalisation, Household Contact, Non-household Contact, Other, Social gathering | Not Significant | Unspecified | 225 | Canada | 23 Feb 2003 - 1 Jul 2003 | Hospital | Unspecified | Svoboda (2004) |

continued on next page

continued from previous page

| Risk Factor | Occupation | Method | Sample Size | Country | Study Dates | Study Setting | Study Group | Source |
|---|---|---|---|---|---|---|---|---|
| Occupation, Other | Not Significant | Adjusted | 176 | Canada | 25 May 2003 - 31 Oct 2003 | Household | Persons Under Investigation | Wilson-Clark (2006) |
| Other | Not Significant | Adjusted | 187 | Hong Kong SAR; China | 21 Mar 2003 - 1 Apr 2003 | Community | Persons Under Investigation | Yu (2004b) |
| Other | Not Significant | Adjusted | 216 | Hong Kong SAR; China | 28 Mar 2003 - 25 May 2003 | Hospital | Healthcare Workers | Lau (2004d) |
| Other | Not Significant | Adjusted | 477 | China | 5 Mar 2003 - 17 May 2003 | Hospital | General Population | Liu (2009a) |
| Other | Not Significant | Not Adjusted | 47 | Hong Kong SAR; China | 4 Mar 2003 - 10 Mar 2003 | Hospital | Healthcare Workers | Wong (2004) |
| Other | Not Significant | Not Adjusted | 110 | China | 15 Mar 2003 - | Travel | General Population | Lei (2018) |
| Other | Not Significant | Unspecified | 32 | Canada | 8 Mar 2003 - 3 Apr 2003 | Hospital | Healthcare Workers | Loeb (2004) |
| Other, Sex | Not Significant | Adjusted | 86 | Singapore | 1 Mar 2003 - 31 Mar 2003 | Hospital | Healthcare Workers | TELEMAN (2004) |
| Other, Sex | Not Significant | Not Adjusted | 2139 | Hong Kong SAR; China | 4 Apr 2003 - 10 Jun 2003 | Population | Household Contacts Of Survivors | Lau (2004c) |
| Close Contact, Household Contact, Non-household Contact, Occupation | Unspecified | Unspecified | 1112 | China | 2003 | Community | Persons Under Investigation | Zeng (2009) |
| Occupation | Unspecified | Not Adjusted | 182 | Hong Kong SAR; China | 13 Mar 2003 - 31 May 2003 | Hospital | Healthcare Workers | Gomersall (2006) |
| Occupation | Unspecified | Unspecified | 321 | Taiwan; China | 18 Mar 2003 - 19 Jun 2003 | Trade | Healthcare Workers | Ko (2004b) |
| **Severe disease** | | | | | | | | |
| Age | Significant | Not Adjusted | 44 | Hong Kong SAR; China | 14 Mar 2003 - 10 Jun 2003 | Hospital | Children | Leung (2004c) |
| Age | Significant | Unspecified | 43 | Hong Kong SAR; China | 14 Mar 2003 - 10 Jun 2003 | Hospital | Children | Leung (2004c) |
| Age, Cormobidity | Significant | Adjusted | 78 | China | 22 Dec 2002 - Mar 2003 | Hospital | General Population | Xiao (2003) |
| Age, Cormobidity, Other | Significant | Adjusted | 67 | Taiwan; China | 20 Mar 2003 - 5 Jul 2003 | Hospital | Persons Under Investigation | Chen (2005d) |
| Age, Cormobidity, Other, Sex | Significant | Not Adjusted | 234 | Singapore | | Hospital | Mixed Groups | Leong (2006a) |
| Age, Cormobidity, Other, Sex | Significant | Not Adjusted | 67 | Taiwan; China | 20 Mar 2003 - 5 Jul 2003 | Hospital | Persons Under Investigation | Chen (2005d) |
| Age, Cormobidity, Other, Sex | Significant | Not Adjusted | 201 | Hong Kong SAR; China | 26 Feb 2003 - 10 Apr 2003 | Hospital | General Population | Chau (2004) |
| Age, Cormobidity, Other, Sex | Significant | Not Adjusted | 144 | Canada | 7 Mar 2003 - 10 Apr 2003 | Hospital | General Population | Booth (2003) |
| Age, Other | Significant | Adjusted | 234 | Singapore | | Hospital | Mixed Groups | Leong (2006a) |
| Cormobidity | Significant | Adjusted | 144 | Canada | 7 Mar 2003 - 10 Apr 2003 | Hospital | General Population | Booth (2003) |
| Other | Significant | Adjusted | 44 | Hong Kong SAR; China | 14 Mar 2003 - 10 Jun 2003 | Hospital | Children | Leung (2004c) |
| Other | Significant | Adjusted | 225 | China | Feb 2003 - May 2003 | Population | General Population | Chang (2006b) |
| Other | Significant | Not Adjusted | 29 | Taiwan; China | 28 Mar 2003 - 30 Jun 2003 | Hospital | Persons Under Investigation | Jang (2004) |
| Age | Not Significant | Adjusted | 144 | Canada | 7 Mar 2003 - 10 Apr 2003 | Hospital | General Population | Booth (2003) |
| Age | Not Significant | Unspecified | 44 | Hong Kong SAR; China | 14 Mar 2003 - 10 Jun 2003 | Hospital | Children | Leung (2004c) |
| Age, Cormobidity, Household Contact, Non-household Contact, Other, Sex | Not Significant | Not Adjusted | 29 | Taiwan; China | 28 Mar 2003 - 30 Jun 2003 | Hospital | Persons Under Investigation | Jang (2004) |

continued on next page

continued from previous page

| Risk Factor | Occupation | Method | Sample Size | Country | Study Dates | Study Setting | Study Group | Source |
|---|---|---|---|---|---|---|---|---|
| Other | Not Significant | Adjusted | 225 | China | Feb 2003 - May 2003 | Population | General Population | Chang (2006b) |
| Other | Not Significant | Not Adjusted | 234 | Singapore | | Hospital | Mixed Groups | Leong (2006a) |
| Other | Not Significant | Not Adjusted | 67 | Taiwan; China | 20 Mar 2003 - 5 Jul 2003 | Hospital | Persons Under Investigation | Chen (2005d) |
| Other | Not Significant | Not Adjusted | 201 | Hong Kong SAR; China | 26 Feb 2003 - 10 Apr 2003 | Hospital | General Population | Chau (2004) |
| Other | Not Significant | Not Adjusted | 144 | Canada | 7 Mar 2003 - 10 Apr 2003 | Hospital | General Population | Booth (2003) |
| **Probable case** | | | | | | | | |
| Contact with Animal, Cormobidity, Other | Significant | Adjusted | 375 | China | 28 Apr 2003 - 4 Jul 2003 | Population | General Population | Wu (2004) |
| Cormobidity, Other | Significant | Not Adjusted | 375 | China | 28 Apr 2003 - 4 Jul 2003 | Population | General Population | Wu (2004) |
| Contact with Animal, Other | Not Significant | Not Adjusted | 375 | China | 28 Apr 2003 - 4 Jul 2003 | Population | General Population | Wu (2004) |
| Other | Not Significant | Adjusted | 375 | China | 28 Apr 2003 - 4 Jul 2003 | Population | General Population | Wu (2004) |
| **Superspreading** | | | | | | | | |
| Close Contact | Significant | Not Adjusted | 77 | China | 5 Feb 2003 - 4 May 2003 | Hospital | Persons Under Investigation | Shen (2004) |
| Close Contact, Other | Significant | Adjusted | 124 | China; Hong Kong SAR; China | Sep 2004 - Nov 2005 | Hospital | General Population | Yu (2007) |
| Close Contact, Other | Significant | Adjusted | 127 | China | Sep 2004 - Nov 2005 | Hospital | General Population | Sung (2009) |
| Close Contact, Other | Significant | Not Adjusted | 124 | China; Hong Kong SAR; China | Sep 2004 - Nov 2005 | Hospital | General Population | Yu (2007) |
| Close Contact, Other | Significant | Not Adjusted | 127 | China | Sep 2004 - Nov 2005 | Hospital | General Population | Sung (2009) |
| Cormobidity, Other | Significant | Not Adjusted | 12 | Singapore | 1 Mar 2003 - 31 May 2003 | Hospital | Persons Under Investigation | Chen (2006d) |
| Age, Occupation, Other, Sex | Not Significant | Not Adjusted | 12 | Singapore | 1 Mar 2003 - 31 May 2003 | Hospital | Persons Under Investigation | Chen (2006d) |
| Cormobidity, Other | Not Significant | Not Adjusted | 124 | China; Hong Kong SAR; China | Sep 2004 - Nov 2005 | Hospital | General Population | Yu (2007) |
| Other | Not Significant | Adjusted | 124 | China; Hong Kong SAR; China | Sep 2004 - Nov 2005 | Hospital | General Population | Yu (2007) |
| Other | Not Significant | Adjusted | 127 | China | Sep 2004 - Nov 2005 | Hospital | General Population | Sung (2009) |
| Other | Not Significant | Not Adjusted | 127 | China | Sep 2004 - Nov 2005 | Hospital | General Population | Sung (2009) |
| Sex | Not Significant | Not Adjusted | 77 | China | 5 Feb 2003 - 4 May 2003 | Hospital | Persons Under Investigation | Shen (2004) |
| **Other** | | | | | | | | |
| Age | Significant | Unspecified | 44 | Hong Kong SAR; China | 14 Mar 2003 - 10 Jun 2003 | Hospital | Children | Leung (2004c) |
| Age | Significant | Unspecified | 44 | Hong Kong SAR; China | 14 Mar 2003 - 10 Jun 2003 | Hospital | Children | Leung (2004c) |
| Age, Cormobidity, Other, Sex | Significant | Not Adjusted | 76 | Taiwan; China | 8 Mar 2003 - 15 Jun 2003 | Hospital | Persons Under Investigation | Wang (2004a) |
| Age, Cormobidity, Other, Sex | Significant | Not Adjusted | 303 | Hong Kong SAR; China | 1 Jun 2004 - 31 Dec 2004 | Hospital | Persons Under Investigation | Lee (2006b) |
| Age, Occupation | Significant | Adjusted | 417 | Singapore | 24 Feb 2003 - 29 Apr 2003 | Household | Other | Goh (2004) |

continued on next page

continued from previous page

| Risk Factor | Occupation | Method | Sample Size | Country | Study Dates | Study Setting | Study Group | Source |
|---|---|---|---|---|---|---|---|---|
| Age, Occupation | Significant | Not Adjusted | 417 | Singapore | 24 Feb 2003 - 29 Apr 2003 | Household | Other | Goh (2004) |
| Age, Other | Significant | Adjusted | 76 | Taiwan; China | 8 Mar 2003 - 15 Jun 2003 | Hospital | Persons Under Investigation | Wang (2004a) |
| Close Contact, Other | Significant | Not Adjusted | 273 | Canada | Feb 2003 - Jun 2003 | Hospital | Persons Under Investigation | Muller (2006) |
| Other | Significant | Adjusted | | Hong Kong SAR; China | 11 Mar 2003 - 22 May 2003 | Hospital | Healthcare Workers | Lin (2006) |
| Other | Significant | Adjusted | | Hong Kong SAR; China | 11 Mar 2003 - 22 May 2003 | Housing | General Population | Lin (2006) |
| Other | Significant | Adjusted | | Hong Kong SAR; China | 11 Mar 2003 - 22 May 2003 | Community | General Population | Lin (2006) |
| Other | Significant | Adjusted | 350 | China | 1 Jan 2003 - 31 May 2003 | Mixed | Persons Under Investigation | Cai (2007) |
| Other | Significant | Not Adjusted | 45 | Singapore | Apr 2003 - Jun 2003 | Hospital | Healthcare Workers | Wilder-Smith (2005) |
| Other | Significant | Not Adjusted | 138 | Hong Kong SAR; China | 11 Mar 2003 - 25 Mar 2003 | Hospital | Persons Under Investigation | Sung (2004) |
| Other | Significant | Not Adjusted | 209 | China | 2003 | Mixed | Persons Under Investigation | Cai (2006) |
| Other | Significant | Not Adjusted | 350 | China | 1 Jan 2003 - 31 May 2003 | Mixed | Persons Under Investigation | Cai (2007) |
| Other | Significant | Unspecified | 8378 | Singapore | 13 Mar 2003 - 31 May 2003 | Hospital | Persons Under Investigation | Tham (2004) |
| Other | Significant | Unspecified | 45 | Hong Kong SAR; China | 24 Mar 2003 - 4 May 2003 | Hospital | Persons Under Investigation | Chu (2005) |
| Age | Not Significant | Unspecified | 44 | Hong Kong SAR; China | 14 Mar 2003 - 10 Jun 2003 | Hospital | Children | Leung (2004c) |
| Age, Cormobidity, Other, Sex | Not Significant | Unspecified | 45 | Hong Kong SAR; China | 24 Mar 2003 - 4 May 2003 | Hospital | Persons Under Investigation | Chu (2005) |
| Age, Non-household Contact, Other, Sex | Not Significant | Not Adjusted | 45 | Singapore | Apr 2003 - Jun 2003 | Hospital | Healthcare Workers | Wilder-Smith (2005) |
| Age, Occupation, Other, Sex | Not Significant | Not Adjusted | 273 | Canada | Feb 2003 - Jun 2003 | Hospital | Persons Under Investigation | Muller (2006) |
| Age, Occupation, Other, Sex | Not Significant | Not Adjusted | 209 | China | 2003 | Mixed | Persons Under Investigation | Cai (2006) |
| Age, Other, Sex | Not Significant | Not Adjusted | 138 | Hong Kong SAR; China | 11 Mar 2003 - 25 Mar 2003 | Hospital | Persons Under Investigation | Sung (2004) |
| Age, Other, Sex | Not Significant | Not Adjusted | 417 | Singapore | 24 Feb 2003 - 29 Apr 2003 | Household | Other | Goh (2004) |
| Cormobidity, Other, Sex | Not Significant | Adjusted | 76 | Taiwan; China | 8 Mar 2003 - 15 Jun 2003 | Hospital | Persons Under Investigation | Wang (2004a) |
| Other | Not Significant | Adjusted | 350 | China | 1 Jan 2003 - 31 May 2003 | Mixed | Persons Under Investigation | Cai (2007) |
| Other | Not Significant | Not Adjusted | 76 | Taiwan; China | 8 Mar 2003 - 15 Jun 2003 | Hospital | Persons Under Investigation | Wang (2004a) |
| Other | Not Significant | Not Adjusted | 350 | China | 1 Jan 2003 - 31 May 2003 | Mixed | Persons Under Investigation | Cai (2007) |
| Hospitalisation | Unspecified | Unspecified | 1512 | Hong Kong SAR; China | 3 Mar 2003 - 6 May 2003 | Mixed | Mixed Groups | Riley (2003) |
| **Death** | | | | | | | | |
| Age | Significant | Adjusted | 46 | Taiwan; China | 18 Apr 2003 - 31 May 2003 | Hospital | Persons Under Investigation | Lim (2003) |
| Age | Significant | Adjusted | 138 | Hong Kong SAR; China | 11 Mar 2003 - 25 Mar 2003 | Hospital | Persons Under Investigation | Lee (2003) |
| Age | Significant | Adjusted | 267 | Hong Kong SAR; China | 26 Feb 2003 - 31 Mar 2003 | Hospital | Mixed Groups | Choi (2003a) |

continued on next page

continued from previous page

| Risk Factor | Occupation | Method | Sample Size | Country | Study Dates | Study Setting | Study Group | Source |
|---|---|---|---|---|---|---|---|---|
| Age | Significant | Not Adjusted | 77 | Hong Kong SAR; China | 22 Feb 2003 - 31 May 2003 | Hospital | Persons Under Investigation | Chan (2004a) |
| Age | Significant | Unspecified | 2444 | China | 5 Mar 2003 - 20 May 2003 | Hospital | Persons Under Investigation | Liang (2004) |
| Age | Significant | Unspecified | 345 | Taiwan; China | - 12 Oct 2003 | Hospital | Persons Under Investigation | Chang (2007) |
| Age, Close Contact, Cormobidity, Other, Sex | Significant | Not Adjusted | 1312 | Hong Kong SAR; China | 2003 | Hospital | General Population | Chan (2007) |
| Age, Close Contact, Hospitalisation, Other | Significant | Not Adjusted | 2521 | China | Mar 2003 - 16 Aug 2003 | Mixed | Persons Under Investigation | Chen (2005b) |
| Age, Close Contact, Other, Sex | Significant | Adjusted | 1312 | Hong Kong SAR; China | 2003 | Hospital | General Population | Chan (2007) |
| Age, Cormobidity, Hospitalisation, Occupation, Other | Significant | Not Adjusted | 1649 | China | 2003 - 7 Jul 2003 | Population | Persons Under Investigation | Liu (2006a) |
| Age, Cormobidity, Occupation, Other | Significant | Adjusted | 3336 | China; Hong Kong SAR; China; Taiwan; China | | Population | General Population | Lau (2010) |
| Age, Cormobidity, Occupation, Other, Sex | Significant | Adjusted | 1755 | Hong Kong SAR; China | | Population | Persons Under Investigation | Leung (2009b) |
| Age, Cormobidity, Occupation, Other, Sex | Significant | Not Adjusted | 90 | China | 2003 | Hospital | Mixed Groups | Wei (2009) |
| Age, Cormobidity, Occupation, Other, Sex | Significant | Not Adjusted | 346 | Taiwan; China | | Population | General Population | Chang (2006a) |
| Age, Cormobidity, Other | Significant | Not Adjusted | 76 | Taiwan; China | 8 Mar 2003 - 15 Jun 2003 | Hospital | Persons Under Investigation | Wang (2004a) |
| Age, Cormobidity, Other | Significant | Not Adjusted | 38 | Canada | - 15 Apr 2003 | Hospital | Persons Under Investigation | Fowler (2003) |
| Age, Cormobidity, Other, Sex | Significant | Adjusted | | Canada; Hong Kong SAR; China | | Hospital | Persons Under Investigation | Cowling (2006) |
| Age, Cormobidity, Other, Sex | Significant | Not Adjusted | 218 | Hong Kong SAR; China | 26 Feb 2003 - 31 Mar 2003 | Hospital | Persons Under Investigation | Tsang (2003a) |
| Age, Cormobidity, Other, Sex | Significant | Not Adjusted | 234 | Singapore | | Hospital | Mixed Groups | Leong (2006a) |
| Age, Cormobidity, Other, Sex | Significant | Not Adjusted | 668 | Taiwan; China | 14 Mar 2003 - 30 Jul 2003 | Hospital | Persons Under Investigation | Chen (2005c) |
| Age, Hospitalisation, Occupation, Other | Significant | Adjusted | 1649 | China | 2003 - 7 Jul 2003 | Population | Persons Under Investigation | Liu (2006a) |
| Age, Occupation | Significant | Adjusted | | China | 3 Mar 2003 - | Hospital | Mixed Groups | Cooper (2009) |
| Age, Occupation, Other | Significant | Unspecified | 78 | Taiwan; China | Aug 2003 - Feb 2004 | Hospital | Persons Under Investigation | Liu (2009b) |
| Age, Occupation, Other, Sex | Significant | Not Adjusted | 5327 | China | 16 Nov 2002 - 2003 | Population | General Population | Na (2009) |
| Age, Occupation, Sex | Significant | Not Adjusted | | China | 3 Mar 2003 - | Hospital | Mixed Groups | Cooper (2009) |
| Age, Other | Significant | Adjusted | 82 | China | 2003 | Hospital | Mixed Groups | Wei (2009) |
| Age, Other | Significant | Adjusted | 5327 | China | 16 Nov 2002 - 2003 | Population | General Population | Na (2009) |
| Age, Other | Significant | Adjusted | 234 | Singapore | | Hospital | Mixed Groups | Leong (2006a) |
| Age, Other | Significant | Adjusted | 401 | China | Dec 2002 - Jun 2003 | Hospital | Persons Under Investigation | Chen (2006c) |
| Age, Other | Significant | Adjusted | 115 | Hong Kong SAR; China | 9 Mar 2003 - 31 May 2003 | Hospital | General Population | Chan (2003c) |
| Age, Other | Significant | Adjusted | 346 | Taiwan; China | | Population | General Population | Chang (2006a) |
| Age, Other | Significant | Not Adjusted | 77 | China | 5 Feb 2003 - 4 May 2003 | Hospital | Persons Under Investigation | Shen (2004) |

continued on next page

continued from previous page

| Risk Factor | Occupation | Method | Sample Size | Country | Study Dates | Study Setting | Study Group | Source |
|---|---|---|---|---|---|---|---|---|
| Age, Other | Significant | Not Adjusted | 46 | Taiwan; China | 18 Apr 2003 - 31 May 2003 | Hospital | Persons Under Investigation | Lim (2003) |
| Age, Other | Significant | Not Adjusted | 115 | Hong Kong SAR; China | 9 Mar 2003 - 31 May 2003 | Hospital | General Population | Chan (2003c) |
| Age, Other | Significant | Unspecified | 52 | Taiwan; China | 26 Apr 2003 - 25 May 2003 | Hospital | Persons Under Investigation | Ko (2004a) |
| Age, Sex | Significant | Adjusted | 1755 | Hong Kong SAR; China | Mar 2003 - 22 Sep 2003 | Hospital | Persons Under Investigation | Karlberg (2004) |
| Age, Sex | Significant | Not Adjusted | 138 | Hong Kong SAR; China | 11 Mar 2003 - 25 Mar 2004 | Hospital | Persons Under Investigation | Lee (2003) |
| Cormobidity, Other | Significant | Adjusted | 76 | Taiwan; China | 8 Mar 2003 - 15 Jun 2003 | Hospital | Persons Under Investigation | Wang (2004a) |
| Cormobidity, Other | Significant | Adjusted | 218 | Hong Kong SAR; China | 26 Feb 2003 - 31 Mar 2003 | Hospital | Persons Under Investigation | Tsang (2003a) |
| Cormobidity, Other, Sex | Significant | Adjusted | 1755 | Hong Kong SAR; China | 15 Feb 2003 - 31 May 2003 | Population | Persons Under Investigation | Leung (2004a) |
| Other | Significant | Adjusted | 308 | Hong Kong SAR; China | Feb 2003 - Jul 2003 | Hospital | Persons Under Investigation | Virlogeux (2015) |
| Other | Significant | Adjusted | 234 | Hong Kong SAR; China | Feb 2003 - Jul 2003 | Hospital | Persons Under Investigation | Virlogeux (2015) |
| Other | Significant | Adjusted | 1632 | China | 15 Mar 2003 - 14 May 2003 | Population | General Population | Zhang (2006) |
| Other | Significant | Not Adjusted | 5327 | China | Apr 2003 - May 2003 | Population | Persons Under Investigation | Cui (2003) |
| Other | Significant | Not Adjusted | 678 | Hong Kong SAR; China | 16 Apr 2003 - | Hospital | General Population | Chan (2003b) |
| Other | Significant | Not Adjusted | 1373 | Hong Kong SAR; China | Mar 2003 - Jul 2003 | Hospital | General Population | Antonio (2005) |
| Other | Significant | Unspecified | | | | | | Ho (2005) |
| Sex | Significant | Not Adjusted | 1755 | Hong Kong SAR; China | Mar 2003 - 22 Sep 2003 | Hospital | Persons Under Investigation | Karlberg (2004) |
| Sex, Age, Occupation | Significant | Not Adjusted | 1755 | Hong Kong SAR; China | Feb 2003 - Jul 2003 | Hospital | Mixed Groups | Virlogeux (2015) |
| Age, Occupation, Other | Not Significant | Adjusted | 1755 | Hong Kong SAR; China | | Population | Persons Under Investigation | Leung (2009b) |
| Age, Other | Not Significant | Not Adjusted | 1649 | China | 2003 - 7 Jul 2003 | Population | Persons Under Investigation | Liu (2006a) |
| Age, Other, Sex | Not Significant | Adjusted | 76 | Taiwan; China | 8 Mar 2003 - 15 Jun 2003 | Hospital | Persons Under Investigation | Wang (2004a) |
| Age, Other, Sex | Not Significant | Adjusted | 346 | Taiwan; China | | Population | General Population | Chang (2006a) |
| Close Contact, Other, Sex | Not Significant | Adjusted | 1312 | Hong Kong SAR; China | 2003 | Hospital | General Population | Chan (2007) |
| Cormobidity, Occupation, Other, Sex | Not Significant | Adjusted | 82 | China | 2003 | Hospital | Mixed Groups | Wei (2009) |
| Occupation, Other | Not Significant | Not Adjusted | 5327 | China | 16 Nov 2002 - 2003 | Population | General Population | Na (2009) |
| Occupation, Other, Sex | Not Significant | Not Adjusted | 38 | Canada | - 15 Apr 2003 | Hospital | Persons Under Investigation | Fowler (2003) |
| Occupation, Sex | Not Significant | Adjusted | 5327 | China | 16 Nov 2002 - 2003 | Population | General Population | Na (2009) |
| Other | Not Significant | Adjusted | 46 | Taiwan; China | 18 Apr 2003 - 31 May 2003 | Hospital | Persons Under Investigation | Lim (2003) |
| Other | Not Significant | Adjusted | 1755 | Hong Kong SAR; China | Mar 2003 - 22 Sep 2003 | Hospital | Persons Under Investigation | Karlberg (2004) |
| Other | Not Significant | Adjusted | | Canada; Hong Kong SAR; China | | Hospital | Persons Under Investigation | Cowling (2006) |

continued on next page

continued from previous page

| Risk Factor | Occupation | Method | Sample Size | Country | Study Dates | Study Setting | Study Group | Source |
|---|---|---|---|---|---|---|---|---|
| Other | Not Significant | Adjusted | 1632 | China | 15 Mar 2003 - 14 May 2003 | Population | General Population | Zhang (2006) |
| Other | Not Significant | Not Adjusted | 1755 | Hong Kong SAR; China | Feb 2003 - Jul 2003 | Hospital | Mixed Groups | Virlogeux (2015) |
| Other | Not Significant | Not Adjusted | 90 | China | 2003 | Hospital | Mixed Groups | Wei (2009) |
| Other | Not Significant | Not Adjusted | 46 | Taiwan; China | 18 Apr 2003 - 31 May 2003 | Hospital | Persons Under Investigation | Lim (2003) |
| Other | Not Significant | Not Adjusted | 1743 | Hong Kong SAR; China | | Population | General Population | Lau (2009) |
| Other | Not Significant | Not Adjusted | 234 | Singapore | | Hospital | Mixed Groups | Leong (2006a) |
| Other | Not Significant | Not Adjusted | 374 | Hong Kong SAR; China | 16 Apr 2003 - | Hospital | General Population | Chan (2003b) |
| Other | Not Significant | Not Adjusted | 1312 | Hong Kong SAR; China | 2003 | Hospital | General Population | Chan (2007) |
| Other | Not Significant | Not Adjusted | 346 | Taiwan; China | | Population | General Population | Chang (2006a) |
| Other | Not Significant | Not Adjusted | 1373 | Hong Kong SAR; China | Mar 2003 - Jul 2003 | Hospital | General Population | Antonio (2005) |
| Other | Not Significant | Not Adjusted | | China | 3 Mar 2003 - | Hospital | Mixed Groups | Cooper (2009) |
| Other, Sex | Not Significant | Adjusted | | China | 3 Mar 2003 - | Hospital | Mixed Groups | Cooper (2009) |
| Other, Sex | Not Significant | Not Adjusted | 76 | Taiwan; China | 8 Mar 2003 - 15 Jun 2003 | Hospital | Persons Under Investigation | Wang (2004a) |
| Other, Sex | Not Significant | Not Adjusted | 115 | Hong Kong SAR; China | 9 Mar 2003 - 31 May 2003 | Hospital | General Population | Chan (2003c) |
| Sex | Not Significant | Adjusted | 3336 | China; Hong Kong SAR; China; Taiwan; China | | Population | General Population | Lau (2010) |
| Sex | Not Significant | Adjusted | 138 | Hong Kong SAR; China | 11 Mar 2003 - 25 Mar 2003 | Hospital | Persons Under Investigation | Lee (2003) |
| Sex | Not Significant | Not Adjusted | 2521 | China | Mar 2003 - 16 Aug 2003 | Mixed | Persons Under Investigation | Chen (2005b) |
| Sex | Not Significant | Unspecified | 52 | Taiwan; China | 26 Apr 2003 - 25 May 2003 | Hospital | Persons Under Investigation | Ko (2004a) |
| Age, Occupation, Other | Unspecified | Adjusted | 1755 | Hong Kong SAR; China | 15 Feb 2003 - 31 May 2003 | Population | Persons Under Investigation | Leung (2004a) |
| Cormobidity, Other | Unspecified | Unspecified | 8 | Taiwan; China | 26 Mar 2003 - 25 May 2003 | Hospital | Persons Under Investigation | Wong (2003) |
| Occupation | Unspecified | Unspecified | 405 | Hong Kong SAR; China | 15 Feb 2003 - 31 May 2003 | Hospital | Persons Under Investigation | Leung (2004a) |
| **Serology** | | | | | | | | |
| Age | Significant | Not Adjusted | | Hong Kong SAR; China | 22 Feb 2003 - 31 May 2003 | Hospital | Persons Under Investigation | Chan (2004a) |
| Age | Significant | Unspecified | 375 | Hong Kong SAR; China | 4 Mar 2003 - 6 Jun 2003 | Hospital | General Population | Chan (2004c) |
| Occupation | Significant | Not Adjusted | 2197 | Taiwan; China | 1 Jul 2003 - | Hospital | Healthcare Workers | Wang (2007) |
| Occupation | Significant | Not Adjusted | 882 | Taiwan; China | 1 Jul 2003 - | Contact | Healthcare Workers | Wang (2007) |
| Other | Significant | Adjusted | | | | | | Ho (2005) |
| Age, Sex | Not Significant | Adjusted | 665 | Taiwan; China | Mar 2003 - Jul 2003 | Community | Persons Under Investigation | Ho (2005) |
| Occupation, Sex | Not Significant | Not Adjusted | 2197 | Taiwan; China | 1 Jul 2003 - | Hospital | Healthcare Workers | Wang (2007) |

continued on next page

continued from previous page

| Risk Factor | Occupation | Method | Sample Size | Country | Study Dates | Study Setting | Study Group | Source |
|---|---|---|---|---|---|---|---|---|
| Occupation, Sex | Not Significant | Not Adjusted | 882 | Taiwan; China | 1 Jul 2003 - | Contact | Healthcare Workers | Wang (2007) |
| Other | Not Significant | Not Adjusted | | Hong Kong SAR; China | Sep 2003 - Oct 2003 | Population | Children | Lee (2006a) |
| **Attack Rate** | | | | | | | | |
| Age, Sex | Significant | Unspecified | | China | 5 Mar 2003 - 20 May 2003 | Population | General Population | Liang (2004) |
| Age, Sex | Significant | Unspecified | | China | 8 Mar 2003 - 28 May 2003 | Population | General Population | Liang (2007) |
| **Incidence** | | | | | | | | |
| Other | Significant | Unspecified | | China | 8 Mar 2003 - 28 May 2003 | Population | General Population | Liang (2007) |
| **Mortality** | | | | | | | | |
| Age | Significant | Unspecified | | China | 8 Mar 2003 - 28 May 2003 | Population | General Population | Liang (2007) |
| Sex | Not Significant | Unspecified | | China | 8 Mar 2003 - 28 May 2003 | Population | General Population | Liang (2007) |
| **Recovery** | | | | | | | | |
| Age, Cormobidity, Other | Significant | Adjusted | 88 | Hong Kong SAR; China | 9 Mar 2003 - 28 Apr 2003 | Hospital | General Population | Lau (2004e) |
| Age, Cormobidity, Other | Significant | Not Adjusted | 54 | | 12 Mar 2003 - 18 Apr 2003 | Hospital | Persons Under Investigation | Gomersall (2004) |
| Other, Sex | Not Significant | Adjusted | 88 | Hong Kong SAR; China | 9 Mar 2003 - 28 Apr 2003 | Hospital | General Population | Lau (2004e) |
| Other, Sex | Not Significant | Not Adjusted | 54 | | 12 Mar 2003 - 18 Apr 2003 | Hospital | Persons Under Investigation | Gomersall (2004) |
| **Fever being febrile** | | | | | | | | |
| Other | Significant | Unspecified | | Singapore | 22 Apr 2003 - 5 Jun 2003 | Hospital | Healthcare Workers | Ho (2004) |
| **Incubation Period** | | | | | | | | |
| Age, Occupation, Other | Significant | Adjusted | 317 | Canada; Hong Kong SAR; China | | Population | Persons Under Investigation | Cowling (2007) |
| Age, Sex | Not Significant | Adjusted | 317 | Canada; Hong Kong SAR; China | | Population | Persons Under Investigation | Cowling (2007) |
| **Admission-Death Delay** | | | | | | | | |
| Age | Significant | Unspecified | 345 | Taiwan; China | - 12 Oct 2003 | Hospital | Persons Under Investigation | Chang (2007) |
| **Admission-Discharge Delay** | | | | | | | | |
| Age | Significant | Unspecified | 345 | Taiwan; China | - 12 Oct 2003 | Hospital | Persons Under Investigation | Chang (2007) |
| **Incidence Rate** | | | | | | | | |
| Occupation, Other | Significant | Adjusted | 295 | Hong Kong SAR; China | 12 Apr 2003 - Jun 2003 | Population | General Population | Bucchianeri (2010) |
| Occupation, Other | Not Significant | Adjusted | 295 | Hong Kong SAR; China | 12 Apr 2003 - Jun 2003 | Population | General Population | Bucchianeri (2010) |

Table B.13: Published risk factors by outcome. Study characteristics are reported for each set of reported risk factors.

#### B.5 Additional Figures

##### B.5.1 Main text figures without quality assessment

Figures 3, 4, and 5 in the main manuscript only present parameters from papers with QA scores > 0.5. This section recreates these figures but without restrictions on QA scores. All extracted parameters are included.

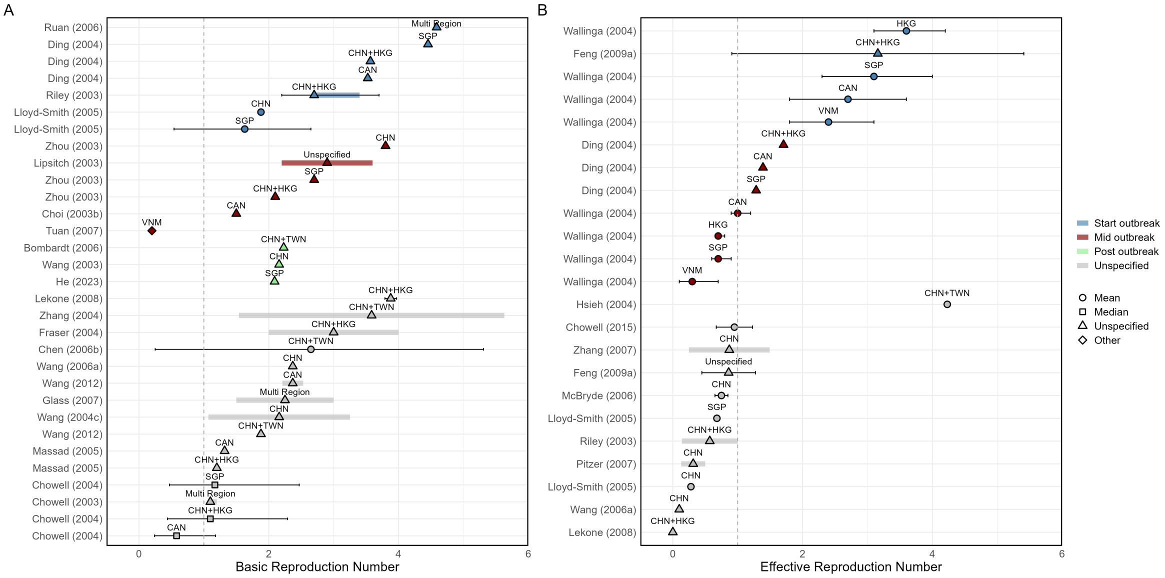

Figure B.4: Overview of estimated SARS (A) basic reproduction numbers ($R_0$) and (B) effective reproduction numbers ($R_e$). Note, that this Figure is analogous to Figure 3 in our main manuscript text, but with all parameters included, not just those from papers with QA scores > 0.5. Circles represent mean midpoint estimates, squares median midpoint estimates, diamonds "other" midpoint estimates, and triangles represent unspecified midpoint estimates. Thin solid lines represent uncertainty estimates, and solid shaded bars represent the range of central estimates when disaggregated by other parameters (e.g. age, sex, region, time). Colour represents when during the relative outbreak the study was conducted. Estimates are labelled with the country of study. CHN = China, HKG = Hong Kong, SGP = Singapore, CAN = Canada, TWN = Taiwan, VNM = Vietnam.

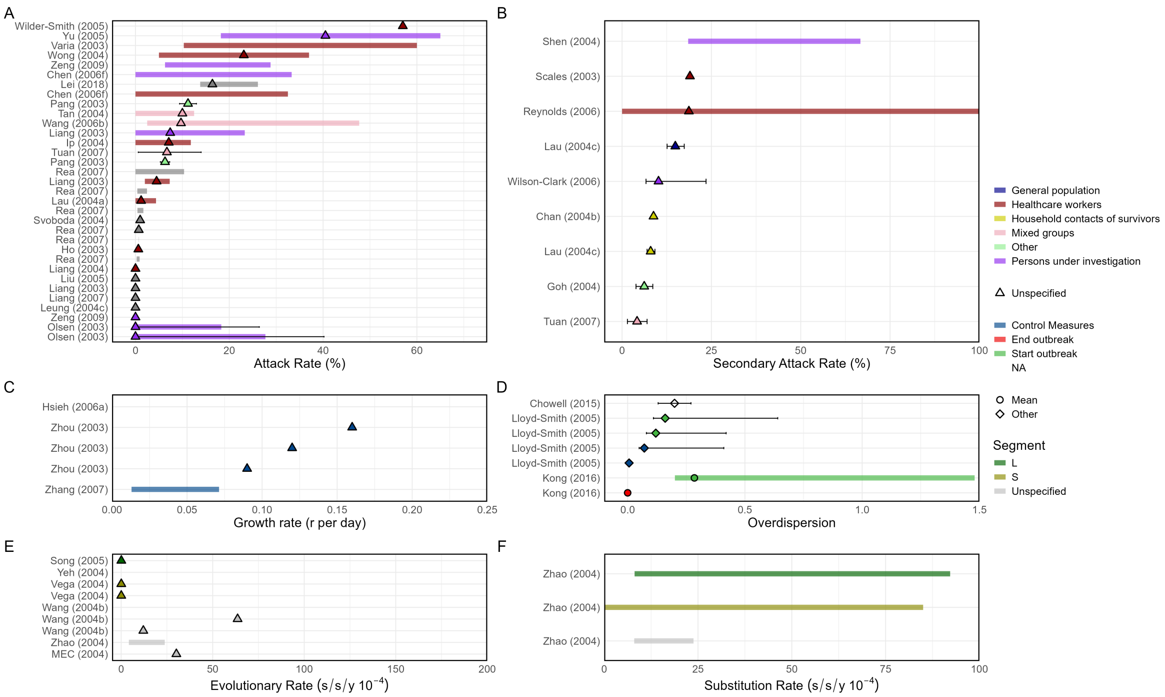

Figure B.5: Overview of estimated SARS (A) attack rates, (B) secondary attack rates, (C) growth rates, (D) overdispersion, (E) evolutionary rates, (F) substitution rates. Note, that this Figure is analogous to Figure 4 in our main manuscript text, but with all parameters included, not just those from papers with QA scores > 0.5. Circles represent mean midpoint estimates, diamonds represent "other" midpoint estimates, and triangles represent unspecified midpoint estimates. Thin solid lines represent uncertainty estimates, and solid shaded bars represent the range of central estimates when disaggregated by other parameters (e.g. age, sex, region, time), or broader unspecified parameter ranges. Colour represents (A & B) the study population considered, (C & D) when during the outbreak the study was conducted, where "control measures" refers to a time when interventions were reported to be in place, (E & F) long/short gene segment. S/s/y refers to nucleotide substitutions per site per year.

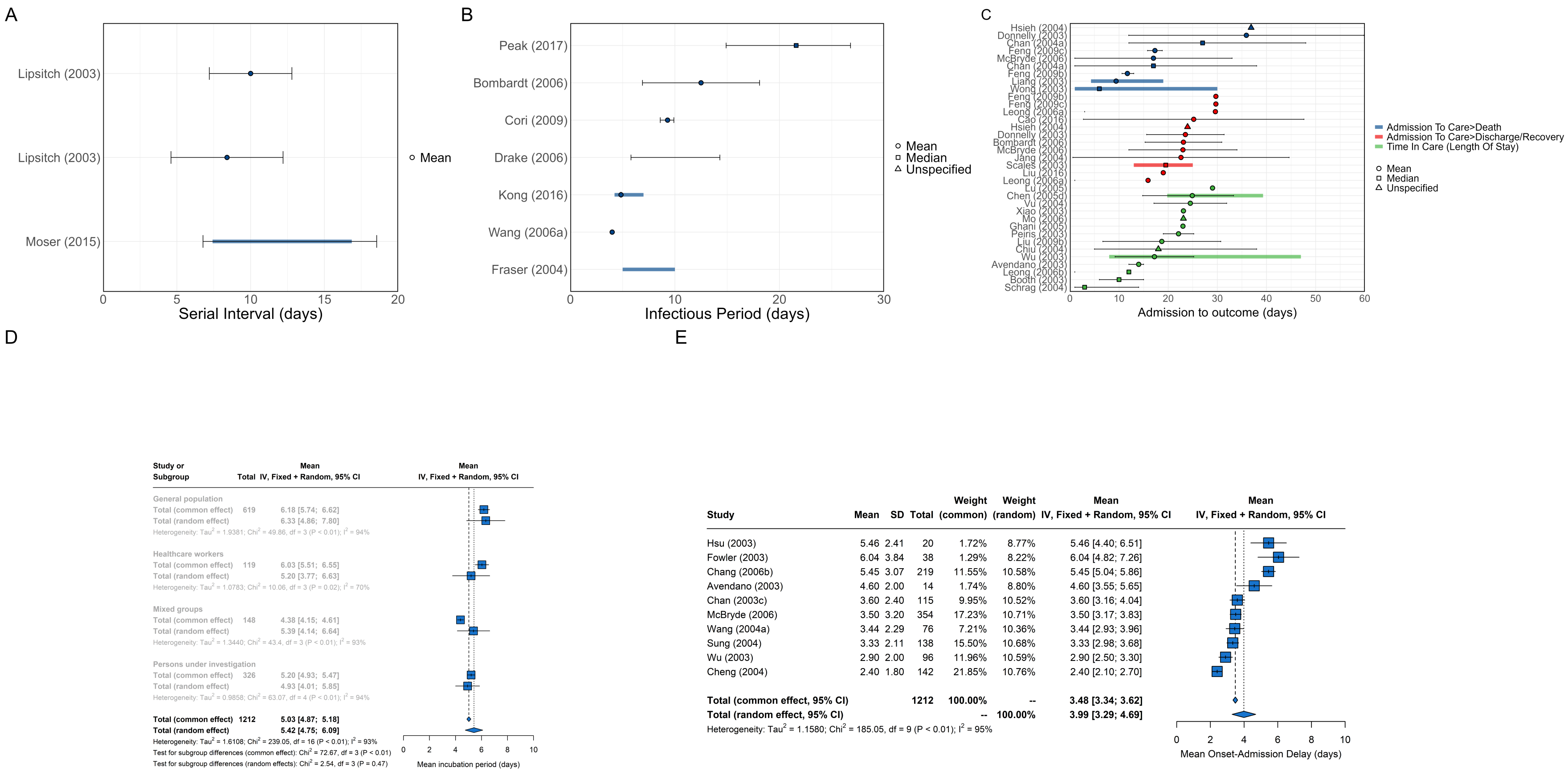

Figure B.6: Overview of SARS epidemiological delays – estimates of (A) serial interval, (B) infectious period, (C) time duration from hospital admission to final health outcome, and meta-analysis of epidemiological delays – central estimates of (D) incubation period, (E) time duration from symptom onset to hospital admission. Note, that this Figure is analogous Figure 5 in our main manuscript text, but with all parameters included, not just those from papers with QA scores > 0.5 (this has not impacted the meta-analysis as low quality studies had insufficient data to be included). In plots A, B, & C Circles represent mean midpoint estimates, squares represent median midpoint estimates, and triangles represent unspecified midpoint estimates. In plot C, colour represents different final health outcomes. In meta-analyses (D) blue squares indicate common effect and random effect estimates across different study populations, and blue diamonds represent: overall common effect estimates - in which all aggregated data are assumed to come from a single data-generating process, and overall random effect estimates, (E) blue squares represent study-specific estimates, and blue diamonds represent overall common effect and random effect estimates. Thin solid lines represent uncertainty estimates, and solid shaded bars represent the range of central estimates when disaggregated by other parameters (e.g. age, sex, region, time)

##### B.5.2 Alternate variable grouping

This section recreates figures from the main manuscript and elsewhere in the Supplementary Material, but visualing different grouping variables by colour. These plots are provided to aid interpretation of the role played by these grouping variables.

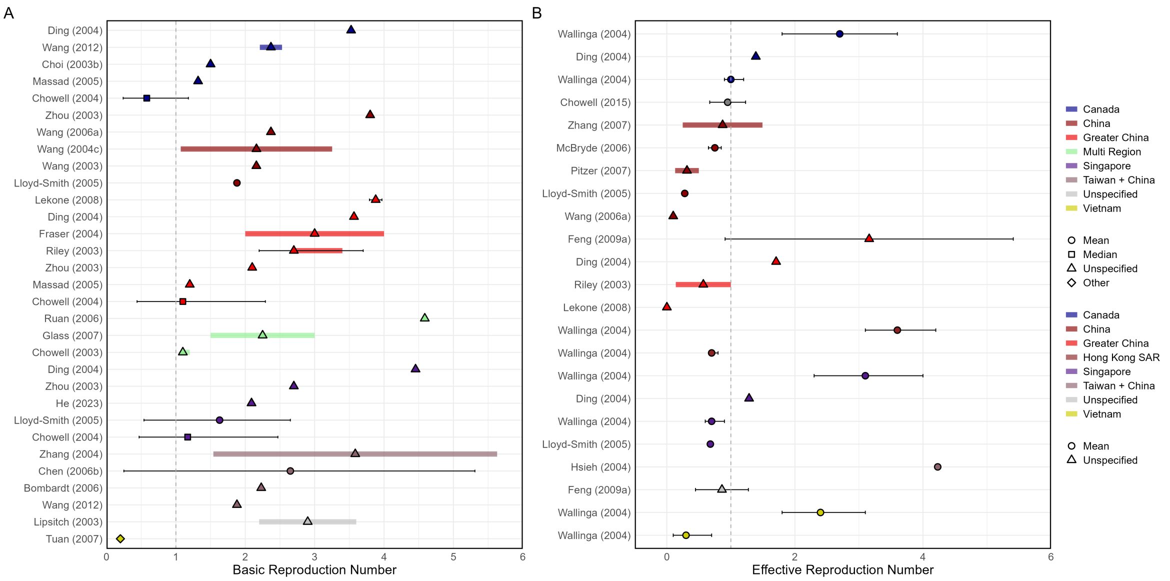

Figure B.7: Overview of estimated SARS (A) basic reproduction numbers ($R_0$) and (B) effective reproduction numbers ($R_e$). Note, that this Figure is analogous to Figure B.4 above, but where parameters are now colour-coded by study country, as opposed to when the study occurred. Circles represent mean midpoint estimates, squares median midpoint estimates, diamonds "other" midpoint estimates, and triangles represent unspecified midpoint estimates. Thin solid lines represent uncertainty estimates, and solid shaded bars represent the range of central estimates when disaggregated by other parameters (e.g. age, sex, region, time). Colour represents which country the study was conducted in.

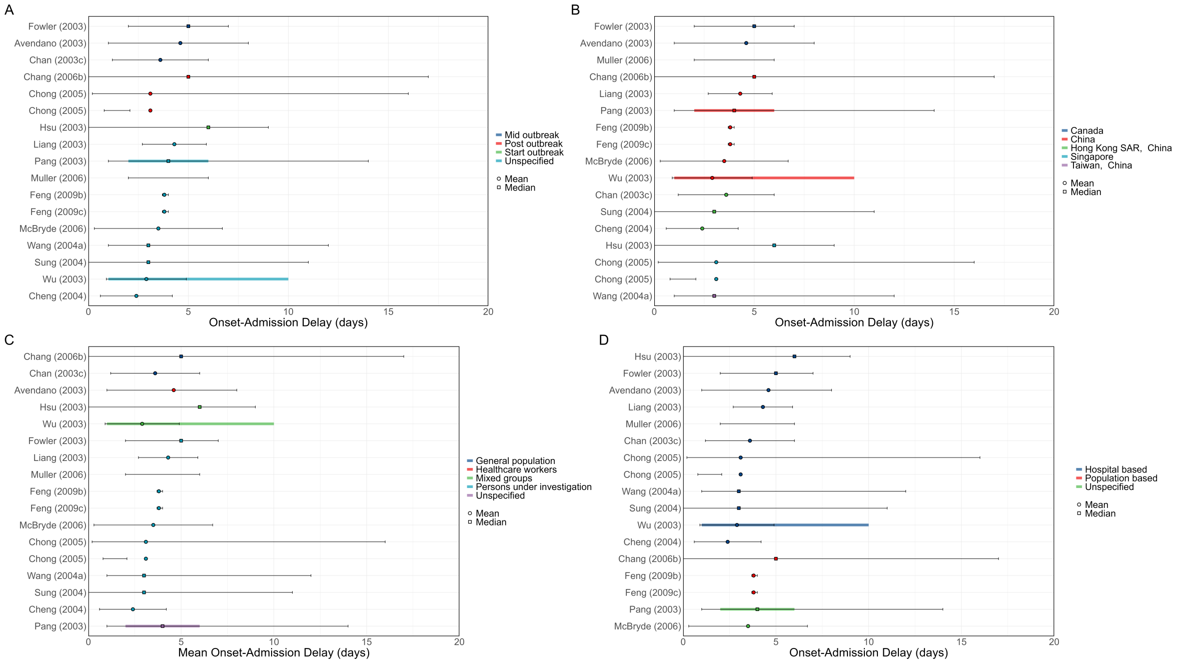

Figure B.8: Overview of estimated mean onset to admission delays sorted by different variables (A) outbreak phase (B) country (C) population group and (D) population setting for high QA score studies. Circles represent mean midpoint estimates, squares median midpoint estimates, diamonds “other” midpoint estimates, and triangles represent unspecified midpoint estimates. Thin solid lines represent uncertainty estimates, and solid shaded bars represent the range of central estimates when disaggregated by other parameters (e.g. age, sex, region, time). Colours represent the respective variables by which estimates are sorted by. Please note that Chong (2005) is plotted as stated in the paper (mean outside 95% CI).

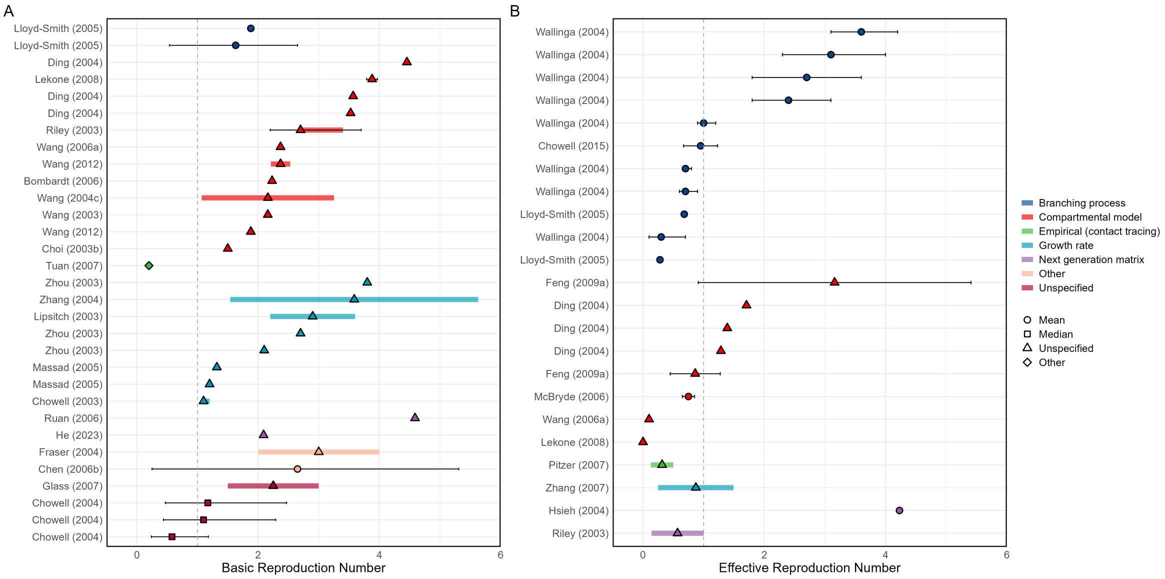

Figure B.9: Overview of estimated SARS (A) basic reproduction numbers ($R_0$) and (B) effective reproduction numbers ($R_e$). Note, that this Figure is analogous to Figure B.4 above, but where parameters are now colour-coded by the method by which the parameter was estimated, as opposed to when the study occurred. Circles represent mean midpoint estimates, squares median midpoint estimates, diamonds "other" midpoint estimates, and triangles represent unspecified midpoint estimates. Thin solid lines represent uncertainty estimates, and solid shaded bars represent the range of central estimates when disaggregated by other parameters (e.g. age, sex, region, time). Colour represents the method by which the parameter was estimated.

##### B.5.3 Individual study meta-analysis

In Figure 5D of the main text we plot the meta-analysis of mean incubation period estimates. In that Figure, blue squares represented only total common effects by study group. In Figure B.10 individual study effects are now also shown, presented as blue squares.

| Study or Subgroup | Mean | SD | Total | Weight (common) | Weight (random) | Mean IV, Fixed + Random, 95% CI |
|---|---|---|---|---|---|---|
| **General population** | | | | | | |
| Lau (2010) | 4.40 | 4.60 | 168 | 4.97% | 6.77% | 4.40 [3.70; 5.10] |
| Lau (2010) | 5.70 | 9.70 | 97 | 0.65% | 4.55% | 5.70 [3.77; 7.63] |
| Lau (2010) | 6.90 | 6.10 | 210 | 3.54% | 6.57% | 6.90 [6.07; 7.73] |
| Booth (2003) | 8.21 | 5.24 | 144 | 3.29% | 6.52% | 8.21 [7.35; 9.06] |
| **Total (common effect, 95% CI)** | | | **619** | **12.44%** | -- | **6.18 [5.74; 6.62]** |
| **Total (random effect, 95% CI)** | | | | -- | **24.42%** | **6.33 [4.86; 7.80]** |
| Heterogeneity: $Tau^2 = 1.9381$; $Chi^2 = 49.86$, df = 3 ($P < 0.01$); $I^2 = 94\%$ | | | | | | |
| **Healthcare workers** | | | | | | |
| Scales (2003) | 6.43 | 5.18 | 7 | 0.16% | 2.16% | 6.43 [2.60; 10.27] |
| Liu (2016) | 6.32 | 2.81 | 98 | 7.78% | 6.95% | 6.32 [5.77; 6.88] |
| Avendano (2003) | 4.00 | 3.00 | 4 | 0.28% | 3.04% | 4.00 [1.06; 6.94] |
| Avendano (2003) | 3.50 | 3.00 | 10 | 0.70% | 4.68% | 3.50 [1.64; 5.36] |
| **Total (common effect, 95% CI)** | | | **119** | **8.91%** | -- | **6.03 [5.51; 6.55]** |
| **Total (random effect, 95% CI)** | | | | -- | **16.84%** | **5.20 [3.77; 6.63]** |
| Heterogeneity: $Tau^2 = 1.0783$; $Chi^2 = 10.06$, df = 3 ($P = 0.02$); $I^2 = 70\%$ | | | | | | |
| **Mixed groups** | | | | | | |
| Wu (2003) | 5.90 | 3.50 | 96 | 4.91% | 6.76% | 5.90 [5.20; 6.60] |
| Hsu (2003) | 4.41 | 2.22 | 7 | 0.89% | 5.08% | 4.41 [2.77; 6.05] |
| Hsu (2003) | 7.33 | 2.40 | 13 | 1.42% | 5.72% | 7.33 [6.03; 8.64] |
| Chen (2003) | 4.08 | 0.72 | 32 | 38.15% | 7.22% | 4.08 [3.83; 4.33] |
| **Total (common effect, 95% CI)** | | | **148** | **45.37%** | -- | **4.38 [4.15; 4.61]** |
| **Total (random effect, 95% CI)** | | | | -- | **24.79%** | **5.39 [4.14; 6.64]** |
| Heterogeneity: $Tau^2 = 1.3440$; $Chi^2 = 43.4$, df = 3 ($P < 0.01$); $I^2 = 93\%$ | | | | | | |
| **Persons under investigation** | | | | | | |
| Varia (2003) | 4.36 | 1.83 | 42 | 7.82% | 6.95% | 4.36 [3.80; 4.91] |
| Wong (2004) | 3.35 | 1.26 | 11 | 4.34% | 6.70% | 3.35 [2.60; 4.09] |
| McBryde (2006) | 5.30 | 4.50 | 85 | 2.63% | 6.36% | 5.30 [4.34; 6.26] |
| Lee (2003) | 6.46 | 2.68 | 138 | 12.02% | 7.07% | 6.46 [6.01; 6.90] |
| Goh (2006) | 5.10 | 2.20 | 50 | 6.47% | 6.88% | 5.10 [4.49; 5.71] |
| **Total (common effect, 95% CI)** | | | **326** | **33.28%** | -- | **5.20 [4.93; 5.47]** |
| **Total (random effect, 95% CI)** | | | | -- | **33.96%** | **4.93 [4.01; 5.85]** |
| Heterogeneity: $Tau^2 = 0.9858$; $Chi^2 = 63.07$, df = 4 ($P < 0.01$); $I^2 = 94\%$ | | | | | | |
| **Total (common effect, 95% CI)** | | | **1212** | **100.00%** | **--** | **5.03 [4.87; 5.18]** |
| **Total (random effect, 95% CI)** | | | | **--** | **100.00%** | **5.42 [4.75; 6.09]** |
| Heterogeneity: $Tau^2 = 1.6108$; $Chi^2 = 239.05$, df = 16 ($P < 0.01$); $I^2 = 93\%$ | | | | | | |
| Test for subgroup differences (common effect): $Chi^2 = 72.67$, df = 3 ($P < 0.01$) | | | | | | |
| Test for subgroup differences (random effects): $Chi^2 = 2.54$, df = 3 ($P = 0.47$) | | | | | | |

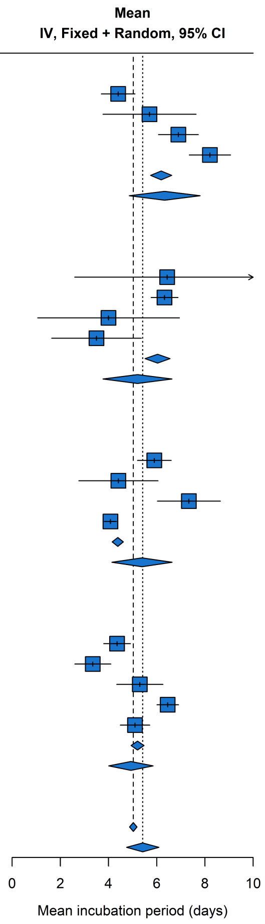

Figure B.10: A meta-analysis of the mean incubation period, subgrouped by the specific type of population studied. Blue squares represent study-specific estimates, and blue diamonds represent: common effect estimates - in which all aggregated data are assumed to come from a single data-generating process, and random effect estimates. Thin solid lines represent uncertainty estimates. Note, this figure only includes studies with a QA score greater than 0.5.

##### B.5.4 Risk factor plots

In this section we plot the reported risk factors for different epidemiological outcomes. This includes both significant and not significant risk factor reportings.

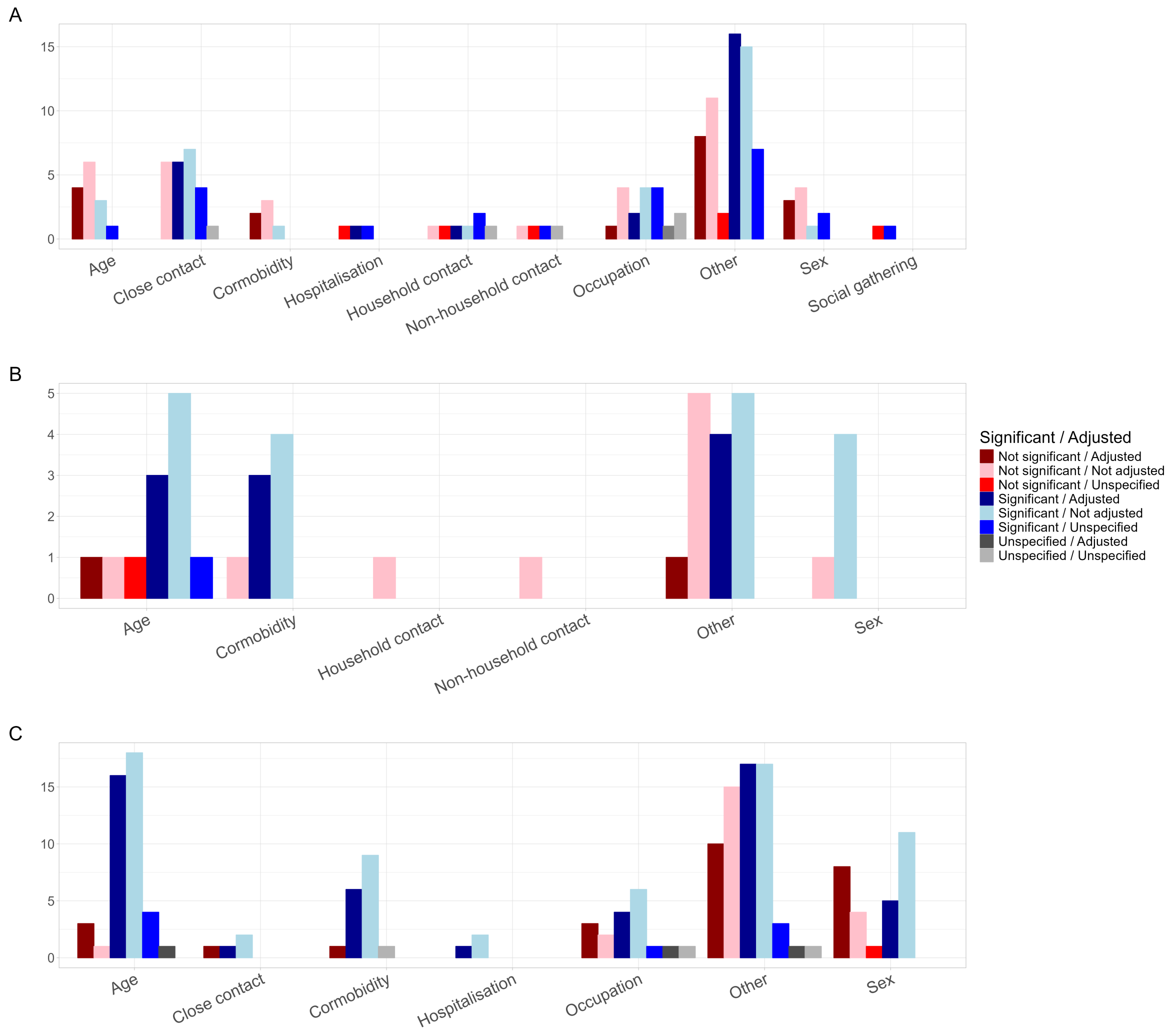

Figure B.11: Tally of reported significant and not significant risk factors associated with (A) infection, (B) severe disease, (C) death. Red shades indicate a non-significant relationship, blue shades represent a significant relationship, and grey shades an unspecified relationship. Different shades of these colours indicate whether the method used constitutes an adjusted or non-adjusted analysis. All risk factor studies are included here, regardless of QA score.

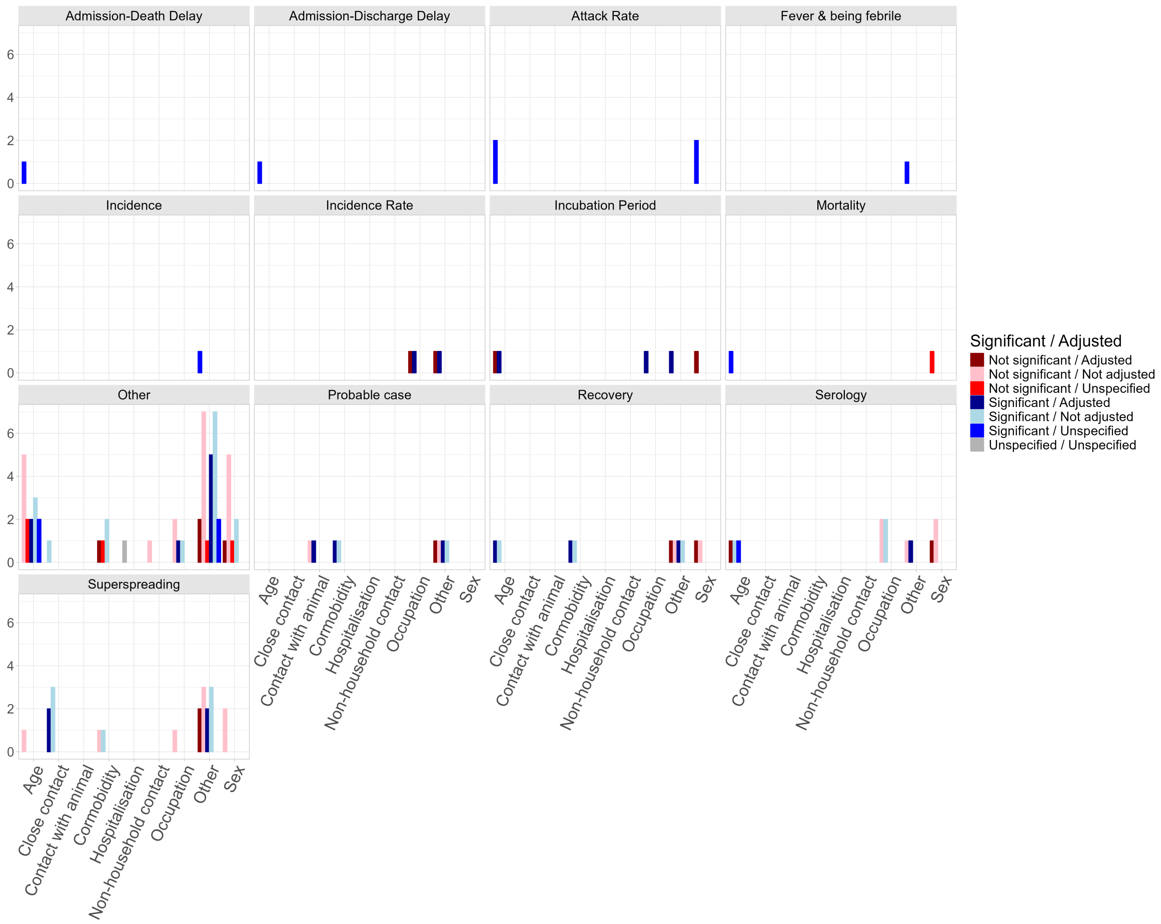

Figure B.12: Tally of reported significant and not significant risk factors associated with all other epidemiological phenomena and health outcomes. Red shades indicate a non-significant relationship, blue shades represent a significant relationship, and grey shades an unspecified relationship. Different shades of these colours indicate whether the method used constitutes an adjusted or non-adjusted analysis. All risk factor studies are included here, regardless of QA score.

#### B.6 Screening statistics

Both the initial screening and the full text review stages of study selection were conducted by two reviewers. This section reports the degrees of concordance between reviewers in this stage.

**Screening**

| Reviewer A | Reviewer B | n | Proportionate Agreement | Yes Probability | No Probability | Random Agreement Probability | Cohen's Kappa |
|---|---|---|---|---|---|---|---|
| P1 | P2 | 1768 | 89% | 1% | 79% | 80% | 46% |
| P3 | P4 | 1427 | 96% | 0% | 93% | 93% | 50% |
| P3 | P1 | 664 | 94% | 1% | 84% | 85% | 61% |
| P4 | P5 | 729 | 95% | 0% | 88% | 88% | 55% |
| P5 | P1 | 335 | 94% | 0% | 88% | 89% | 47% |
| P3 | P2 | 1329 | 89% | 2% | 76% | 78% | 52% |
| P4 | P1 | 3037 | 99% | 0% | 97% | 97% | 80% |
| P3 | P5 | 306 | 96% | 0% | 90% | 90% | 65% |
| P5 | P2 | 631 | 88% | 1% | 80% | 81% | 38% |
| P4 | P2 | 1338 | 93% | 1% | 78% | 80% | 64% |
| P2 | P6 | 337 | 84% | 2% | 72% | 74% | 41% |
| P7 | P8 | 2408 | 98% | 0% | 96% | 96% | 43% |
| P7 | P9 | 608 | 97% | 0% | 91% | 91% | 64% |

**Full Text**

| Reviewer A | Reviewer B | n | Proportionate Agreement | Yes Probability | No Probability | Random Agreement Probability | Cohen's Kappa |
|---|---|---|---|---|---|---|---|
| P3 | P1 | 166 | 85% | 26% | 24% | 50% | 70% |
| P5 | P1 | 124 | 75% | 22% | 28% | 50% | 50% |
| P3 | P5 | 175 | 86% | 18% | 33% | 51% | 71% |
| P4 | P1 | 101 | 85% | 12% | 43% | 55% | 67% |
| P7 | P8 | 136 | 97% | 1% | 83% | 84% | 82% |
| P4 | P5 | 22 | 82% | 20% | 29% | 49% | 65% |
| P3 | P4 | 139 | 86% | 5% | 61% | 66% | 58% |

Figure B.13: Cohen's Kappa for screening and full text review. P1-P9 are members of PERG (see appendix G), and we have provided who participated at each stage in the methods section of the main text. $n$ is the number of studies considered by each Reviewer A/B pair. We highlight that *all* studies were screened by two people and any conflicts were resolved by discussion and mutual agreement of Reviewer A and B. The same applies to the subsequent Full text reviews.

### C The `epireview` package

In parallel with this work, we developed an R package called `epireview` to provide a central location to host and access the extracted data for the nine priority pathogens in our review series. The package allows for submissions of outbreak, model, and parameter data from new peer-reviewed papers via pull requests, and includes functions to produce the tables and figures included in this paper and update them with any additional data. This package will be updated as the overall project by the Pathogen Epidemiology Review Group (PERG) continues to extract modelling parameters for the rest of the nine priority pathogens as defined by WHO.

There are several vignettes available:

- A vignette for Marburg Virus Disease, Lassa fever and SARS-CoV-1 with tables and figures from the respective papers, which will be updated as data are added to the database.
- A vignette that lists the options for each outbreak, model, or parameter field and describes how to access them using a function in the package.
- A vignette to explain the process of updating the database with new article, outbreak, model, or parameter data.

### D PRISMA 2020 Checklists

TO BE UPDATED

| Section & Topic | Item # | Checklist item | Reported (Yes/No) |
|---|---|---|---|
| **Title** | | | |
| Title | 1 | Identify the report as a systematic review. | Yes |
| **Background** | | | |
| Objectives | 2 | Provide an explicit statement of the main objective(s) or question(s) the review addresses. | Yes |
| **Methods** | | | |
| Eligibility criteria | 3 | Specify the inclusion and exclusion criteria for the review. | Yes |
| Information sources | 4 | Specify the information sources (e.g. databases, registers) used to identify studies and the date when each was last searched. | Yes |
| Risk of bias | 5 | Specify the methods used to assess risk of bias in the included studies. | Yes |
| Synthesis of results | 6 | Specify the methods used to present and synthesise results. | Yes |
| **Results** | | | |
| Included studies | 7 | Give the total number of included studies and participants and summarise relevant characteristics of studies. | Yes |
| Synthesis of results | 8 | Present results for main outcomes, preferably indicating the number of included studies and participants for each. If meta-analysis was done, report the summary estimate and confidence/credible interval. If comparing groups, indicate the direction of the effect (i.e. which group is favoured). | Yes |
| **Discussion** | | | |
| Limitations of evidence | 9 | Provide a brief summary of the limitations of the evidence included in the review (e.g. study risk of bias, inconsistency and imprecision). | Yes |
| Interpretation | 10 | Provide a general interpretation of the results and important implications. | Yes |
| **Other** | | | |
| Funding | 11 | Specify the primary source of funding for the review. | Yes |
| Registration | 12 | Provide the register name and registration number. | Yes |

Table D.14: PRISMA 2020 Abstracts Checklist. ([10])

| Section & Topic | Item # | Checklist item | Location where item is reported |
|---|---|---|---|
| **Title** | | | |
| Title | 1 | Identify the report as a systematic review. | page 1 |
| **Abstract** | | | |
| Abstract | 2 | See the PRISMA 2020 for Abstracts checklist. | Table D.14 |
| **Introduction** | | | |
| Rationale | 3 | Describe the rationale for the review in the context of existing knowledge. | page 2 |
| Objectives | 4 | Provide an explicit statement of the objective(s) or question(s) the review addresses. | page 2/3 |
| **Methods** | | | |
| Eligibility criteria | 5 | Specify the inclusion and exclusion criteria for the review and how studies were grouped for the syntheses. | page 3 |

Table D.15: PRISMA 2020 Checklist. ([10])

| Section & Topic | Item # | Checklist item | Location where item is reported |
|---|---|---|---|
| Information sources | 6 | Specify all databases, registers, websites, organisations, reference lists and other sources searched or consulted to identify studies. Specify the date when each source was last searched or consulted. | page 3 |
| Search strategy | 7 | Present the full search strategies for all databases, registers and websites, including any filters and limits used. | page 3 + Figure 1 |
| Selection process | 8 | Specify the methods used to decide whether a study met the inclusion criteria of the review, including how many reviewers screened each record and each report retrieved, whether they worked independently, and if applicable, details of automation tools used in the process. | page 3 |
| Data collection process | 9 | Specify the methods used to collect data from reports, including how many reviewers collected data from each report, whether they worked independently, any processes for obtaining or confirming data from study investigators, and if applicable, details of automation tools used in the process. | page 3/4 |
| Data items | 10a | List and define all outcomes for which data were sought. Specify whether all results that were compatible with each outcome domain in each study were sought (e.g. for all measures, time points, analyses), and if not, the methods used to decide which results to collect. | page 3/4 |
| | 10b | List and define all other variables for which data were sought (e.g. participant and intervention characteristics, funding sources). Describe any assumptions made about any missing or unclear information. | page 3/4 |
| Study risk of bias assessment | 11 | Specify the methods used to assess risk of bias in the included studies, including details of the tool(s) used, how many reviewers assessed each study and whether they worked independently, and if applicable, details of automation tools used in the process. | page 4 |
| Effect measures | 12 | Specify for each outcome the effect measure(s) (e.g. risk ratio, mean difference) used in the synthesis or presentation of results. | page 3/4 |
| Synthesis methods | 13a | Describe the processes used to decide which studies were eligible for each synthesis (e.g. tabulating the study intervention characteristics and comparing against the planned groups for each synthesis (item #5)). | page 4 |
| | 13b | Describe any methods required to prepare the data for presentation or synthesis, such as handling of missing summary statistics, or data conversions. | page 4 |
| | 13c | Describe any methods used to tabulate or visually display results of individual studies and syntheses. | page 4 |
| | 13d | Describe any methods used to synthesize results and provide a rationale for the choice(s). If meta-analysis was performed, describe the model(s), method(s) to identify the presence and extent of statistical heterogeneity, and software package(s) used. | page 4 |
| | 13e | Describe any methods used to explore possible causes of heterogeneity among study results (e.g. subgroup analysis, meta-regression). | - |
| | 13f | Describe any sensitivity analyses conducted to assess robustness of the synthesized results. | - |
| Reporting bias assessment | 14 | Describe any methods used to assess risk of bias due to missing results in a synthesis (arising from reporting biases). | page 4 |
| Certainty assessment | 15 | Describe any methods used to assess certainty (or confidence) in the body of evidence for an outcome. | page 4 |
| **Results** | | | |
| Study selection | 16a | Describe the results of the search and selection process, from the number of records identified in the search to the number of studies included in the review, ideally using a flow diagram. | page 4 |
| | 16b | Cite studies that might appear to meet the inclusion criteria, but which were excluded, and explain why they were excluded. | Table ?? |
| Study characteristics | 17 | Cite each included study and present its characteristics. | pages 4-13 |
| Risk of bias in studies | 18 | Present assessments of risk of bias for each included study. | page 7 |
| Results of individual studies | 19 | For all outcomes, present, for each study: (a) summary statistics for each group (where appropriate) and (b) an effect estimate and its precision (e.g. confidence/credible interval), ideally using structured tables or plots. | pages 4-13 |

Table D.15: PRISMA 2020 Checklist. ([10])

| Section & Topic | Item # | Checklist item | Location where item is reported |
|---|---|---|---|
| Results of syntheses | 20a | For each synthesis, briefly summarise the characteristics and risk of bias among contributing studies. | page 6/7 |
| | 20b | Present results of all statistical syntheses conducted. If meta-analysis was done, present for each the summary estimate and its precision (e.g. confidence/credible interval) and measures of statistical heterogeneity. If comparing groups, describe the direction of the effect. | page 4-13 |
| | 20c | Present results of all investigations of possible causes of heterogeneity among study results. | page 4-13 |
| | 20d | Present results of all sensitivity analyses conducted to assess the robustness of the synthesized results. | page 4-13 |
| Reporting biases | 21 | Present assessments of risk of bias due to missing results (arising from reporting biases) for each synthesis assessed. | page 7 |
| Certainty of evidence | 22 | Present assessments of certainty (or confidence) in the body of evidence for each outcome assessed. | pages 4-13 |
| **Discussion** | | | |
| Discussion | 23a | Provide a general interpretation of the results in the context of other evidence. | page 14/15 |
| | 23b | Discuss any limitations of the evidence included in the review. | page 14-15 |
| | 23c | Discuss any limitations of the review processes used. | page 14-15 |
| | 23d | Discuss implications of the results for practice, policy, and future research. | page 14-15 |
| **Other Information** | | | |
| Registration and protocol | 24a | Provide registration information for the review, including register name and registration number, or state that the review was not registered. | page 14 |
| | 24b | Indicate where the review protocol can be accessed, or state that a protocol was not prepared. | page 16 |
| | 24c | Describe and explain any amendments to information provided at registration or in the protocol. | n/a |
| Support | 25 | Describe sources of financial or non-financial support for the review, and the role of the funders or sponsors in the review. | page 15/16 |
| Competing interests | 26 | Declare any competing interests of review authors. | page 16 |
| Availability of data, code and other materials | 27 | Report which of the following are publicly available and where they can be found: template data collection forms; data extracted from included studies; data used for all analyses; analytic code; any other materials used in the review. | page 16 |

Table D.15: PRISMA 2020 Checklist. ([10])

### E Other Systematic Reviews

Systematic reviews were excluded from our analysis, but the systematic reviews that were identified in the database search and used for validation purposes are presented in Table E.16.

| Study | Title | Journal | DOI |
|---|---|---|---|
| Vink 2014 | Serial intervals of respiratory infectious diseases: a systematic review and analysis | Am J Epidemiol | 10.1093/aje/kwu209 |
| Tran 2012 | Aerosol generating procedures and risk of transmission of acute respiratory infections to healthcare workers: a systematic review | PLoS One | 10.1371/journal.pone.0035797 |
| Olowokure 2004 | SARS | Nat Rev Microbiol | 10.1038/nrmicro824 |
| Kwok 2019 | Epidemic Models of Contact Tracing: Systematic Review of Transmission Studies of Severe Acute Respiratory Syndrome and Middle East Respiratory Syndrome | Comput Struct Biotechnol J | 10.1016/j.csbj.2019.01.003 |
| Jefferson 2008 | Physical interventions to interrupt or reduce the spread of respiratory viruses: systematic review | Bmj | 10.1136/bmj.39393.510347.BE |
| Lessler 2009 | Incubation periods of acute respiratory viral infections: a systematic review | Lancet Infect Dis | 10.1016/s1473-3099(09)70069-6 |
| Donnelly 2004 | Epidemiological and genetic analysis of severe acute respiratory syndrome | Lancet Infect Dis | 10.1016/s1473-3099(04)01173-9 |
| Bin-Reza 2012 | The use of masks and respirators to prevent transmission of influenza: a systematic review of the scientific evidence | Influenza Other Respir Viruses | 10.1111/j.1750-2659.2011.00307.x |
| Chan 2021 | Transmission of Severe Acute Respiratory Syndrome Coronavirus 1 and Severe Acute Respiratory Syndrome Coronavirus 2 During Aerosol-Generating Procedures in Critical Care: A Systematic Review and Meta-Analysis of Observational Studies* | CRITICAL CARE MEDICINE | 10.1097/CCM.0000000000004965 |
| BinNafisah 2021 | The risk of coronavirus to healthcare providers during aerosol-generating procedures: A systematic review and meta-analysis | ANNALS OF THORACIC MEDICINE | 10.4103/atm.ATM_497_20 |
| Barman 2022 | Respiratory rehabilitation in patients recovering from severe acute respiratory syndrome: A systematic review and meta-analysis. | Heart Lung | 10.1016/j.hrtlng.2022.01.005 |
| Yiwen 2010 | A comprehensive systematic review of healthcare workers' perceptions of risk from exposure to emerging acute respiratory infectious diseases and the perceived effectiveness of strategies used to facilitate healthy coping in acute hospital and community healthcare settings | JBI Libr Syst Rev | |
| Stockman 2006 | SARS: systematic review of treatment effects | PLoS Med | 10.1371/journal.pmed.0030343 |
| Pedrosa 2011 | Viral infections in workers in hospital and research laboratory settings: a comparative review of infection modes and respective biosafety aspects | Int J Infect Dis | 10.1016/j.ijid.2011.03.005 |
| Patarcic 2015 | The role of host genetic factors in respiratory tract infectious diseases: systematic review, meta-analyses and field synopsis | Sci Rep | 10.1038/srep16119 |
| Offeddu 2017 | Effectiveness of Masks and Respirators Against Respiratory Infections in Healthcare Workers: A Systematic Review and Meta-Analysis | Clin Infect Dis | 10.1093/cid/cix681 |
| Mair-Jenkins 2015 | The effectiveness of convalescent plasma and hyperimmune immunoglobulin for the treatment of severe acute respiratory infections of viral etiology: a systematic review and exploratory meta-analysis | J Infect Dis | 10.1093/infdis/jiu396 |

Table E.16: Other reviews of SARS identified during manuscript screening.

| Study | Title | Journal | DOI |
|---|---|---|---|
| Momattin 2013 | Therapeutic options for Middle East respiratory syndrome coronavirus (MERS-CoV)–possible lessons from a systematic review of SARS-CoV therapy | Int J Infect Dis | 10.1016/j.ijid.2013.07.002 |
| Liu 2004 | Chinese herbal medicine for severe acute respiratory syndrome: a systematic review and meta-analysis | J Altern Complement Med | 10.1089/acm.2004.10.1041 |
| Li 2007 | Role of ventilation in airborne transmission of infectious agents in the built environment - a multidisciplinary systematic review | Indoor Air | 10.1111/j.1600-0668.2006.00445.x |
| Kramer 2006 | How long do nosocomial pathogens persist on inanimate surfaces? A systematic review | BMC Infect Dis | 10.1186/1471-2334-6-130 |
| Koh 2011 | Comprehensive systematic review of healthcare workers' perceptions of risk and use of coping strategies towards emerging respiratory infectious diseases | Int J Evid Based Healthc | 10.1111/j.1744-1609.2011.00242.x |
| Jefferson 2009 | Physical interventions to interrupt or reduce the spread of respiratory viruses: systematic review | Bmj | 10.1136/bmj.b3675 |
| Browne 2016 | The roles of transportation and transportation hubs in the propagation of influenza and coronaviruses: a systematic review | J Travel Med | 10.1093/jtm/tav002 |
| Teasdale 2014 | Public perceptions of non-pharmaceutical interventions for reducing transmission of respiratory infection: systematic review and synthesis of qualitative studies | BMC Public Health | 10.1186/1471-2458-14-589 |

Table E.16: Other reviews of SARS identified during manuscript screening.

### F Excluded Studies

We list all excluded studies with reasons for exclusion in the Priority Pathogens GitHub repo. The full list can be access at `https://github.com/mrc-ide/priority-pathogens/blob/main/data/SARS-CoV-1_excluded_studies.csv`

| Title | Pubmed link |
|---|---|
| Efficiency of the quarantine system during the epidemic of severe acute respiratory syndrome in Beijing, 2003 | https://pubmed.ncbi.nlm.nih.gov/14761622/ |
| Epidemiologic features, clinical diagnosis and therapy of first cluster of patients with severe acute respiratory syndrome in Beijing area | https://pubmed.ncbi.nlm.nih.gov/12899773/ |
| Study on the epidemiology and measures for control on severe acute respiratory syndrome in Guangzhou city | https://pubmed.ncbi.nlm.nih.gov/12820926/ |
| An epidemiological study on the index cases of severe acute respiratory syndrome occurred in different cities among Guangdong province | https://pubmed.ncbi.nlm.nih.gov/12820924/ |
| Effectiveness of personal protective measures in prevention of nosocomial transmission of severe acute respiratory syndrome | https://pubmed.ncbi.nlm.nih.gov/15061941/ |

Table F.17: Excluded Chinese language papers identified during reverse search of other systematic reviews.

### G Pathogen Epidemiology Review Group (PERG) membership

| First name | Surname | Affiliation |
|---|---|---|
| Aaron | Morris | University of Oxford |
| Alpha | Forna | University of Georgia |
| Amy | Dighe | Johns Hopkins |
| Anna | Vicco | Imperial College London |
| Anna-Maria | Hartner | Imperial College London |
| Anne | Cori | Imperial College London |
| Arran | Hamlet | Imperial College London |
| Ben | Lambert | University of Oxford |
| Bethan | Cracknell Daniels | Imperial College London |
| Charles | Whittaker | Imperial College London |
| Christian | Morgenstern | Imperial College London |
| Cosmo | Santoni | Imperial College London |
| Cyril | Geismar | Imperial College London |
| Dariya | Nikitin | Imperial College London |
| David | Jorgensen | Imperial College London |
| Dominic | Dee | Imperial College London |
| Ed | Knock | Imperial College London |
| Gina | Cuomo-Dannenburg | Imperial College London |
| Hayley | Thompson | PATH |
| Ilaria | Dorigatti | Imperial College London |
| Isobel | Routledge | UCSF |
| Jack | Wardle | Imperial College London |
| Janetta | Skarp | Imperial College London |
| Joseph | Hicks | Imperial College London |
| Juliette | Unwin | University of Bristol |
| Kanchan | Parchani | Imperial College London |
| Keith | Fraser | Imperial College London |
| Kelly | Charniga | Imperial College London |
| Kelly | McCain | Imperial College London |
| Kieran | Drake | Imperial College London |
| Lily | Geidelberg | Imperial College London |
| Lorenzo | Cattarino | UKHSA |
| Mantra | Kusumgar | Imperial College London |
| Mara | Kont | Imperial College London |
| Marc | Baguelin | Imperial College London |
| Natsuko | Imai-Eaton | Wellcome Trust |
| Pablo | Perez Guzman | Imperial College London |
| Patrick | Doohan | Imperial College London |
| Paul | Lietar | Imperial College London |
| Paula | Christen | Imperial College London |
| Rebecca | Nash | Imperial College London |
| Rich | Fitzjohn | Imperial College London |

| | | |
|---|---|---|
| Richard | Sheppard | Imperial College London |
| Rob | Johnson | Imperial College London |
| Ruth | McCabe | Imperial College London |
| Sabine | van Elsland | Imperial College London |
| Sangeeta | Bhatia | UK Health Security Agency |
| Sequoia | Leuba | Imperial College London |
| Shazia | Ruybal-Pesantez | Imperial College London |
| Sreejith | Radhakrishnan | University of Glasgow |
| Thomas | Rawson | Imperial College London |
| Tristan | Naidoo | Imperial College London |
| Zulma | Cucunuba Perez | Pontificia Universidad Javeriana |

Table G.18: Pathogen Epidemiology Review Group (PERG) membership as of July 2024.
